# A rapid review exploring the effectiveness of artificial intelligence for cancer diagnosis

**DOI:** 10.1101/2023.11.09.23298257

**Authors:** Alesha Wale, Hannah Shaw, Toby Ayres, Chukwudi Okolie, Helen Morgan, Jordan Everitt, Kirsty Little, Rhiannon Tudor Edwards, Jacob Davies, Ruth Lewis, Alison Cooper, Adrian Edwards

**Affiliations:** Public Health Wales Evidence Service, Wales, United Kingdom; Centre for Health Economics & Medicines Evaluation, Bangor University, United Kingdom; Health and Care Research Wales Evidence Centre, Bangor University, United Kingdom; Health and Care Research Wales Evidence Centre, Cardiff University, United Kingdom

## Abstract

There is growing demand for diagnostic services in the UK. This rapid review aimed to assess the effectiveness of artificial intelligence (AI) in diagnostic radiology with a focus on cancer diagnosis. A range of AI models including machine learning, deep learning and ensemble models, were assessed in this review.

The review included an initial broad mapping exercise and a more in-depth synthesis of a specific sub-set of the evidence. The review included evidence available from 2018 until June 2023.

A total of 92 comparative primary studies were included in the evidence map. The evidence map identified 52 studies in which the AI models were in the early stages of development and validation, and highlighted breast, lung and prostate cancers as the type of cancers most frequently reported on. 28 studies evaluating an established model and focusing on the diagnosis of breast, lung, and prostate cancer were included in the in-depth synthesis. All studies included in the in-depth synthesis were classified as diagnostic accuracy studies. Only one study evaluated an AI model that was commercially available in the UK.

Most studies reported results in favour of the AI models, however, these improvements were not always statistically significant. The studies also varied considerably in terms of AI models studied, type of cancer, images used, and comparison made; and were limited in terms of their methodology. When used as a standalone diagnostic tool, there is evidence to suggest that AI can improve diagnostic accuracy or is comparable to experienced radiologists, however this may be dependent on the AI model being used. There is evidence to suggest that AI may be beneficial when used as a support tool for clinicians/radiologists with less experience. The impact of AI on the timeline involved in diagnosis appeared inconsistent. AI may speed up the diagnostic timeline when the level of cancer suspicion is low but may increase diagnostic timelines when the level of cancer suspicion is high. The evidence suggests that clinicians are accepting of AI-based assistance for cancer diagnosis.

**Policy and practice implications:** The overall evidence for effectiveness appeared in favour of AI and several factors were identified that impact the effectiveness of the AI models. AI may improve diagnostic accuracy in clinicians/radiologists with less experience of interpreting radiological images. However, further well-designed high-quality research is needed from the UK and similar countries to better understand the effectiveness of AI in cancer diagnosis.

**Economic considerations:** There is little evidence on the cost-effectiveness of using AI for cancer diagnosis. In theory, it might be possible for AI to assist with earlier diagnosis of cancer with both health and economic benefits.

**Funding statement:** The Public Health Wales Observatory was funded for this work by the Health and Care Research Wales Evidence Centre, itself funded by Health and Care Research Wales on behalf of Welsh Government.

**EXECUTIVE SUMMARY:** *What is a Rapid Review?:* Our rapid reviews (RR) use a variation of the systematic review approach, abbreviating or omitting some components to generate the evidence to inform stakeholders promptly whilst maintaining attention to bias.

*Who is this Rapid Review for?:* The review question was suggested by the Health Sciences Directorate (Policy).

*Background / Aim of Rapid Review:* There is growing demand for diagnostic services in the UK. The use of artificial intelligence in diagnosis is part of the Welsh Government’s programme for transforming and modernising planned care and reducing waiting lists in Wales. This rapid review aimed to assess the effectiveness of artificial intelligence (AI) in diagnostic radiology with a focus on cancer diagnosis. A range of AI models including machine learning, deep learning and ensemble models, were assessed in this review. The term ‘AI models’ was therefore used to encompass these different types of AI models described in the literature. The review included an initial broad mapping exercise and a more in-depth synthesis of a specific sub-set of the evidence. The focus of the in-depth synthesis was informed by the review’s stakeholders based on the findings of the mapping exercise.

*Results:* Recency of the evidence base

- The review included evidence available from 2018 until June 2023. Extent of the evidence base

- A total of 92 comparative primary studies were included in the evidence map.
- The evidence map identified 52 studies in which the AI models were in the early stages of development and validation, and highlighted breast, lung and prostate cancers as the type of cancers most frequently reported on.
- 28 studies evaluating an established model and focusing on the diagnosis of breast (n=14), lung (n=7) and prostate (n=7) cancer were included **in the in-depth synthesis.**
- Studies included in the in-depth synthesis were conducted in the USA (n=8), Japan (n=5), UK (n=2), Italy (n=2), Turkey (n=2), Germany (n=2), Netherlands (n=2), Portugal (n=1), Greece (n=1) and Norway (n=1). Two studies were conducted across multiple countries.
- All studies included in the in-depth synthesis were classified as **diagnostic accuracy studies**.
- Only one study evaluated an AI model that was commercially available in the UK.
- A total of 14 studies compared AI models to human readers or to other diagnostic methods used in practice, 13 studies compared the impact of AI on human interpretation of radiologic images when diagnosing cancer, four studies compared multiple AI models, and one study compared an inexperienced AI-assisted reader with an experienced reader without AI.
- Five studies reported on the impact of AI on diagnostic timelines (time to diagnosis, assessment time, evaluation times, and reading time).
- Four studies also reported on the impact of AI on inter/intra-reader variability, reliability, and agreement.
- **One study** reported on **clinicians’ acceptance and receptiveness** of the use of AI for cancer diagnosis. Key findings and certainty of the evidence

- Most studies reported results in favour of the AI models, however, these improvements were not always statistically significant. The studies also varied considerably in terms of AI models studied, type of cancer, images used, and comparison made; and were limited in terms of their methodology (unclear level of certainty).
- When used as a standalone diagnostic tool, there is evidence to suggest that AI can improve diagnostic accuracy or is comparable to experienced radiologists, however this may be dependent on the AI model being used (unclear level of certainty).
- There is evidence to suggest that AI may be beneficial when used as a support tool for clinicians/radiologists with less experience (unclear level of certainty).
- The impact of AI on the timeline involved in diagnosis appeared inconsistent. AI may speed up the diagnostic timeline when the level of cancer suspicion is low but may increase diagnostic timelines when the level of cancer suspicion is high (low level of certainty).
- The evidence suggests that clinicians are accepting of AI-based assistance for cancer diagnosis (low level of certainty). Research Implications and Evidence Gaps

- No study reported on any patient outcomes, including patient harms.
- No study reported on any economic outcomes.
- No study reported on equity outcomes, including equity of access.
- Further research in a real-world setting is needed to better understand the cost implications and impact on patient safety of AI for cancer diagnosis. Policy and Practice Implications

- The overall evidence for effectiveness appeared in favour of AI and several factors were identified that impact the effectiveness of the AI models.
- AI may improve diagnostic accuracy in clinicians/radiologists with less experience of interpreting radiological images.
- AI models are continually being developed and updated and findings are likely to vary between different AI models.
- Further well-designed high-quality research is needed from the UK and similar countries to better understand the effectiveness of AI in cancer diagnosis. Economic considerations

- In theory it might be possible for AI to assist with earlier diagnosis of cancer with both health and economic benefits.
- There is little evidence on the cost-effectiveness of using AI for cancer diagnosis. One modelling paper from the United States (US) suggests using AI in lung cancer screening using low-dose computerised tomography (CT) scans can be cost-effective, up to a cost of $1,240 per patient screened.
- The UK (and its constituent countries) perform consistently poorly against European and international comparators in terms of cancer survival rates. Cancer screening was suspended and routine diagnostic work deferred in the UK during the COVID-19 pandemic.
- The cost of cancer to the UK economy in 2019 was estimated to be least £1.4 billion a year in lost wages and benefits alone. When widening the perspective to include mortality, this figure rises to £7.6 billion a year. Pro-rating both figures to the Welsh economy and adjusting for inflation gives figures of £79 million and £429 million per annum respectively

## 1. BACKGROUND

### 1.1 Who is this review for?

This Rapid Review was conducted as part of the Health and Care Research Wales Evidence Centre Work Programme. The above question was suggested by the Health Sciences Directorate (Policy).

### 1.2 Background and purpose of this review

There has been a growing demand across multiple aspects of diagnostic services in the UK. This has impacted on waiting times for both diagnostics and treatment. Data from March 2023 showed that over 116,000 patients were waiting eight weeks or more for diagnostic services, of which approximately 68,000 were waiting specifically for radiology tests (Stats Wales, 2023). As part of the ‘Programme for transforming and modernising planned care and reducing waiting lists in Wales’, the Welsh Government recommended the use of Artificial Intelligence (AI) technologies to help transform diagnostic services and reduce waiting lists (Welsh Government, 2022). A range of techniques can be used to create Artificially Intelligent Systems (AIS) that are capable of carrying out health and care tasks that until now have only been able to be completed by humans (NHS, 2022). For the purposes of this rapid review, the term ‘AI model’ will encorporate any computer algorithm described within the literature that is programmed to detect cancer from a range of radiologic images.

The NHS Artificial Intelligence Laboratory (NHS AI lab) aims to incorporate AI into the health and care sector, with the goal of reducing waiting times, improving diagnosis and saving healthcare professionals’ time (NHS England, 2022). To support this the ‘AI in Health and Care Award’ was created (Department of Health and Social Care, 2021). Over three rounds of funding, the NHS AI lab have invested £123m in 86 AI technologies, including some which process images to detect cancers allowing for faster, more accurate diagnosis (Department of Health and Social Care, 2023).

With growing investment in the use of AI in diagnostic radiology, and the rapid rate of development of AI models available that could potentially be utilised by the NHS in Wales, it is important to determine if AI is effective. The puropse of this rapid review is to assess the effectiveness of AI in diagnostic radiology with a focus on cancer diagnosis. Stakeholders were interested in the following sub-questions (listed in order of priority):

– Is there any documented **harm** from use of the AI models /applications /approaches in radiology for cancer diagnosis?
– To what extent does the use of AI models /applications /approaches in radiology for cancer diagnosis improve **patient outcomes**?
– Are the AI models /applications /approaches effective in **diagnosing cancer in a real-world setting?**
– Is there evidence of the **adoption** of AI models /applications /approaches in diagnosing cancer **within the UK?**
– Are the AI models /applications /approaches described in the primary literature, licensed for use in the UK**?**
– To what extent do the AI models /applications /approaches used in radiology for the diagnosis of cancer reduce **time for completion of diagnostic testing, review, reporting** within a given clinical workflow?
– Does the evidence suggest the AI models /applications /approaches used in radiology for the diagnosis of cancer are able to be **replicated in Wales?**
– What is the **cost-effectiveness** of the AI models /applications /approaches used in radiology for the diagnosis of cancer?
– To what extent do the use of AI models /applications /approaches in radiology for cancer diagnosis reduce overall **clinician time**, reduce need for follow up, and reduce need for further intervention?
– What are the **perceptions of clinicians** with the AI models /applications/approaches used in radiology for the diagnosis of cancer?

This rapid review was conducted in two parts. Firstly, a broad mapping exercise of the existing literature on the use of AI in cancer diagnostics was conducted in order to identify and classify the available evidence. Secondly, the findings of this mapping exercise were then used to identify a focus for an in-depth synthesis of the evidence relating to the effectiveness of AI in breast, lung and prostate cancer diagnosis.

## 2. Mapping the wider evidence base

Our literature searches identified 21,403 records. This was narrowed to a total of 92 published comparative primary studies included in the mapping exercise and 21 ongoing trials. The mapping exercise sought to outline the outcome measures reported in the literature pertaining to cancer diagnostic radiology and provide details on the types of AI models assessed and the datasets utilised in the evidence base. The eligibility criteria used to select studies for the mapping exercise are outlined in Section 6.1. A reference list of all the studies included in the map can be found in appendix 1. A list containing the titles and weblinks of the 21 ongoing trials identified can be found in appendix 2.

As outlined in the evidence map presented in Table 1, 40 studies focussed on the diagnosis of breast cancer, 14 on lung cancer, 12 on prostate cancer, while 26 focussed on a range of other cancers (including gynaecological n=7, renal n=4, brain n=2, bone n=2, liposarcoma n=2, pancreatic n=1, salivary gland n=1, thyroid n=1, liver n=1, colon n=1, bowel n=1, peripheral nerve sheath n=1, soft tissue n=1, oesophageal n=1). Only diagnostic accuracy outcome measures were consistently reported across all included studies. Some studies reported ‘other’ outcomes as can be seen in the map. These included: clinician perceptions, image quality, and the impact of different manufacturers on the ability of AI to read the images. The number of images used to test the various AI models varied greatly, but generally ranged from between 101 to 500 images. The AI models identified also varied considerably. Studies were categorised in the map by those that were reporting on commercially available AI models (as stated by the publication’s study authors), those that evaluated models that had been developed previously for use in other research work, and those that included the development and validation of new models. The evidence map also sought to differentiate between studies that compered AI models with human reader comparators, and those that made comparisons between different AI models.

**Table 1.**
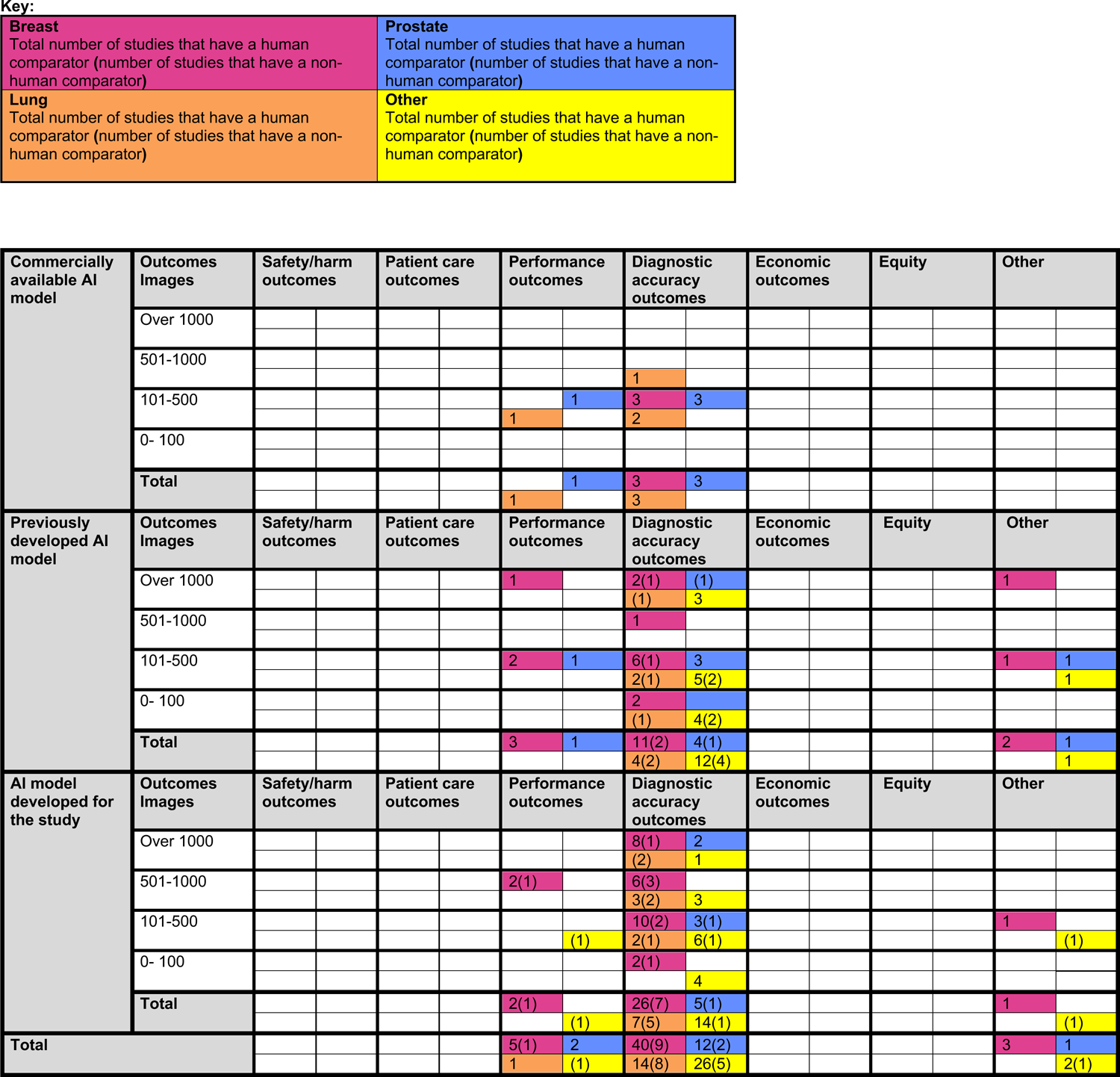
Map showing the outcome measures, stage of development of AI model and size of the overall datasets used of the evidence base (n=92).

The evidence map was presented to stakeholders in order to aid their selection of a substantive focus for a more in-depth review of the research, given the short time frame allocated for completion of this review.

## 3. Results of the in-depth synthesis

### 3.1 Overview of the evidence base

As part of the prioritisation process, stakeholders agreed that the in-depth evidence synthesis should focus on previously developed or commercially available AI models^1^ (see Table 2). Similarly, a focus on breast, lung and prostate cancers was agreed, as these were the most prevalent cancers in Wales requiring urgent action. The detailed eligibility criteria used for selecting studies for the in-depth synthesis is presented in Section 6.2 and a full summary of the included studies can be seen in Section 7.2.

**Table 2.** Name or descriptor of the artificial intelligence (AI) models utilised in the included studies.

| **Commercially available AI models** | **Previously developed AI models** |
| --- | --- |
| Quantib® - AI software | BreastScreening-AI –DNN |
| The Koios DS – DL | Xception – DLCNN |
| Prostate AI, Version Syngo.Via VB60, | InceptionV3, Inception, -DLCNN |
| Prostate AI, version 1.3.2, - DLCAD | DenseNet121, DenseNet161, - DLCNN |
| EIRL Chest X-ray Lung nodule (LPIXEL Inc) – CAD | NASNetMobile – DLCNN |
| QVCAD Qview Medical Inc, | ResNetV2, ResNet50, ResNet101, ResNet 152, - CNN |
| Red Dot, Behold.ai, -DNN | QuantX – CAD |
|  | Koios DS – DL |
|  | S-Detect – CNN |
|  | MammoScreen V1 – DCNN |
|  | Transpara. version 1.6.0 – CAD |
|  | AlexNet, - DCNN |
|  | VGG, VGG-16 – DCNN |
|  | LeNet DCNN |
|  | GoogLeNet/Inception, - CNN |
|  | Residual Networks (ResNets) – DL |
|  | AdaBoost, GBoost, XGBoost, LightGBM – Ensemble model |
|  | f-AnoGAN, HealhtyGAN, StarGAN, StarGAN-v2, FP-GAN, DeScarGAN - GAN |
|  | Grt123 – DL |
|  | JWDH- DL |
|  | Aidence – DL |
|  | CANARY* |
|  | Radiomics + Machine learning model* - ML |
|  | Random forests, back propagation neural networks (BPN), extreme learning machines, support vector machines, and K-nearest neighbors* - ML |
|  | Convolutional neural network (CNN)* |
|  | Lung Cancer Prediction CNN (LCP-CNN)* |
|  | CAD system* |

\* Only the description of type of AI model used provided

The in-depth synthesis included a total of 28 studies (breast cancer n=14, prostate cancer n=7 and lung cancer n=7). Included studies were conducted in a range of countries including USA (n=8), Japan (n=5), UK (n=2), Italy (n=2), Turkey (n=2), Germany (n=2), Netherlands (n=2), Portugal (n=1), Greece (n=1), and Norway (n=1). Two studies were conducted across multiple countries. Study designs were poorly reported across all 28 studies; however, the majority were retrospective in nature (n = 25), and most were observational. Fourteen studies explored the effectiveness of AI as an alternative method to radiologists or other conventional methods for cancer diagnosis (two of which were prospective). Thirteen studies explored the impact of AI on human interpretation of radiological images for cancer diagnosis, one of which was prospective (a total of four studies explored the effectiveness of AI as an alternative method and when assisting human interpretation). Four studies compared multiple AI models to determine the most effective models for diagnosing cancer and one study compared an inexperienced AI-assisted reader with an experienced reader without AI. All studies examined the diagnostic accuracy of AI models. Seven studies (two of which were prospective) investigated the effectiveness of commercially available AI tools while 21 studies (one of which was prospective) investigated the effectiveness of a previously developed tool.

The majority of studies relied on either existing databases of patients or datasets of images for evaluating the effectiveness of AI. Participants/images were selected from institutional databases (n=17), multiple sources (n=6), open-source datasets (n=4) and from previous studies (n=1). Most often institutional datasets were utilised for breast cancer and prostate cancer studies. The datasets used did not always originate from the country in which the study was conducted (See appendix 3). Usually, most participants or images used were those that had been flagged as having abnormal images or were confirmed as having lesions or nodules identified previously by a gold standard (often biopsy). However, the information on study population was often poorly reported, and as such it was not always clear how included patients or images were selected into the included studies.

The retrospective studies generally obtained images from historic datasets that are publicly available. The three studies that self-identified as prospective investigated the use of AI in the diagnosis of breast cancer (O’Connell et al, 2022 and Uhlig et al, 2018) and prostate cancer (Forookhi et al, 2023). The two breast cancer studies identified their populations from patients who were found to have abnormalities initially identified from ultrasound or mammography during screening or from a prior study (O’Connell et al, 2022 and Uhlig et al, 2018), the final diagnosis was already known at the time of the study, as this was used as the reference standard. However, the prostate cancer study was a truly prospective study as consecutive patients were prospectively enrolled from a cohort undergoing MRI examination for clinical suspicion of prostate cancer due to either an increase from baseline prostate-specific antigen (PSA) levels or positive digital rectal examination (DRE) findings. This study used an expert radiologist as the reference standard. Population numbers in the prospective studies tended to be small, between 35 and 299 participants and between 80 and 299 images. It should be noted that participants had already begun the diagnostic pathway and it was the initial images that were utilised in these prospective studies. Also, important to note is that study authors excluded patients on active surveillance (Forookhi et al, 2023), those with a prior diagnosis (Forookhi et al, 2023 and O’Connell et al, 2022) and in one study those who were unable to read or understand English (O’Connell et al, 2022), or those participating in a breast screening program (Uhlig et al, 2018).

Patient characteristics generally included clinical and pathological rather than demographic information. Those that did report demographic characteristics (n=23), age was the most commonly reported. This was reported in all prostate cancer studies. Six of the seven lung cancer studies also reported sex. All, but one breast cancer study (O’Connell et al, 2022) were conducted in women only. The majority of breast cancer studies included participants with lesions, although these could be normal, benign or malignant in nature. One of these (van Zelst et al, 2020) included only women with dense breasts and one study (Pacilè et al, 2020) included women with no clinical symptoms. In addition, one study (Uhlig et al, 2018) identified participants who were pre- and post-menopausal. Lastly, one breast cancer study (Lo Gullo et al, 2020) included only BRCA 1 or BRCA 2 mutation carriers. Two studies (O’Connell et al, 2022 and Maldonado et al, 2021) included ethnicity, of which Caucasians predominated. Only one lung cancer study (Maldonado et al, 2021) reported participant smoking status. Generally, participants with a prior history of or those under active surveillance or treatment for the specific cancer of interest were excluded from studies.

The included studies utilised a range of diagnostic imaging techniques including MRI (n=12), CT scans (n=6), ultrasound (n=4), X-rays (n=2), mammograms (n=2), and digital breast tomosynthesis (DBT)(n=1). One study included a combination of mammograms, ultrasound and MRI. Outcome measures reported included: the impact of AI on diagnostic accuracy, inter/intra-variability/agreement, time to diagnosis, assessment time, evaluation time, reading time, and clinicians’ acceptance and receptiveness of the use of AI for cancer diagnosis.

The type of AI models used within the studies were not always clearly described. The studies included deep learning (DL) models (n=16), machine learning (ML) models (n=5), ‘AI software’ n=2, ensemble learning models (n=1), Generative Adversarial Networks (GAN) (n=1), convolutional neural networks (CNN) (n=1), Computer aided diagnosis software (CAD) (n=1), and Computer-Aided Nodule Assessment and Risk Yield (CANARY) (n=1). Within the studies using a DL model, nine were reported to be deep learning convolutional neural networks (DLCNNs), three were deep learning computer aided diagnosis software (DLCADs) and four were just described as DL models).

The AI models evaluated in the included studies were at different stages of development. Seven studies explored the use of commercially available AI models. It should be noted that the commercially available models were licenced for use in different countries, however, this was not always clearly stated in the studies. Only one AI model was commercially available within the UK (Red Dot, Behold.ai). The remaining 21 studies explored the use of previously developed AI models. The majority of AI models that were included in this in-depth synthesis were specifically named as shown in Table 2. However, some AI models were only given descriptors rather than a specific name, with the descriptors outlining details about the type of AI model used (e.g. radiomics, prediction CNN, CAD system). Full details about the AI models used in each study can be seen in section 7.2.

The methodological quality of included studies was assessed using the QUADAS-2 (Whiting et al, 2011) and QUADAS-C (Yang et al, 2021) tools. Quality appraisal identified three studies to be at low risk of bias (Lo Gullo et al, 2020, Maldonado et al, 2021, Tong et al, 2023), while the remaining studies had methodological limitations and were therefore judged to be at high or unclear risk of bias. Common methodological limitations across studies included poor reporting of patient/image selection. Studies also often failed to adequately describe how images were distributed among the intervention and control groups. In addition, several studies excluded images that were considered of poor quality or images containing several or complex lesions which could have limited the generalisability of the findings. Two of the three prospective studies were determined to have an unclear risk of bias due to missing details around how the comparators were conducted (Uhlig et al 2018, and Forookhi et al 2023) and one was determined to have a high risk of bias due to some participants being removed from the analysis (O’Connell et al 2022).

A clear description of how the index test was conducted and interpreted was lacking among some studies. Many studies utilised the diagnosis reported in the database from where the images were taken as the final diagnosis. Other studies did interpret images independently, or in the case of images collected at local hospitals, used the final diagnosis as the reference standard. However, in the case of AI being compared against human readers, the timing of the index test and reference standard was unclear, which could have introduced bias. Further details of the quality appraisal can be found in section 7.3.

### 3.2 Impact of AI on diagnostic accuracy

All studies reported on the impact of AI on diagnostic accuracy, the findings of which are summarised in Table 3. When assessing the diagnostic accuracy of a test, multiple measurements can be reported, which include but are not limited to: sensitivity and specificity, positive and negative predictive values (PPV, NPV), and the area under the receiver operating characteristic curve (ROC AUC, often reported as AUC) (Šimundić 2009). The sensitivity of a diagnostic test measures the proportion of true positives identified by the test, whereas the specificity of a diagnostic test represents the proportion of true negatives identified (Wong and Lim 2011). PPV represents the likelihood that a patient with a positive test actually has the disease and the NPV represents the likelihood that a patient with a negative test does not have the disease (Safari et al 2015). The AUC represents how well the diagnostic test can discriminate, in this case between cancer and non-cancer, an AUC of 1 would be a perfect diagnostic test whereas a non-discriminative test would give an AUC of 0.5 (Eusebi 2013).

**Table 3:**
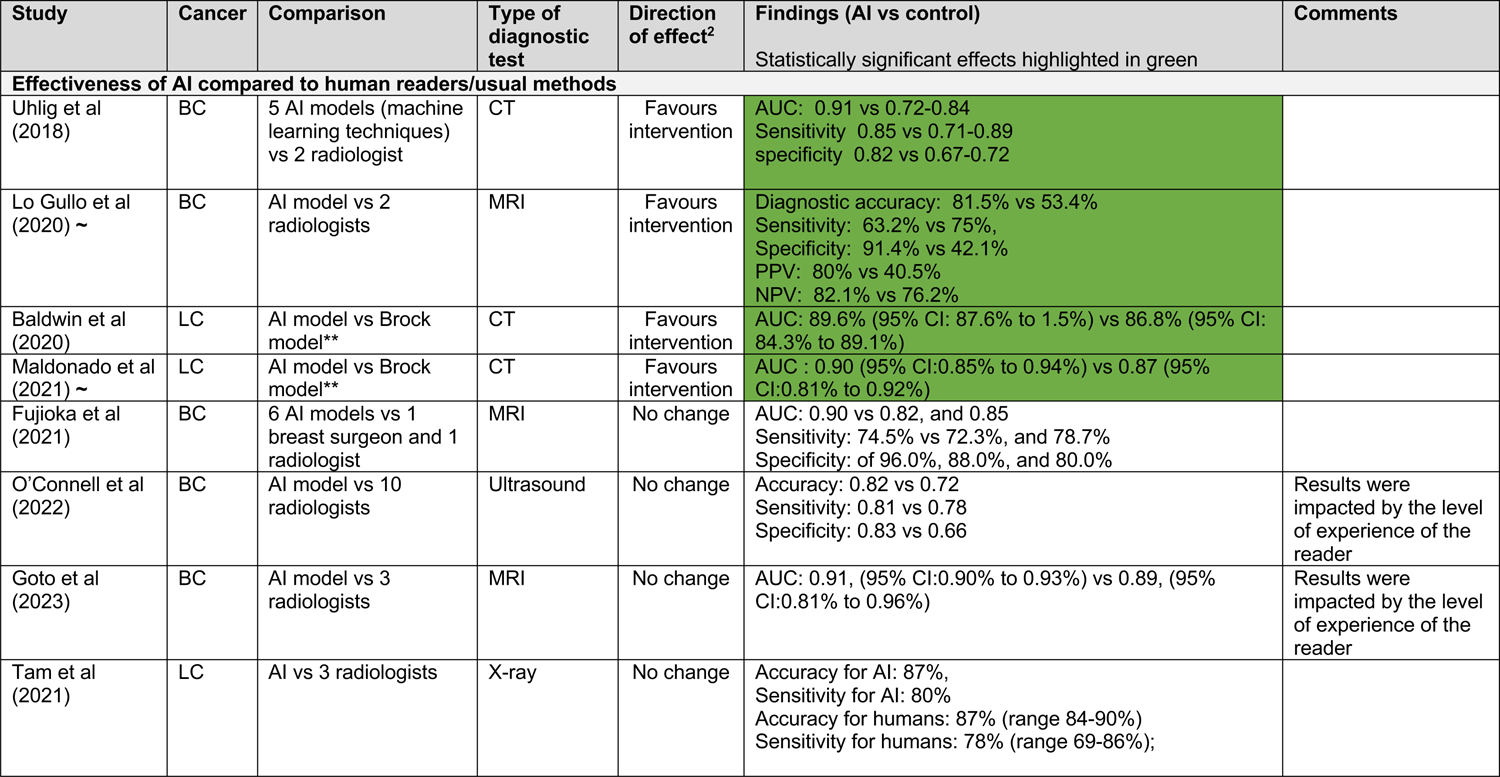
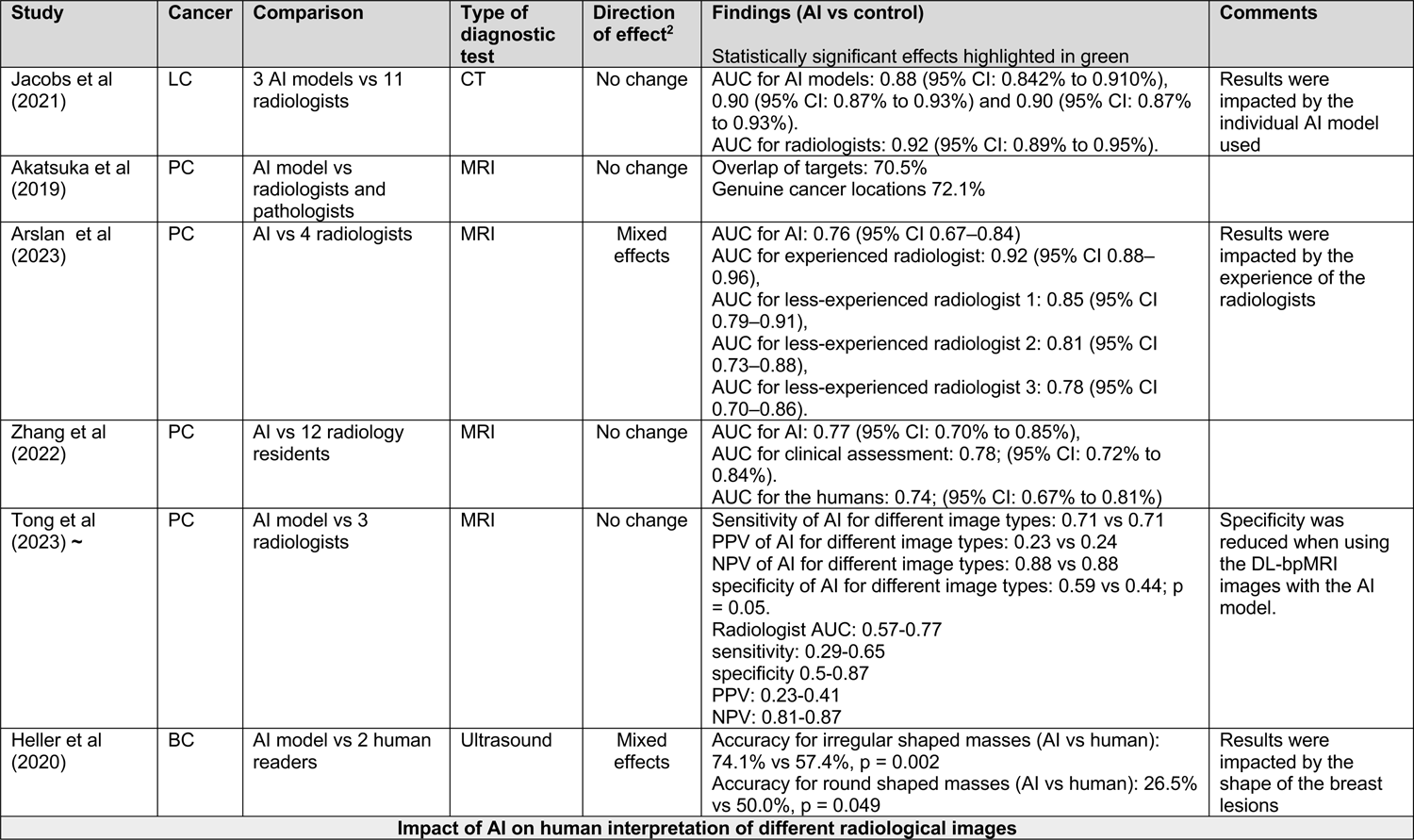
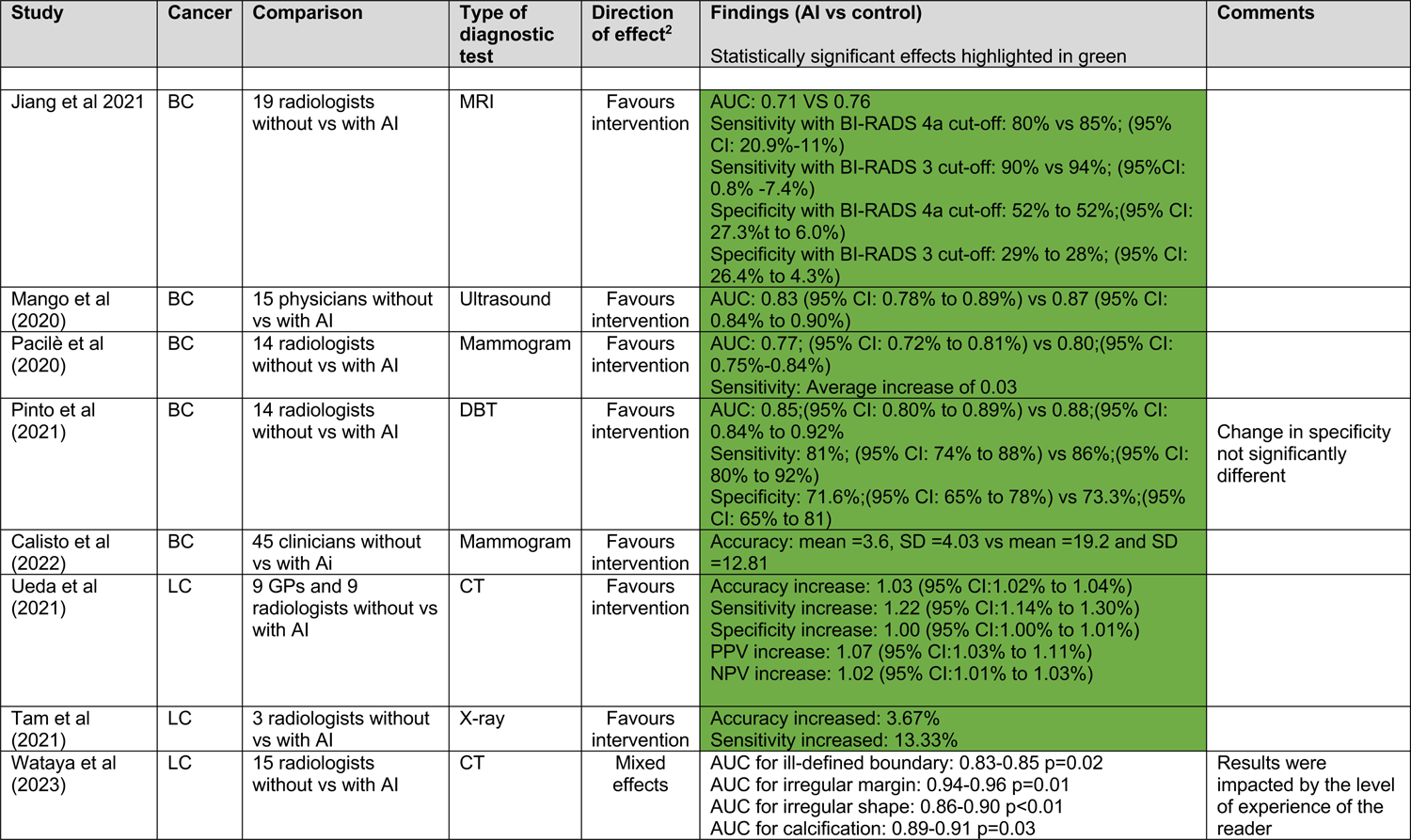
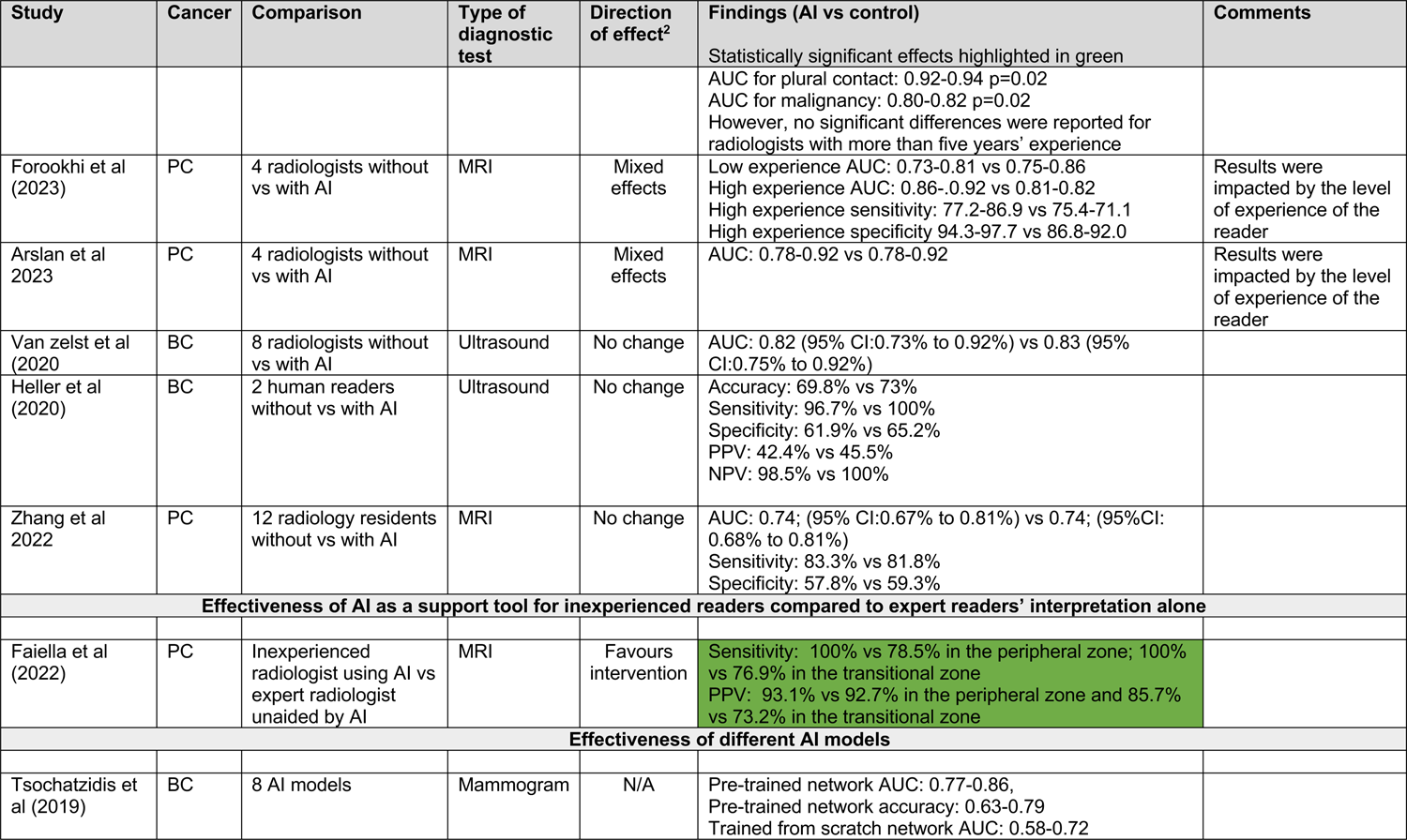
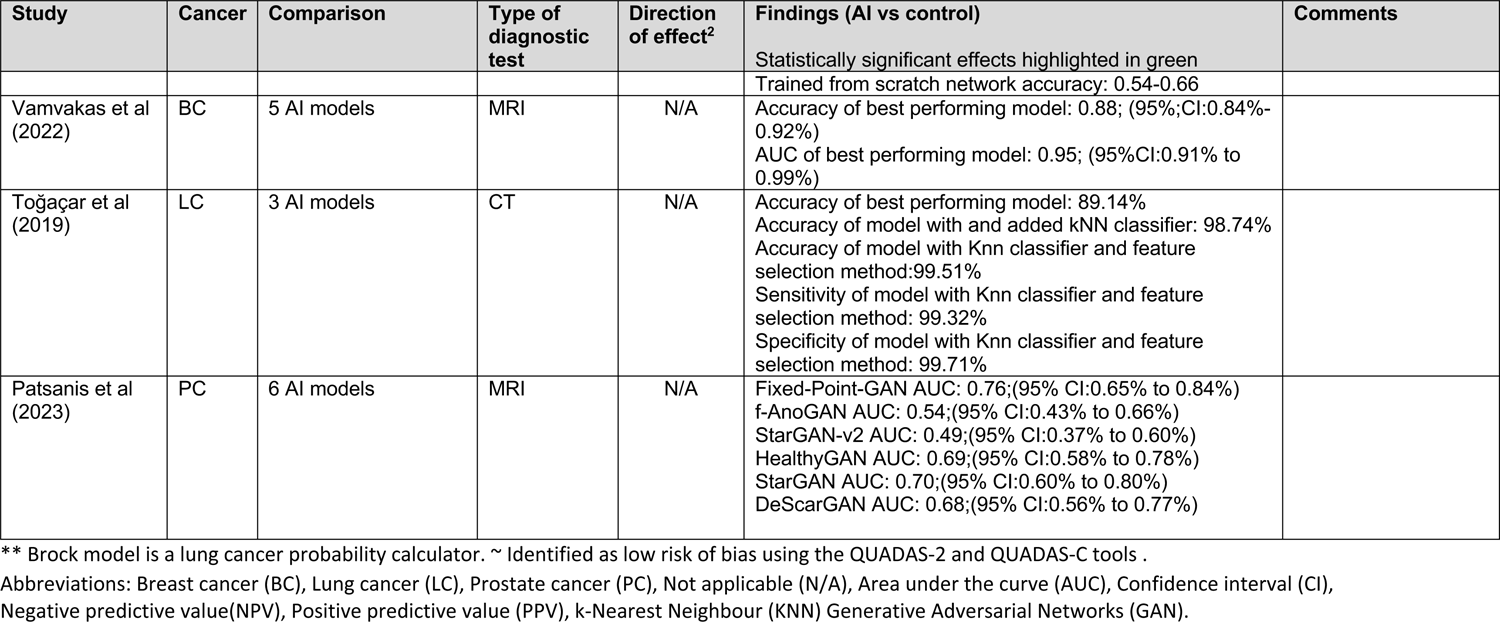
Summary of the findings for the impact of artificial intelligence (AI) on diagnostic accuracy.

The results for diagnostic accuracy are grouped according to the comparisons made into the following categories:

- AI compared to human readers/usual methods
- AI assisting human interpretation
- Comparison of different AI models

(individual studies may have multiple aims and are therefore reported under more than one category).

As the AI models, comparators and datasets varied widely across studies, each study has been narratively reported separately. It was not always appropriate to combine findings. Where this has been done, it should be highlighted that the studies may have used different imaging techniques for the diagnosis of different types of cancer.

### Effectiveness of AI compared to human readers/conventional methods

A total of 14 studies assessed the effectiveness of AI models in detecting cancer compared to human readers (radiologists/clinicians) or other conventional diagnostic methods (e.g., the Brock model, a lung cancer probability calculator). The findings were inconsistent.

Four studies (breast n=2, lung n=2) found evidence that the use of AI may improve cancer diagnosis (Uhlig et al, 2018, Lo Gullo et al, 2020, Baldwin et al, 2020, Maldonado et al, 2021). Three studies used CT images and one study used MRI scans.

Uhlig et al (2018) compared the diagnostic performance of five machine learning techniques (random forests, back propagation neural networks [BPNs], extreme learning machines, support vector machines, and K-nearest neighbours) with that of two independent human readers (radiologists) for the diagnosis of breast cancer from Cone-beam Computed Tomography (CBCT) images. BPNs were found to be the highest performing model and also **performed better than the human readers** (0.91 AUC, 0.85 sensitivity, 0.82 specificity for the BPNs vs 0.72 to 0.84 AUC, 0.71 to 0.89 sensitivity, 0.67 to 0.72 specificity for human readers). **The AUC was statistically significantly higher for BPNs when compared with both human reader 1 (p = 0.01) and human reader 2 (p < 0.001)**.

Lo Gullo et al (2020) investigated the diagnostic accuracy of radiomic analysis and ML in differentiating between benign and malignant breast lesions from MRIs compared to independent assessments by two human readers (radiologists). The findings showed **an improved diagnostic performance by the machine learning model compared to the radiologists** (diagnostic accuracy 81.5% vs 53.4%, sensitivity 63.2% vs 75%, specificity 91.4% vs 42.1%, PPV 80% vs 40.5%, NPV 82.1% vs 76.2%; respectively).

Baldwin et al (2020) compared the effectiveness of an AI model (LCP-CNN) to the Brock model for the diagnosis of lung cancer from CT scans. This study found that **the AI model was statistically significantly better at predicting the risk of malignancy compared to the Brock model**. The AUC for the AI model was 89.6% (95% CI: 87.6% to 1.5%), compared with 86.8% (95% CI: 84.3% to 89.1%) for the Brock model (p≤ 0.005).

Maldonado et al (2021) assessed the effectiveness of the BRODERS radiomic predictive model in predicting the probability of malignancy in an independent dataset of incidentally detected indeterminate pulmonary nodules by comparing its performance to that of the Brock model on CT scans. The findings showed **a significantly greater AUC for the BRODERS model compared to the Brock model at all pre-test malignancy probabilities** 0.90 (95% CI:0.85% to 0.94%) vs 0.87 (95% CI:0.81% to 0.92%) (p<0.001).

Eight studies (breast n=3, lung n=2, and prostate n=3) found evidence to suggest the use of AI led to no significant difference between groups or reported similar outcomes when compared to human readers (Fujioka et al, 2021, O’Connell et al, 2022, Goto et al, 2023, Tam et al, 2021, Jacobs et al, 2021, Akatsuka et al, 2019, Arslan et al, 2023, Zhang et al, 2022). However, two of these studies found that the level of experience of radiologists impacted whether the use of AI improved accuracy of diagnosis (O’Connell et al, 2022, Goto et al, 2023). These studies used CT (n= 1), MRI (n= 5) and ultrasound (n= 1) and X-rays (n=1).

Fujioka et al (2021) evaluated the effectiveness of six CNN models in discriminating between benign and malignant breast lesions on MRI by comparing their performance with that of two human readers (a breast surgeon and a radiologist). **The findings showed no significant differences between the CNN models** (DenseNet121, DenseNet169, InceptionResNetV2, InceptionV3, NasNetMobile, and Xception) **and the human readers**. The mean AUC of all AI models was 0.83 (range 0.75 to 0.90). The best performing AI model was InceptionResNetV2, however no statistically significant differences were reported when compared with the two human readers (AUC 0.90 vs 0.82, and 0.85; sensitivity 74.5% vs 72.3%, and 78.7%; and specificities of 96.0%, 88.0%, and 80.0%, respectively [p > 0.125]).

O’Connell et al (2022) studied the performance of an AI model (S-Detect™) in the diagnosis of breast cancer in ultrasound images by comparing this model to manual readings by 10 human readers (radiologists) with varying levels of experience. **The AI model was found to have similar levels of accuracy, sensitivity and specificity compared to the experienced radiologists** (accuracy 0.82 vs 0.72, sensitivity 0.81 vs 0.79, specificity 0.83 vs 0.67, respectively).

Goto et al (2023) compared an AI model (ResNet50) to three human readers (radiologists with varying levels of experience) when determining malignancy from breast MRI. **When precise segmentation was conducted the AI model achieved similar levels of accuracy compared to a highly experienced radiologist** (AUC =0.91, 95% CI:0.90% to 0.93% vs AUC =0.89, 95% CI:0.81% to 0.96%; p=0.45, respectively). When rough segmentation was conducted, the AI model showed similar levels of accuracy to a board-certified radiologist (AUC 0.80, 95% CI:0.78% to 0.82% vs. AUC 0.79, 95% CI:0.70% to 0.89%, respectively). However, regardless of segmentation method, the AI model was found to be significantly more accurate than a radiology resident (AUC =0.64,95% CI: 0.52% to 0.76%; p=0.01).

Tam et al (2021) evaluated the use of a commercially available AI model (Red Dot, Behold.ai) for the diagnosis of lung cancer from X-rays compared to three human readers (radiologists) and in combination with the readers. **The average accuracy and sensitivity for the three human readers alone was 87%** (range 84 to 90%) and 78% (range 69 to 86%); respectively, **which was similar to when the AI model was used alone** (accuracy 0.87%, sensitivity 0.8).

Jacobs et al (2021) compared the performance of three top-performing AI algorithms (grt123, JWDH, and Aidence) to that of 11 human readers (radiologists) in their ability to identify lung cancer from low-dose CT scans. The AUC values were 0.88 (95% CI: 0.84% to 0.91%) for grt123 algorithm, 0.90 (95% CI: 0.87% to 0.93%) for Aidence algorithm, and 0.90 (95% CI: 0.87% to 0.93%) for JWDH algorithm. For the radiologists, the AUCs ranged from 0.841 (95% CI: 0.80% to 0.88%) to 0.94 (95% CI: 0.92% to 0.96%), with an average AUC of 0.92 (95% CI: 0.89% to 0.95%). **The grt123 AI algorithm performed statistically significantly better compared to radiologists** (p = 0.02) **but no differences were found between the other models and radiologists** (JWDH, p = 0.29; and Aidence, p = 0.26).

Akatsuka et al (2019) assessed whether a DeepCNN AI model (Xception) could correctly locate prostate cancer on MRIs compared to human readers (radiologists and pathologists). **The AI model overlapped the reader-identified targets in a statistically significant similar number of the MRI images** (70.5% p < 0.001) and was found to focus on a statistically significant number of genuine cancer locations (72.1% p<0.001).

Arslan et al (2023) compared the diagnostic performance of four human readers (radiologists with different levels of experience), with and without the use of a commercially available deep learning AI model (Prostate AI, Version Syngo.Via VB60) for the diagnosis of prostate cancer using bi-parametric MRI images. **The AUCs of the experienced radiologist and one of the less-experienced radiologists were statistically significantly higher than the AI model on its own** (AUC 0.92; 95% CI:0.88% to 0.96% and 0.85; 95% CI: 0.79% to 0.91% vs 0.76; 95% CI:0.67% to 0.84%; p< 0.0001 and p= 0.04; respectively). **However, no significant differences were reported for the other less-experienced radiologists** (p = 0.63 and p = 0.23 respectively).

Zhang et al (2022) assessed the effectiveness of 12 human readers (radiology residents) when using a deep learning CNN AI model for the diagnosis of prostate cancer from MRIs. **The AUC of the AI model alone was 0.77 (95% CI: 0.70% to 0.85%), which was similar to clinical assessment 0.78; (95% CI: 0.72% to 0.84%). The AUC for the human readers was also similar** 0.74; (95% CI: 0.67% to 0.81%) with no statistically significant differences reported. One study evaluated the impact of different image types on the diagnostic performance of readers and an AI model in diagnosing prostate cancer.

Tong et al (2023) assessed the impact of using conventional Biparametric MRI (CL-bpMRI) and deep learning accelerated Biparametric MRI (DL-bpMRI) images in both a human reader study (three radiologists) and a study using a commercially available deep learning-based computer-assisted detection (DL-CAD) AI model. When using the AI model to assess CL-bpMRI and DL-bpMRI images no differences were reported between sensitivity (0.71 vs 0.71), PPV (0.23 vs 0.24), or NPV (0.88 vs 0.88). However, a statistically significant reduction in specificity when using DL-bpMRI compared to CL-bpMRI was found (0.59 vs 0.44; p = 0.05). **No statistically significant differences were reported between the human readers for the different image types.** The AUC of the human readers ranged from 0.57 to 0.77, sensitivity 0.29 to 0.65, specificity 0.5 to0.87, PPV 0.23 to0.41 and NPV 0.81 to 0.87. One study evaluated the impact of AI on diagnostic accuracy when looking at lesion shape and found mixed results.

Heller et al (2020) assessed the effects of a commercially available deep learning AI support system (Koios DS) for the diagnosis of breast cancer from ultrasound images. The AI model was statistically significantly more accurate than human readers for irregular shaped masses (74.1% vs 57.4%, p = 0.002) and significantly less accurate for round shaped masses (26.5% vs 50.0%, p = 0.049).

### Effectiveness of AI plus human interpretation of radiological images

A total of 13 studies assessed the effect of AI assisted human interpretation of images when diagnosing cancer. Seven studies (breast n=5, lung n=2) reported a positive effect of using AI plus human interpretation on diagnostic accuracy (Jiang et al, 2021, Mango et al, 2020, Pacilè et al, 2020, Pinto et al, 2021, Calisto et al, 2022, Ueda et al, 2021, Tam et al, 2021). These studies used MRI (n=1), ultrasound (n=1), mammograms (n=1), DBT (n=1), CT (n=1), X-ray (n=1), and one study used mammograms, ultrasounds and MRIs (n=1).

Jiang et al 2021 compared the diagnostic performance of 19 human readers (radiologists) with and without the use of an AI model (QuantX) for the diagnosis of breast cancer from dynamic contrast material–enhanced (DCE) MRI**. The average AUC of the human readers significantly improved when using the AI system** (0.71 to 0.76 p = 0.04). Sensitivity improved when BI-RADS category 3 was used as the cut-off point (90% to 94%; 95% CI: 0.8% to 7.4%) but not when using BI-RADS category 4a (80% to 85%; 95% CI: 20.9% to 11%). Specificity showed no difference with either BI-RADS category 4a or category 3 (from 52% to 52%;(95% CI: 27.3% to 6.0%), and from 29% to 28%; (95% CI: 26.4% to 4.3%), respectively, no p value reported). Mango et al (2020) assessed the effects of a deep learning AI support system (Koios DS) for the diagnosis of breast cancer from ultrasound images in 15 human readers (physicians).

**The mean AUC for the human readers statistically significantly improved from 0.83 (95% CI: 0.78% to 0.89%) to 0.87 (95% CI: 0.84% to 0.90%) when using the AI model** (p<0.0001) compared to human readers alone.

Pacilè et al (2020) assessed the effects of an AI model (MammoScreen V1) for the diagnosis of breast cancer from mammograms. **The average AUC across the 14 included human readers (radiologists) significantly improved when using the AI model** from 0.77 (95% CI: 0.72% to 0.81%), to 0.80 (95% CI: 0.75% to 0.84%); average difference 0.03 (95% CI:0.00% to 0.06%; p = 0.035). Sensitivity was also found to significantly improved when using the AI model (average increase of 0.03; p=0.21).

Pinto et al (2021) compared the use of an AI model (Transpara. V1.6.0) for the diagnosis of breast cancer from DBT images with 14 human readers (radiologists). **The average AUC for the 14 human readers was statistically significantly higher when interpreting results with AI** (0.88;95% CI: 0.84% to 0.92% vs 0.85;95% CI: 0.80% to 0.89%, respectively; p = 0.01). The average sensitivity also significantly increased with the use of AI from 81% (95% CI: 74% to 88%) to 86% (95% CI: 80% to 92%) p= 0.006), whereas no differences were found in the specificity (71.6%; 95% CI: 65% to 78%; vs 73.3%; 95% CI: 65% to 81%; p = 0.48).

Calisto et al (2022) assessed the use of a deep neural network (DNN) AI model (DenseNet) for the diagnosis of breast cancer from mammograms, ultrasound and MRIs. **Diagnostic accuracy was higher when the 45 human readers (clinicians) used the AI model** (mean =19.20 and SD =12.81) compared to human reader alone (mean =3.60, SD =4.03). The mean and standard deviation for precision and recall with and without using the AI model was similar, (M = 0.66, SD = 0.34 and M = 0.62, SD = 0.27, respectively). However, it was unclear if these differences were statistically significant.

Ueda et al (2021) compared the diagnostic performance of 18 human readers (nine GPs and nine radiologists with different levels of experience), with and without the use of a commercially available deep learning AI model (EIRL) for the diagnosis of lung cancer using CT images. **All human readers significantly improved accuracy, sensitivity, PPV and NPV when using the AI model** (p<0.001, p<0.001, p=0.002, p<0.001 respectively). The overall increases for sensitivity were 1.22 (95% CI:1.14% to 1.30%), specificity 1.00 (95% CI:1.00% to 1.01%), accuracy 1.03 (95% CI:1.02% to 1.04%), PPV 1.07 (95% CI:1.03% to 1.11%), and NPV 1.02 (95% CI:1.01% to 1.03%).

Tam et al (2021) evaluated the use of a commercially available AI model (Red Dot, Behold.ai) for the diagnosis of lung cancer from X-rays compared to three human readers (radiologists) and in combination with the readers. **The overall accuracy and sensitivity was significantly increased when human readers used the AI model, improving average scores by 3.67% and 13.33%, respectively, p<0.05).**

Three studies (lung n=1, prostate n=2) found that the benefits of using different AI models to assist human readers were impacted by the level of experience of the reader themselves (Wataya et al, 2023, Forookhi et al, 2023, Arslan et al, 2023). These studies used MRI (n=2) and CT images (n=1).

Wataya et al (2023) compared the performance of 15 human readers (radiologists with varying levels of experience) with and without the use of a deep learning AI model (CAD) for the diagnosis of lung cancer from CT images. For all radiologists, significant improvements were found when using the AI model for lesions with an ill-defined boundary (AUC from 0.83 to 0.85 p=0.02), irregular margin (AUC from 0.95 to 0.97 p=0.01), irregular shape (AUC from 0.86 to 0.91 p<0.01), as well as calcification (AUC from 0.89 to 0.91 p=0.03), plural contact (AUC from 0.92 to 0.94 p=0.02) and malignancy (AUC from 0.80 to 0.82 p=0.02). **However, no significant differences were reported in the group of radiologists with more than five years’ experience before and after using the AI model.**

Forookhi et al (2023) compared the diagnostic accuracy of four human readers (radiologists with different levels of experience), with and without the use of a commercially available AI model (Quantib®) for the diagnosis of prostate cancer using mpMRI images. **For less experienced human readers, the AI model improved diagnostic accuracy,** AUC ranges rose when using the AI model (from 0.73-0.81 to 0.75-0.86). However, **more experienced human readers performed better without the use of the AI model** (AUC 0.86; 95% CI:0.81% to 0.91%, sensitivity 77.2%, specificity 94.3% and AUC 0.92; 95% CI:0.89% to 0.96%, sensitivity 86.9, specificity 97.7 to AUC 0.81[95% CI:0.76% to 0.86%, sensitivity 75.4, specificity 86.8 and AUC 0.82 95% CI:0.76% to 0.87%, sensitivity 71.1, specificity 92.0; respectively).

Arslan et al (2023) compared the diagnostic performance of four human readers (radiologists with different levels of experience), with and without the use of a commercially available deep learning AI model (Prostate AI, Version Syngo.Via VB60) for the diagnosis of prostate cancer using bi-parametric MRI images. **The AUCs of radiologists with and without the AI model did not differ overall** (AUC ranged from 0.78 to 0.92 without AI to AUC 0.78 to 0.92 with AI p > 0.05).

Three studies (breast n=2, prostate n=1) found the use of AI made no statistically significant difference to the diagnostic performance of the human readers (Van Zelst et al, 2020, Heller et al, 2020, Zhang et al, 2022). These studies used ultrasound (n=2) and MRIs (n=1).

Van Zelst et al (2020) assessed the effectiveness of eight human readers (radiologists) when using a commercially available AI model (QVCAD) for the diagnosis of breast cancer from ultrasounds**. The overall difference in AUC was not statistically significant before 0.82 (95% CI: 0.73% to 0.92%) and after 0.83 (95% CI: 0.75% to 0.92%) using the AI model (p= 0.74).** However partial AUC improved significantly from 0.13 (95% CI:0.10 to 0.15) to 0.14 (95% CI: 0.12% to 0.17%) (p=0.04) after the AI model was used.

Heller et al (2020) assessed the effects of a commercially available deep learning AI support system (Koios DS) for the diagnosis of breast cancer from ultrasound images. **No statistically significant differences were found in accuracy (69.8% vs 73%) NPV (98.5% vs 100%), PPV (42.4% vs 45.5%), sensitivity (96.7% vs 100%), and specificity (61.9% vs 65.2%; p= 0.12–0.41) before and after two human readers (with breast imaging experience) used the AI model.** The AI model also significantly improved diagnostic accuracy for human reader-rated low-confidence lesions with increased PPV (24.7% AI vs 19.3%, p = 0.004) and specificity (57.8% vs 44.6%, p = 0.008).

Zhang et al (2022) assessed the effectiveness of 12 human readers (radiology residents) when using a deep learning CNN AI model for the diagnosis of prostate cancer from MRIs. Overall **radiology residents achieved similar sensitivity and specificity before and after using the AI model** (83.3% and 57.8% vs 81.8% and 59.3%; p=1.0. The AUC for the human readers was also similar 0.74; (95% CI: pre, 0.67% to 0.81%; post, 0.68% to 0.81%) with no statistically significant differences reported.

### Effectiveness of AI as a support tool for inexperienced readers compared to expert readers’ interpretation alone

While the above studies compared the diagnostic accuracy of readers before and after being assisted by an AI model, only one study assessed the impact of AI on less experienced readers compared to expert readers without the use of AI.

Faiella et al (2022) evaluated the clinical utility of an AI model (Quantib Prostate) for prostate cancer detection on Multiparametric MRI (mpMRIs) by comparing its diagnostic performance when used by human readers with differing levels of experience (an inexperienced radiologist using the AI model and an expert radiologist not aided by the AI model). **The AI-assisted radiologist had a sensitivity of 100% in both zones and a PPV of 93.1% in the peripheral zone and 85.7% in the transitional zone. Whereas the expert radiologist had a sensitivity of 78.5% in the peripheral zone and 76.9% in the transitional zone and a PPV of 92.7% in the peripheral zone and 73.2% in the transitional zone.** However, it was unclear if these differences were statistically significant.

### Effectiveness of different AI models

A total of four studies (breast n=2, lung n=1, prostate n=1) compared the diagnostic accuracy of a range of AI models (Tsochatzidis et al, 2019, Vamvakas et al, 2022, Toğaçar et al, 2019, Patsanis et al, 2023). Findings identified factors that could potentially improve diagnosis when using AI. These studies used MRI (n=2), mammograms (n=1), and CT images (n=1).

Tsochatzidis et al (2019) compared the diagnostic accuracy of eight CNN AI models (AlexNet, VGG-16, VGG-19, ResNet-50, ResNet-101, ResNet-152, GoogLeNet and Inception-BN (V2)) either trained from scratch or pre-trained and fine-tuned for the diagnosis of breast cancer using mammograms obtained from two datasets. The highest performing models were the fine-tuned ResNet-50 and ResNet-101 models in both datasets (AUC 0.86 and 0.80 vs 0.86 and 0.79; accuracy 0.74 and 0.75 vs 0.79 and 0.75, respectively). Fine-tuning a pre-trained network improved accuracy compared to training from scratch (AUC ranged from 0.77 to 0.86, accuracy ranged from 0.63 to 0.79 for pre-trained networks compared to AUC 0.58 to 0.72 and accuracy 0.54 to 0.66; respectively). **However, it was unclear if these differences were statistically significant.**

Vamvakas et al (2022) evaluated the use of ensemble classification AI models for the diagnosis of breast cancer utilising mpMRI. AI models (XGboost, LGBM, Adaboost and GB) were compared to a support vector machine (SVM). **XGboost achieved the highest accuracy and overall performance** (accuracy 0.88; 95% CI: 0.84% to 0.92%, AUC 0.95; 95% CI: 0.91% to 0.99%), followed by LGBM (accuracy 0.87; 95% CI: 0.83% to 0.91%, AUC 0.94; 95% CI: 0.90% to 0.98%). XGBoost also achieved the highest sensitivity (0.91; 95% CI: 0.85% to 0.97%) and specificity (0.90; 95% CI: 0.82% to 0.98%) compared to the other models. The SVM had a statistically significantly lower performance (accuracy 0.84; 95% CI: 0.80% to 0.88%, AUC 0.88; 95% CI: 0.84% to 0.92%) than XGBoost and LGBM but was found to be statistically comparable with the performances demonstrated by AdaBoost and GB.

Toğaçar et al (2019) compared the effectiveness of multiple AI models (LeNet, AlexNet and VGG-16) for lung cancer diagnosis using CT images and found the AlexNet(SGD-Drop(0,5) was the best performing model (accuracy 89.14%). However, using a combination of the AlexNet model and a k -nearest neighbour (kNN) classifier improved the accuracy of the model by 98.74%. Finally, when **adding a minimum redundancy maximum relevance (mRMR) feature selection method to the model along with the kNN classifier the accuracy increased further** (99.51%, sensitivity 99.32% and specificity 99.71%).

Patsanis et al (2023) assessed six previously developed deep learning GANs for the diagnosis of prostate cancer from MRIs. Six GANs (f-AnoGAN, HealthyGAN, StarGAN, StarGAN-v2, Fixed-Point-GAN and DeScarGAN) were evaluated using a validation data set. Fixed-Point-GAN performed significantly better (AUC 0.76; 95% CI: 0.65% to 0.84%) than f-AnoGAN and StarGAN-v2 (AUC 0.54; 95% CI: 0.43% to 0.66%, vs AUC 0.49; 95% CI: 0.37% to 0.60%; respectively) but not compared to HealthyGAN, StarGAN, and DeScarGAN (AUC 0.69; 95% CI: 0.58% to 0.78%, AUC 0.70; 95% CI: 0.60% to 0.80%, AUC 0.68; 95% CI: 0.56% to 0.77% respectively).

#### 3.2.2 Bottom line results for the impact of AI on diagnostic accuracy

The use of AI for the diagnosis of cancer shows inconsistent findings. There is evidence to suggest that AI alone may be used to improve the accuracy of cancer diagnosis, but that this is dependent on the specific AI model being used. There is also evidence to show that the use of AI models to support the accuracy of cancer diagnosis using different radiological techniques may be more beneficial to less experienced human readers, and less helpful to more experienced human readers. There is very limited evidence to indicate the beneficial use of AI when interpreting irregular shaped lesions.

The evidence suggests that pre-training the AI model may improve its diagnostic performance. Ensemble models (a combination of multiple AI models) may be more effective and additional classifiers can be added to further increase the diagnostic performance of an AI model. However, these outcomes were only reported by individual studies and as such firm conclusions cannot be made.

### 3.3 Impact of AI on inter and intra-reader variability, reliability, agreement

Four studies (breast n=3, prostate n=1) reported on inter/intra-variability or agreement before and after readers used AI (Calisto et al, 2022, Pacilè et al, 2020, Mango et al, 2020, Forookhi, et al 2023). These studies used mammograms (n=1), ultrasounds (n=1) and MRIs (n=1), with one study using mammograms, ultrasounds and MRIs (n=1). Inter reliability/variability or agreement is the agreement or differences in diagnosis between the individual human readers and intra reliability/variability or agreement assesses the differences reported by the same reader e.g. if the diagnosis would differ after the reader used AI. All studies reported some improvement in accuracy when using AI for cancer diagnosis.

Calisto et al (2022) assessed the use of a DNN AI model (DenseNet) for the diagnosis of breast cancer from mammograms, ultrasound and MRIs. It was found that the **inter-variability of the diagnosis made by the 45 human readers (clinicians) improved when using the AI model** on patients with low, medium and high severities (depending on BI-RAD classification) (11%, 3.28%, 34.1% respectively). When looking at **the intra-variability, all human readers also improved** their results with the introduction of AI, those with little experience improved by 6.65%, those with 6 to 10yrs experience improved by 23.66%, those with 11 to 30yrs experience improved by 14.58% and those with 31 to 40 yrs improved by 19.64%. **However, it was unclear if these differences were statistically significant.**

Pacilè et al (2020) assessed the effects of an AI model (MammoScreen V1) for the diagnosis of breast cancer from mammograms and reported that the inter reader reliability among the 14 human readers (radiologists) appeared to increase when using AI. A moderate inter-rater reliability was found in both reading conditions. **For the unaided reading condition, the inter-rater agreement between the human readers was 0.59 (95% CI: 0.53% to 0.64%), while for the reading with AI inter-rater agreement increased to 0.68 (95% CI: 0.62% to 0.73%). However, it was unclear if these differences were statistically significant.** Mango et al (2020) assessed the effects of a deep learning AI support system (Koios DS) for the diagnosis of breast cancer from ultrasound images on 15 human readers (physicians).

Inter-reader variability without AI was 0.54 (95% CI: 0.53% to 0.55%; compared to 0.68 (95% CI: 0.67% to 0.69%) with the AI. **Intra-reader variability improved with AI, showing a statistically significant difference** (α = 0.05). Intra-reader variability resulted in less class switching (e.g. from lower than BI-RADS 4A to BI-RADS 4A or higher) with AI than without overall. Although a statistically significant trend toward lower intra reader variability with AI was reported. The class switching rate without AI was 13.6%, and with AI was 10.8% (p = 0.04). Nine readers showed decreased class switching with AI, one reader showed equivalent class switching and five showed more class switching with AI. **The findings indicate that the overall improvement in intra reader variability did not extend to all readers.**

Forookhi et al (2023) compared the diagnostic accuracy of four human readers (radiologists with different levels of experience), with and without the use of a commercially available AI model (Quantib®) for the diagnosis of prostate cancer using mpMRI images. The results showed that the inter-reader agreements at different PI-QUAL scores were higher with the use of the AI model, particularly for less experienced human readers, showing a moderate to slight agreement. **However, it was unclear if the differences were statistically significant.**

#### 3.3.1 Bottom line results for the impact of AI on inter and intra-reader variability, reliability, agreement

There is some evidence to suggest that using an AI model as a support tool may increase agreement between human readers and inter/intra variability. There is also evidence to suggest that agreement between readers may be improved more in those with less experience. However, as a limited number of studies reported this outcome firm conclusions cannot be made.

### 3.4 Impact of AI on cancer diagnostic time intervals

A total of five studies (breast n=3, lung n=1, prostate n=1) reported the impact of AI on cancer diagnostic time intervals, all of which explored the impact of AI on human reader interpretation of different radiological images (Calisto et al, 2022, Wataya et al, 2023, Forookhi et al, 2023, Pinto et al, 2021, Pacilè et al, 2020). These time intervals included time to diagnosis, assessment time, evaluation times, and reading time. The different time intervals were not clearly defined in most studies and as such are reported individually below. The findings appear to be inconsistent.

#### Time to diagnosis

One study reported the impact of AI models on diagnostic time. Calisto et al (2022) assessed the use of a DNN AI model (DenseNet) for the diagnosis of breast cancer from mammograms, ultrasound and MRIs. The findings showed that **when the 45 human readers (clinicians) used the AI model, the time to diagnosis was reduced** by 31% with an average of 308 seconds (s) (SD = 57.03s) when using the AI model in comparison with no assistance in which the average was 377s (SD = 44.56s). However, it is unclear if this difference was statistically significant.

#### Assessment time

One study reported the impact of the AI on assessment time. Wataya et al (2023) compared the performance of 15 human readers (radiologists with varying levels of experience) with and without the use of a deep learning AI model (CAD) for the diagnosis of lung cancer from CT images**. A statistically significant reduction in median assessment time was found when the readers used the AI model** (83.6s without AI to 69.9s with AI; p= 0.01). However, this difference was not seen for all radiologists, with the assessment time actually being prolonged when using the AI model for three of the 15 radiologists included, no reasons were provided for this difference.

#### Reading time

Two studies (breast n=2) reported on the impact of AI specifically on human reading times (Pinto et al, 2021, Pacilè et al, 2020). These studies used DBT images (n=1) and mammograms (n=1). Both of which found that the reading times changed depending on the level of suspicion/likelihood of malignancy.

Pinto et al (2021) assessed the use of an AI model (Transpara. V1.6.0) for the diagnosis of breast cancer from DBT images. **Reading times of the 14 human readers (radiologists) were shown to decrease when using AI for low suspicion examinations (by 8%) but increase when using AI for high suspicion** examinations (by 28%), however no significant differences were found (p=0.35).

Similarly, Pacilè et al (2020) assessed the effects of an AI model (MammoScreen V1) for the diagnosis of breast cancer from mammograms in 14 human readers (radiologists). Reading time was considered from the opening of a new case until a BI-RADS score was provided. **For images with a low likelihood of malignancy the time was similar** in the first reading session and slightly decreased in the second reading session. However, **for images with a higher likelihood of malignancy, the reading time was on average increased with the use of AI.**

#### Time for entire process of image interpretation

One study reported the impact of AI on a range of time outcomes. Forookhi et al (2023) compared the diagnostic accuracy of four human readers (radiologists with different levels of experience), with and without the use of a commercially available AI model (Quantib®) for the diagnosis of prostate cancer using mpMRI images. Reporting time included four different time intervals (mean uploading time, mean time taken for segmentation and contouring, mean time taken for lesion identification, mean report generation time). **The use of the AI model led to a statistically significant increase in reporting time** which ranged from 123.81s (+/− 51.25s) to 189.14s (+/− 67.08s) without Quantib®, and from 697.44s (+/− 98.88s) to 792.47s (+/− 122.37) with Quantib®. (p < 0.001). The uploading time was reported to be the most time-consuming step, followed by the time for segmentation and time for lesion identification.

#### 3.4.1 Bottom line results for the impact of AI on cancer diagnostic time intervals

The evidence regarding the impact of AI on cancer diagnostic time intervals appears to be inconsistent. Time to diagnosis and assessment times were reported to decrease, however these findings were only reported by individual studies meaning firm conclusions cannot be made. The evidence for reading times appears to show that the use of AI could increase or decrease reader time compared to reader only, depending on the suspicion level or likelihood of malignancy. Further research would be needed to better understand the impact of using AI on the workflow of cancer diagnosis.

### 3.5 Clinicians’ acceptance and receptiveness of the use of AI for cancer diagnosis

One study (Calisto et al, 2022) assessed the use of a DNN AI model (DenseNet) for the diagnosis of breast cancer from mammograms, ultrasound and MRIs and explored the acceptance and receptiveness of the human readers (clinicians) who utilised the AI model using a questionnaire**. A total of 98% of the 45 clinicians questioned suggested that they understood what the system was thinking, 93% trusted the AI models capabilities, and 91% were accepting of and preferred the AI approach**.

#### 3.5.1 Bottom line results for clinicians’ acceptance and receptiveness of the use of AI for cancer diagnosis

There is evidence to suggest that human readers (clinicians) may understand and trust the use of AI and in general may be accepting of using AI when diagnosing cancer in practice. However, the evidence was reported by one study only and as such firm conclusions cannot be made. Further research would be needed to better understand the acceptability of clinicians when using AI for cancer diagnosis.

## 4. DISCUSSION

### 4.1 Summary of the findings

The mapping exercise showed that there is a large volume of early stage (developmental and validation) research studies of AI models in cancer diagnosis, covering a wide range of cancers, but none of these studies evaluated the implementation of the AI models in clinical practice. There are a number of studies that have evaluated previously developed or commercially available AI tools, which may be useful to inform practice.

There is evidence to suggest that AI models may be effective at improving cancer diagnosis accuracy however, the evidence appears to be limited and the findings were not always statistically significant. No study reported findings that showed an overall greater degree of diagnostic accuracy in the control group compared to the AI. All studies that were identified showed significant improvements or no significant differences when compared to human readers or other conventional methods, and when the AI assisted human readers.

Regardless of how the AI model was used (i.e., compared to readers or used to assist readers) or the cancer type being diagnosed, studies were identified that demonstrated the benefit of using AI in cancer diagnosis was dependent on the level of experience of the human reader. Evidence suggests that AI models may have a similar level of diagnostic accuracy compared to experienced human readers (clinicians or radiologists) but may increase the accuracy of less experienced human readers when used as a support tool. However, the criteria for being classed as a more or less experienced reader varied between studies so it is unclear how strong this impact is overall.

When comparing a range of AI models several factors were reported to improve diagnostic accuracy. This included pre-training the AI model, adding additional classifiers or combining models to build ensemble models. However, these findings were reported by individual studies and as such further evidence would be needed to confirm this.

Inter and intra-reader variability, reliability, agreement, was reported by a limited number of studies (n= 4). While all studies reported overall improvements, it was noted in one study that improvements were not reported for each human reader although no exploration as to why this occurred was provided. Another study again highlighted the effectiveness of an AI model to improve agreement was dependent on the level of experience of the human reader.

The evidence regarding the use of AI as a time saving measure in cancer diagnosis was also inconsistent. A limited number of studies (n=5) reported on the impact of AI on time and the specific time outcome reported varied between studies. While diagnostic time and assessment time were reported to be reduced overall when using an AI model, the overall image interpretation time was found to be increased when using AI. However, these outcomes were only reported by individual studies, so findings should be interpreted with caution. Two studies identified the impact of AI on reading times was dependent on the level of suspicion or likelihood of malignancy. However, this is based on very limited evidence and as such firm conclusions cannot be made. There was also very limited evidence to suggest that clinicians may be accepting of AI when used as a support tool in cancer diagnosis.

While the results of the quality appraisal showed minimal concern regarding applicability to the review question, the majority of studies had some methodological limitations which led to an increased risk of bias (see Table 7, Fig.1). This was primarily related to the patient image selection process, which was often poorly described within the included studies and how the index tests (comparators) were conducted. In some cases, the same images were used for the human readers to interpret first without the use of the AI model and then with the AI model (with a short time gap between them) which could have introduced bias as the human reader plus AI group would have already seen all of the images being studied when the AI tool was introduced. The reference standard used also varied across studies and in one study the reference standard was not clearly stated.

**Figure 1.**
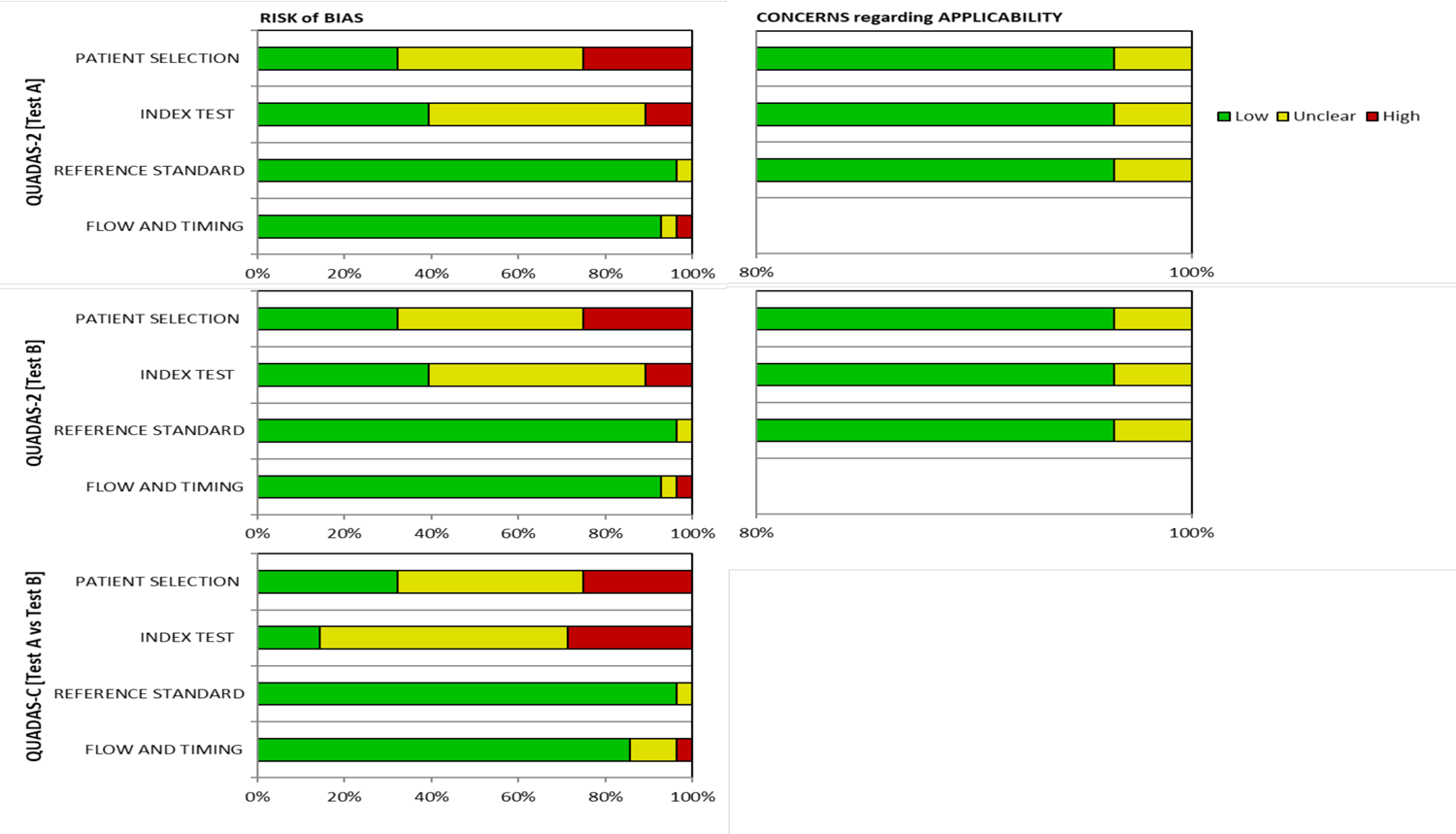
Graphical display of quality appraisal results for studies included in the synthesis of the rapid review

The considerable variation in the individual AI models studied, type of cancer being diagnosed, and types of images being used (e.g. MRI CT, X-ray etc.), as well as the methodological limitations of included studies could limit the applicability of these findings and the results should be interpreted with caution. It should also be noted that some studies did acknowledge potential conflicts of interest as the authors worked for the company that developed the AI model. However, this may be expected, as not all AI models included in the in-depth synthesis were commercially available. The overall findings may also be subject to publication bias, were studies that have identified AI to be less effective than a control are not submitted for publication and therefore not included here.

The National Institute for Health and Care Excellence (NICE) (2019) have created the Evidence Standards Framework For Digital Health Technologies which outlines the standards a health technology, such as AI would need to meet in order to show its effectiveness for use in the UK. However further standards are to be developed for AI models using adaptive algorithms that continually update. It was unclear from the included studies whether the AI models assessed were fixed or adaptive, however only one study described the use of an AI model, Red Dot Behold.ai, that was commercially available in the UK (Tam et al, 2021). This model has been suggested to be able to diagnose multiple conditions including lung cancer and stroke using CT or X-ray images, further details about this model and its potential uses can be found online (<u>behold.ai</u>). While this model has been FDA and CE approved and CQC registered, it is unclear if it has received approval from NICE. However, the model was included in a recent early value assessment published by NICE exploring the use of AI in analysing chest X-rays for suspected lung cancer, further details on the findings of the assessment can be found online (<u>NICE</u> 2023).

### 4.2 Strengths and limitations of the available evidence

All included studies in the in-depth synthesis were comparative in design and as such were best suited to explore effectiveness of AI models in cancer diagnosis in a real-world setting. The evidence included was published within five years of this rapid review being conducted which should increase the relevancy of the findings. Despite some methodological limitations, all included studies were published in peer-reviewed journals.

The majority of the included studies (n=25) were retrospective and gained the images being studied through historical datasets, as such the data used may not reflect the real-world impact of incorporating AI into the healthcare sector.

The findings also highlighted several evidence gaps. As can be seen in Table 1, none of the studies that met the inclusion criteria reported any findings related to patient outcomes (including harms), economic outcomes or any outcomes related to equity. As the majority of studies were retrospective or utilised images from patients who had already been diagnosed with or without cancer no evidence was found to show AI may be effective in diagnosing cancer in a genuine real-world setting. While two studies were conducted in the UK only one of these described using an AI model that was commercially available in the UK. It was unclear if the AI models used for cancer diagnosis are able to be replicated in Wales.

There was limited evidence to suggest the use of AI could reduce the time to diagnosis and the perceptions of clinicians. Due to the heterogeneity of included studies, caution should be applied when interpreting the findings of the review.

The included studies explored the use of a range of AI models, which may explain the inconsistent findings, and is likely to limit the generalisability of the findings to all AI models.

Key details pertaining to the dataset used, and type of AI model were often lacking or not clearly reported within the included studies. Study designs were also poorly reported across included studies and the cancer type and imaging technique varied across studies, which could limit the generalisability of the findings to specific contexts.

### 4.3 Strengths and limitations of this rapid review

The studies included in this rapid review were identified through a comprehensive search of electronic databases. Despite making every effort to capture all relevant publications and reduce the risk of bias in our review process, it is possible that additional eligible publications may have been missed. To ensure the usefulness of our findings, only comparative studies were included in the review, as these are better placed to determine the presence of cause-and-effect relationships when exploring effectiveness.

AI is a complex and fast developing field. Multiple AI models have been assessed within the literature over time, and AI models that have shown promising results are also continually developed, adding for example additional classifiers etc to improve performance. As such, it is challenging to collate the evidence as even within a few months or years the technology evaluated within this rapid review is likely to be outdated compared to the newly developed advanced AI models. This development in technology also makes it difficult to directly compare different AI tools. The reference standard used across studies also varied including histopathologic examinations, decisions made by expert radiologists or follow-up and in one study the reference standard was not clearly stated further limiting the generalisability across studies.

It is also important to note that although the QUADAS-2 tool and its extension QUADAS-C are designed to assess the methodological quality of diagnostic and comparative study designs included in this rapid review, they are not designed to assess any methodological issues related to the use of the AI models. The review team is aware of a further adapted extension to the QUADAS tool (QUADAS-AI) that is due to be published in future, which would better assess specific methodological considerations relating to AI models. However, this extension was unavailable at the time this rapid review was conducted. Sounderajah et al (2021) highlighted potential biases that could occur when using AI. This included the use of open source datasets as these datasets may include duplications, may contain images that are not labelled correctly and may have incomplete data. As such, the results of the quality appraisal should be interpreted with caution. Furthermore, QUADAS-2 may not have been sufficient to assess the quality of the studies for evaluating outcomes other than diagnostic accuracy.

### 4.4 Implications for policy and practice

This rapid review has provided an insight into the effectiveness of AI in cancer diagnosis, and factors that can impact the accuracy of AI models, such as the level of experience of the individual interpreting the AI results. This information may be useful when planning how best to incorporate AI into the health and care sector. Further well-designed high-quality research is needed from the UK and similar countries to better understand the effectiveness of AI in cancer diagnosis. However, given the pace of development in this field, it is difficult to make recommendations on one specific AI tool for use in radiology diagnostics for cancer.

Although the focus of the in-depth synthesis was on the use of AI for interpreting radiological images in breast, lung and prostate cancer diagnosis, other important areas in which AI could provide benefit were also identified during screening and during the mapping exercise. These included the use of AI for radiological image quality improvement, differentiating between different types of cancers (e.g. between glioblastoma and primary central nervous system lymphoma), or in the detection of metastases. These areas should be considered when planning for AI incorporation into the health and care sector.

### 4.5 Implications for future research

– Further research is needed to explore the effectiveness of AI models for cancer diagnosis in a real world setting and to evaluate the ongoing use of AI in cancer diagnosis.
– Further research is needed to validate the use of specific AI models.
– Further research is needed to determine in which context the use of AI would be most effective (e.g. as a support tool for less experienced clinicians/radiologists)
– Further research is needed to explore the impact of cancer diagnosis using AI on patient harm, costs, and equity.

### 4.6 Economic considerations*

- In theory it might be possible for AI to assist with earlier diagnosis of cancer with both health and economic benefits. However, there are currently no minimum requirement guidelines in terms of effectiveness or cost-effectiveness of AI use in cancer screening in the UK. Work from Vargas-Palacios (2023) and colleagues aims to develop such guidelines.
- There is little evidence on the cost-effectiveness of using AI for cancer diagnosis. One modelling paper from the United States (US) suggests using AI in lung cancer screening low-dose computerised tomography (CT) scans can be cost-effective, up to a cost of $1,240 per patient screened, giving a willingness-to-pay of $100,000 per quality-adjusted life year (QALY) gained (Ziegelmayer et al, 2022). This is high in comparison to US and UK payer thresholds.
- The UK (and its constituent countries) perform consistently poorly against European and international comparators in terms of cancer survival rates (Arnold et al, 2019). Cancer screening was suspended and routine diagnostic work deferred in the UK as a result of the COVID-19 pandemic. Modelling suggests up to 3,620 avoidable additional deaths will occur between 2020 and 2025 due to the impact of the pandemic on cancer services (Maringe et al 2021).
- Later stage diagnoses (3 & 4) incur greater costs to the healthcare system across most colorectal, pancreatic, lung and kidney cancers (White 2023). Almost half (46%) of all cancer cases were diagnosed at stage 3 and 4 (out of those with a known stage at diagnosis) in England in 2018 (Cancer research UK, 2023).
- The cost of cancer to the UK economy in 2019 was estimated to be least £1.4 billion a year in lost wages and benefits alone (Hilhorst and Lockey, 2023). When widening the perspective to include mortality, this figure rises to £7.6 billion a year. Pro-rating both figures to the Welsh economy and adjusting for inflation gives figures of £79 million and £429 million per annum respectively (Bank of England, 2023).

*This section has been completed by the Centre for Health Economics & Medicines Evaluation (CHEME), Bangor University

## Data Availability

All data produced in the present study are available upon reasonable request to the authors

## 6. METHODS

The rapid review was conducted in two stages, which included an initial mapping exercise followed by a more in-depth review of a sub-set of study types that stakeholders considered to be the most relevant to inform practice.

### 6.1 Mapping exercise methods

#### 6.1.1 Eligibility criteria

We searched for primary sources to answer the review questions: ‘What is the effectiveness of artificial intelligence in radiology for cancer diagnosis?’ The following eligibility criteria were used to identify studies for inclusion in the rapid review.

**Table 4:**
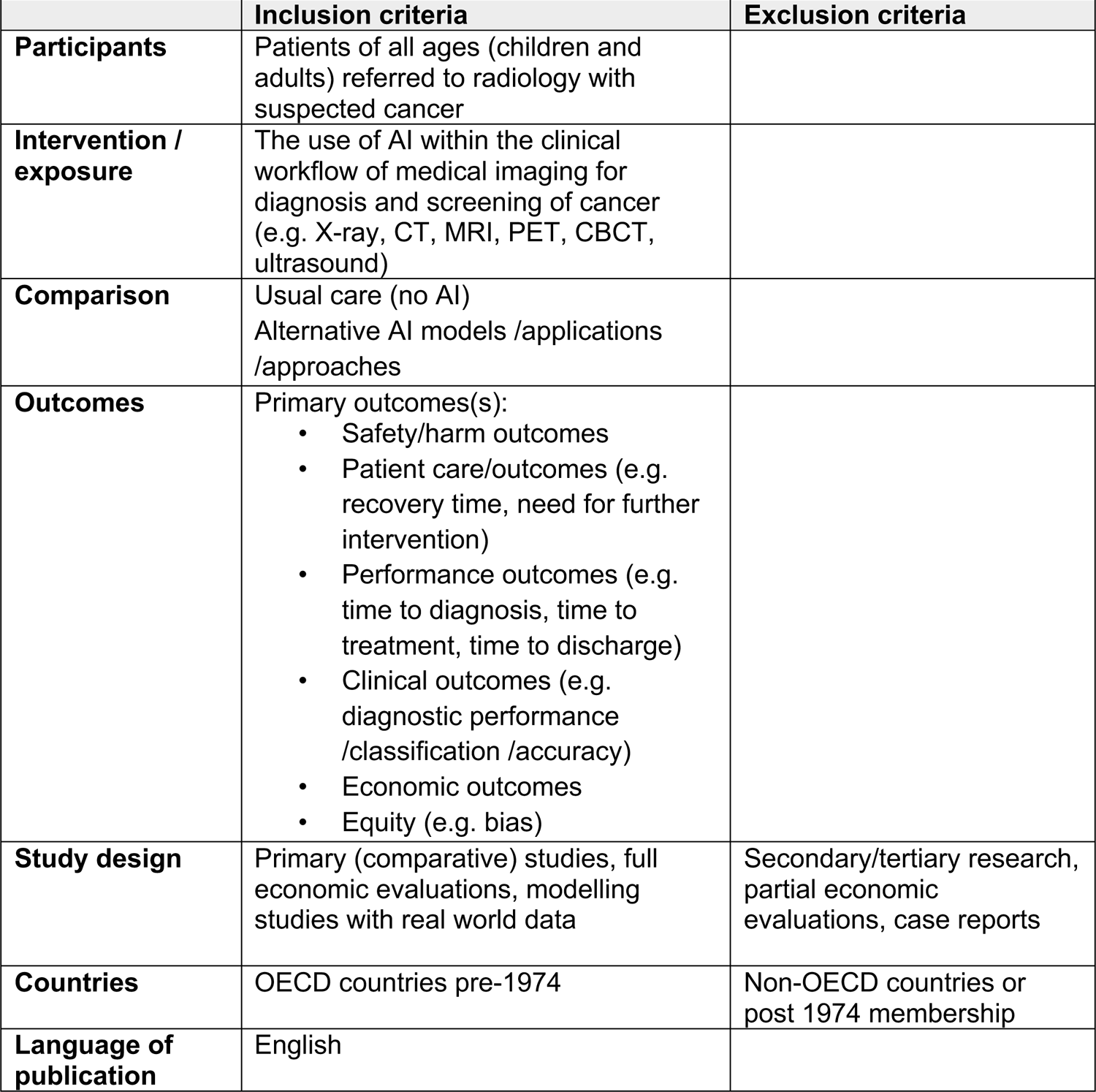
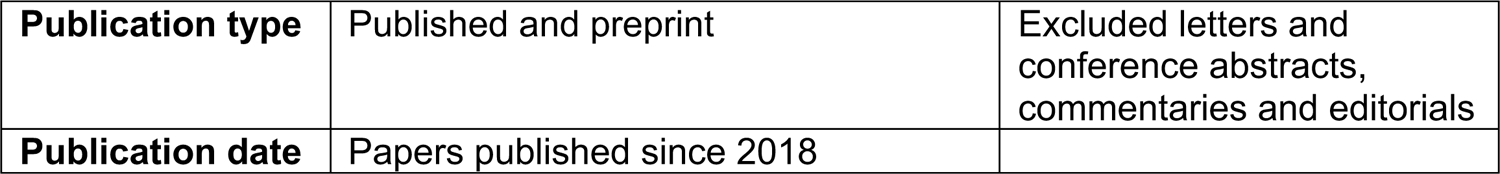
Eligibility criteria for mapping exercise.

#### 6.1.2 Literature search

A search of Medline (Ovid), Embase (Ovid), Cochrane Central Register of Controlled Trials (CENTRAL) and ScanMedicine (NIHR) was conducted on the 20^th^ June 2023. Terms to describe the key concepts of artificial intelligence, radiological imaging and cancer were utilised. Search concepts and keywords included artificial intelligence, deep learning, machine learning, neural networks, cancer, medical imaging (X-ray, CT, MRI, PET, CBCT, ultrasound). The searches included free text words and subject headings. The NICE OECD countries geographic search filter was used for the searches in Medline and Embase.

Searches were limited to English language publications that were published since 2018 and to primary studies. A total of 21,403 records were retrieved which were managed in Endnote 20. Following deduplication, 20,043 records remained. The search strategy used to search MEDLINE is available in Appendix 4.

#### 6.1.3 Study selection process

All studies were uploaded to the systematic reviewing platform Rayyan for title and abstract screening. All studies were screened by a single reviewer and to ensure consistency a proportion (5%) of studies were screened by two independent reviewers. Any conflicts were resolved within the team. A total of 640 articles were screened at full text by two independent reviewers, and any conflicts were discussed and resolved by a third reviewer. A visual representation of the flow of studies throughout the review can be found in Figure 6.1.

#### 6.1.4 Study design classification

The included studies were classified as diagnostic test accuracy studies.

#### 6.1.5 Classification of studies for map

All included studies were coded into the following categories:

- Type of cancer
- AI model development stage (commercially available, previously developed or developed specifically for the purposes of the study)
- The number of images used in the dataset
- Outcome measures reported in primary study
- The comparator (human or AI)

Once coding was complete, the map was constructed and presented to stakeholders to enable them to identify a focus for the rapid review.

### 6.2 In-depth synthesis methods

#### 6.2.1 Study selection process

Once a focus for the in-depth synthesis had been agreed with stakeholders, the 92 studies included in the map were rescreened for eligibility in the in-depth synthesis. A visual representation of the flow of studies throughout the review can be found in section 7.1.

#### 6.2.2 Eligibility criteria for the in-depth synthesis

**Table 5.** Eligibility criteria for inclusion in the in-depth synthesis.

|  | **Inclusion criteria** | **Exclusion criteria** |
| --- | --- | --- |
| **Participants** | Patients of all ages (children and adults) referred to radiology with suspected **breast, prostate or lung cancer** | Focus on other cancer types |
| **Intervention / exposure** | The use of AI within the clinical workflow of medical imaging for diagnosis of cancer (e.g. X-ray, CT, MRI, PET, CBCT, ultrasound) |  |
| **Comparison** | Usual care (no AI)<br>Alternative AI models /applications /approaches | No comparator |
| **Outcomes** | Primary outcomes(s):<br>• Safety/harm outcomes<br>• Patient care/outcomes (e.g. recovery time, need for further intervention)<br>• Performance outcomes (e.g. time to diagnosis, time to treatment, time to discharge)<br>• Clinical outcomes (e.g. diagnostic performance /classification /accuracy)<br>• Economic outcomes<br>• Equity (e.g. bias) |  |
| **Study design** | Primary (comparative) studies, full economic evaluations, modelling studies with real world data | Secondary/tertiary research, Commentaries, Editorials, partial economic evaluations, case reports |
| **Countries** | OECD countries pre-1974 | Non-OECD countries or post 1974 membership |
| **Language of publication** | English |  |
| **Publication date** | Papers published since 2018 |  |
| **Publication type** | Published and preprint |  |
| **Other considerations** | **Commercially available AI models and those that had previously been developed** | AI models specifically for the study |

#### 6.2.3 Data extraction

Data extracted was conducted by a single reviewer and was consistency checked by a second reviewer. Information extracted includes:

– Citation
– Study design
– Intervention (AI model)
– Comparator
– Study aim
– Data collection methods and dates
– Outcomes reported
– Sample size
– Participants
– Dataset details
– Cancer type
– Imaging technique
– Key findings
– Notes

#### 6.2.4 Quality appraisal

The QUADAS-2 tool and the QUADAS-C extension tool were used to assess the methodological quality of each included study. The QUADAS-2 tool is used to assess the quality of diagnostic accuracy studies, however it is not well suited to studies that have multiple index tests (comparators), as such, the QUADAS-C tool was also used in order to account for the comparative nature of the included studies.

Quality assessment was undertaken by a single reviewer, with verification of all judgements by a second reviewer. Any discrepancies were discussed and resolved amongst the review team. The results of quality appraisals for individual studies can be seen in section 7.3. Although some studies were rated as having a low risk of bias, the majority of included studies had some methodological limitations.

#### 6.2.5 Synthesis

A narrative synthesis was conducted reporting results from all included studies in the in-depth synthesis.

#### 6.2.6 Assessment of body of evidence

An assessment of the overall body of evidence was made based on the relevance of the available evidence in addressing the review question and sub-questions, the amount and quality of the evidence, the magnitude and direction of effects and consistency in the findings, and clinical heterogeneity.

## 7. EVIDENCE

### 7.1 Search results and study selection

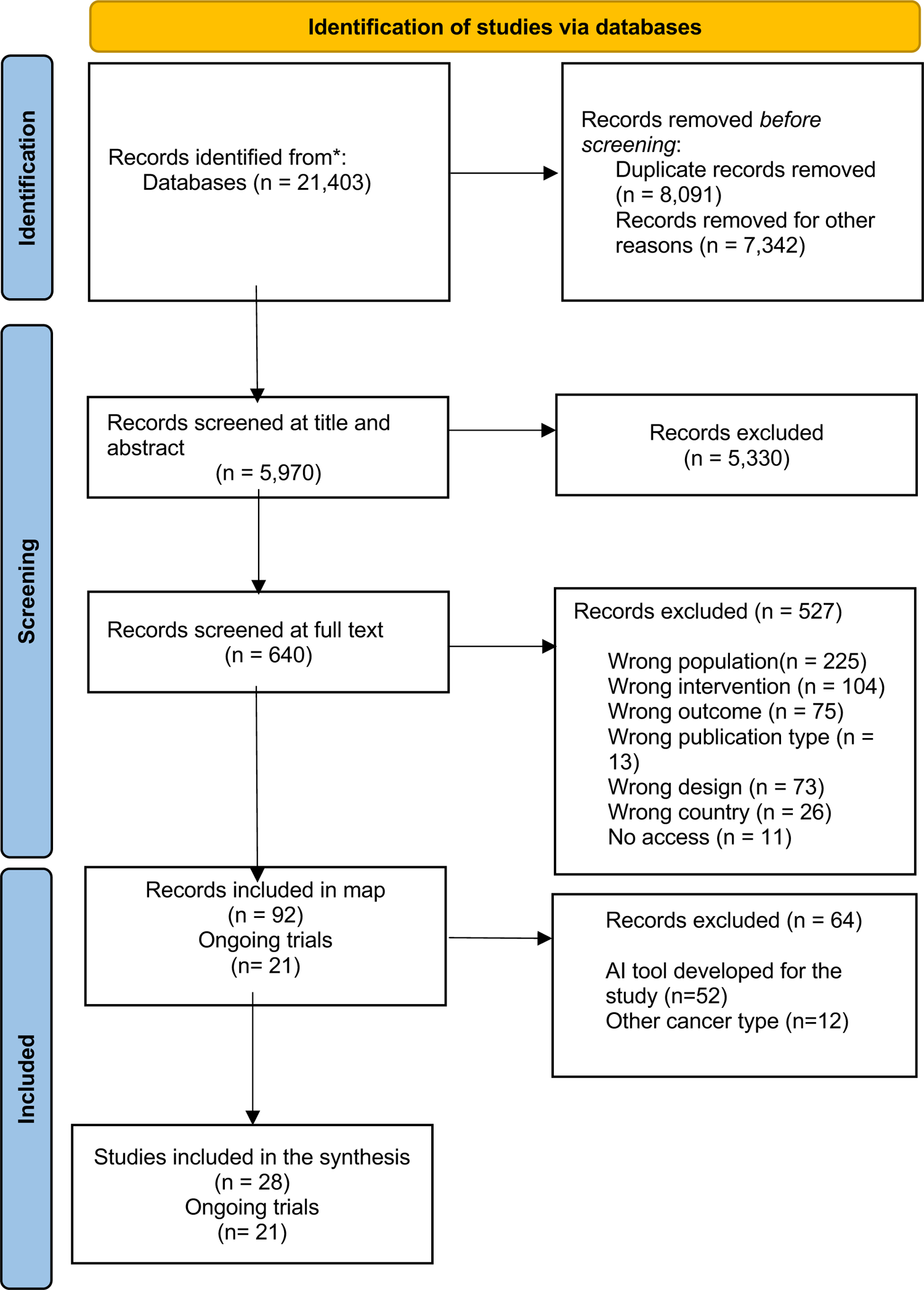

### 7.2 Data extraction

**Table 6:**
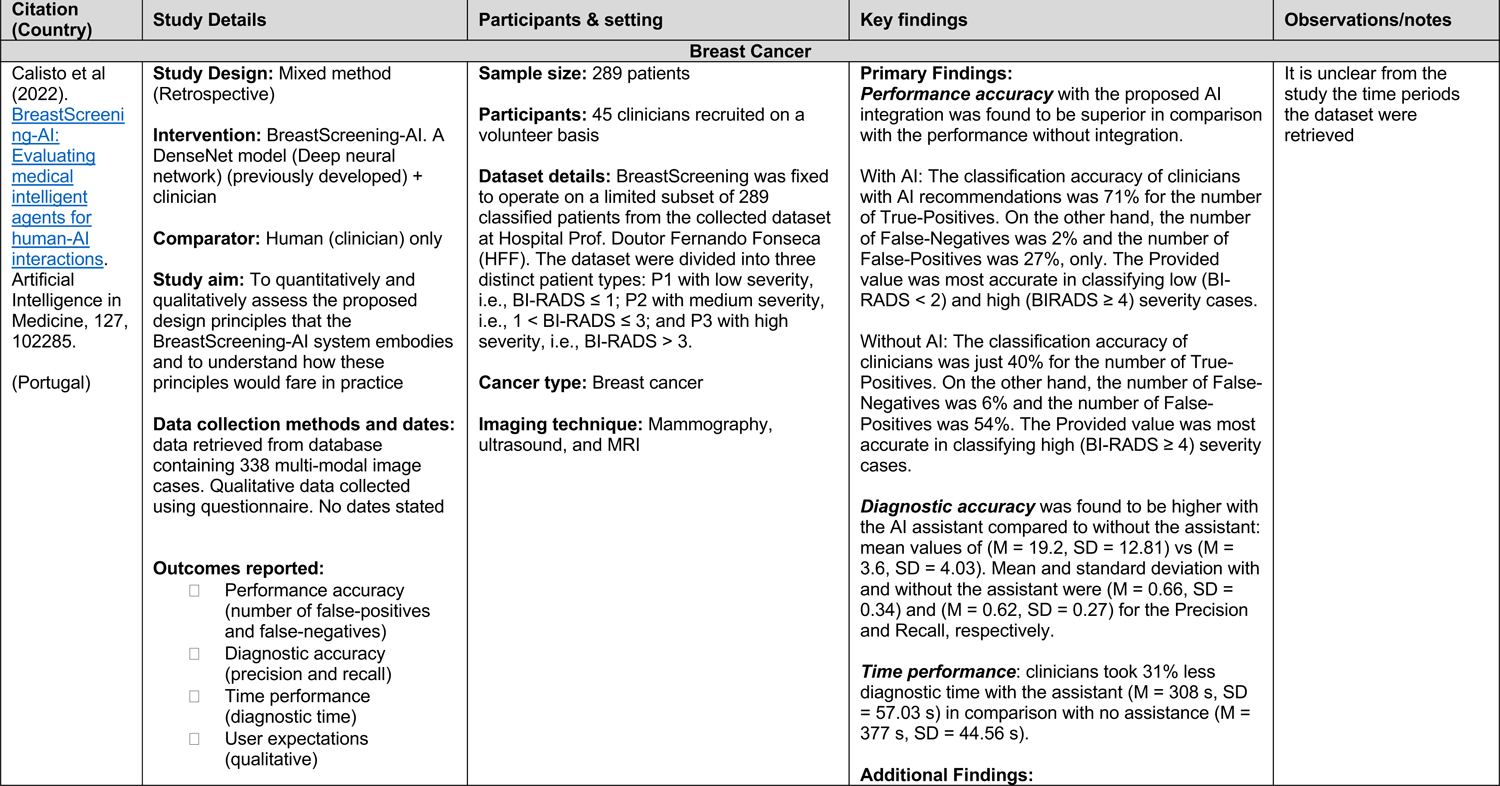
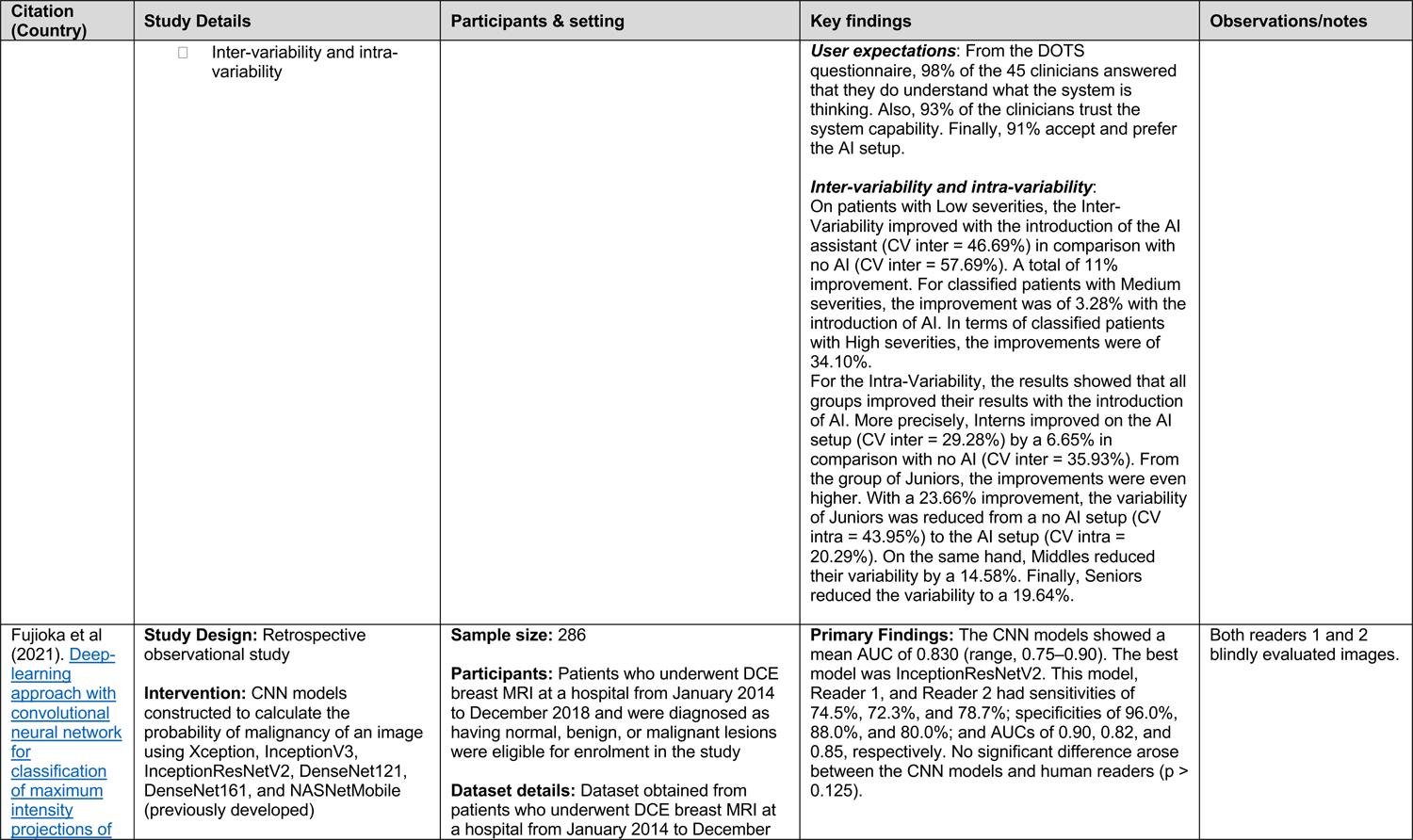
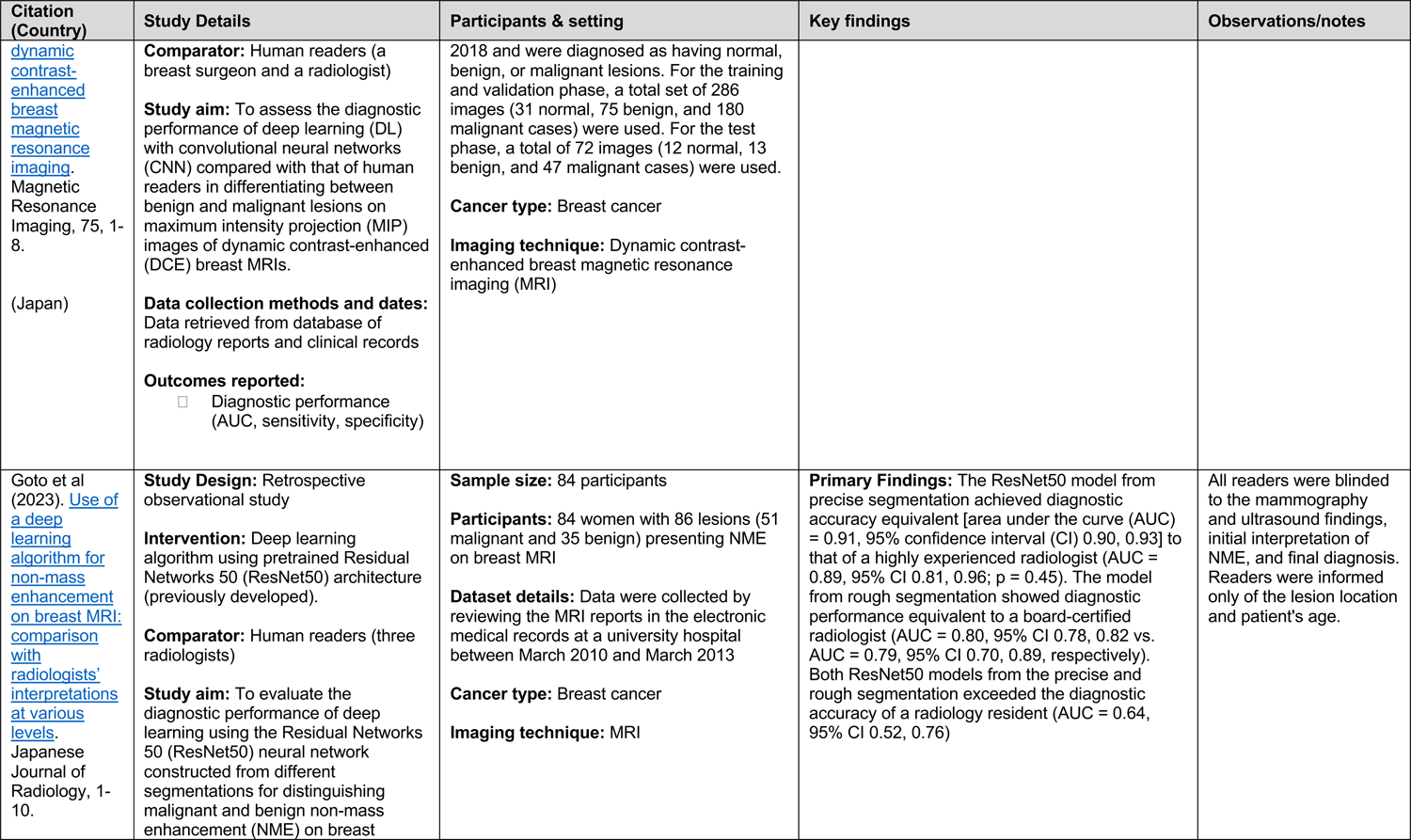
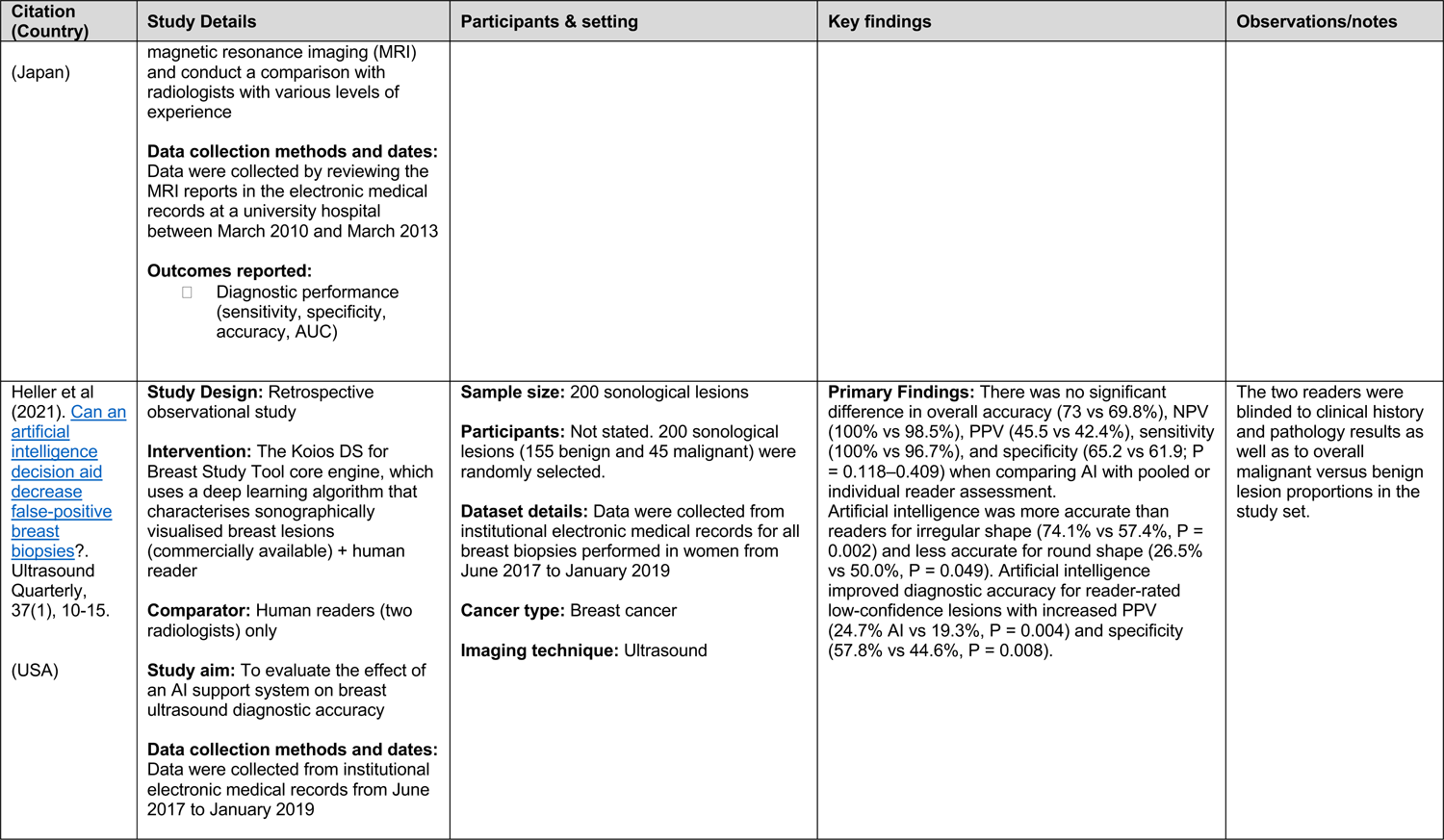
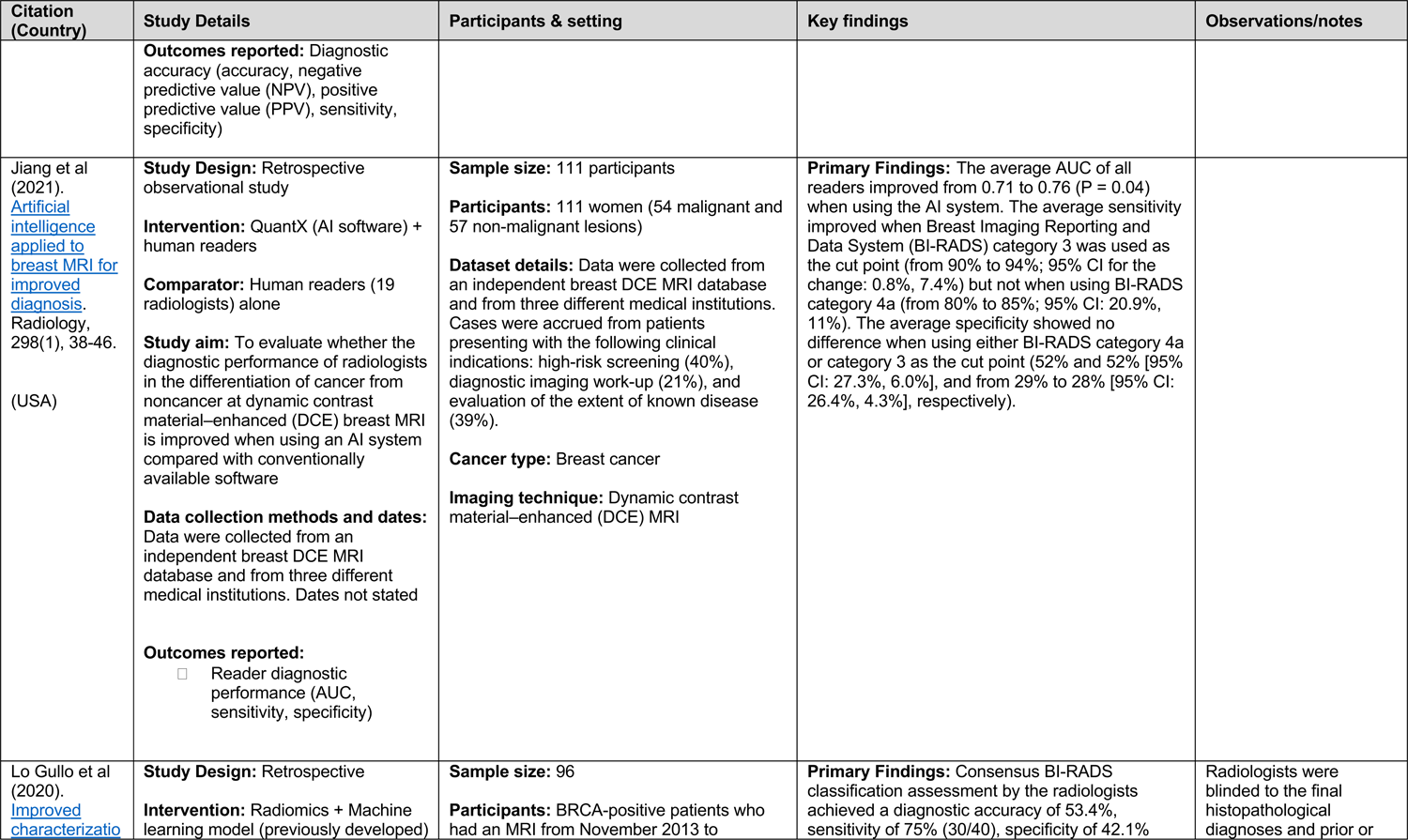
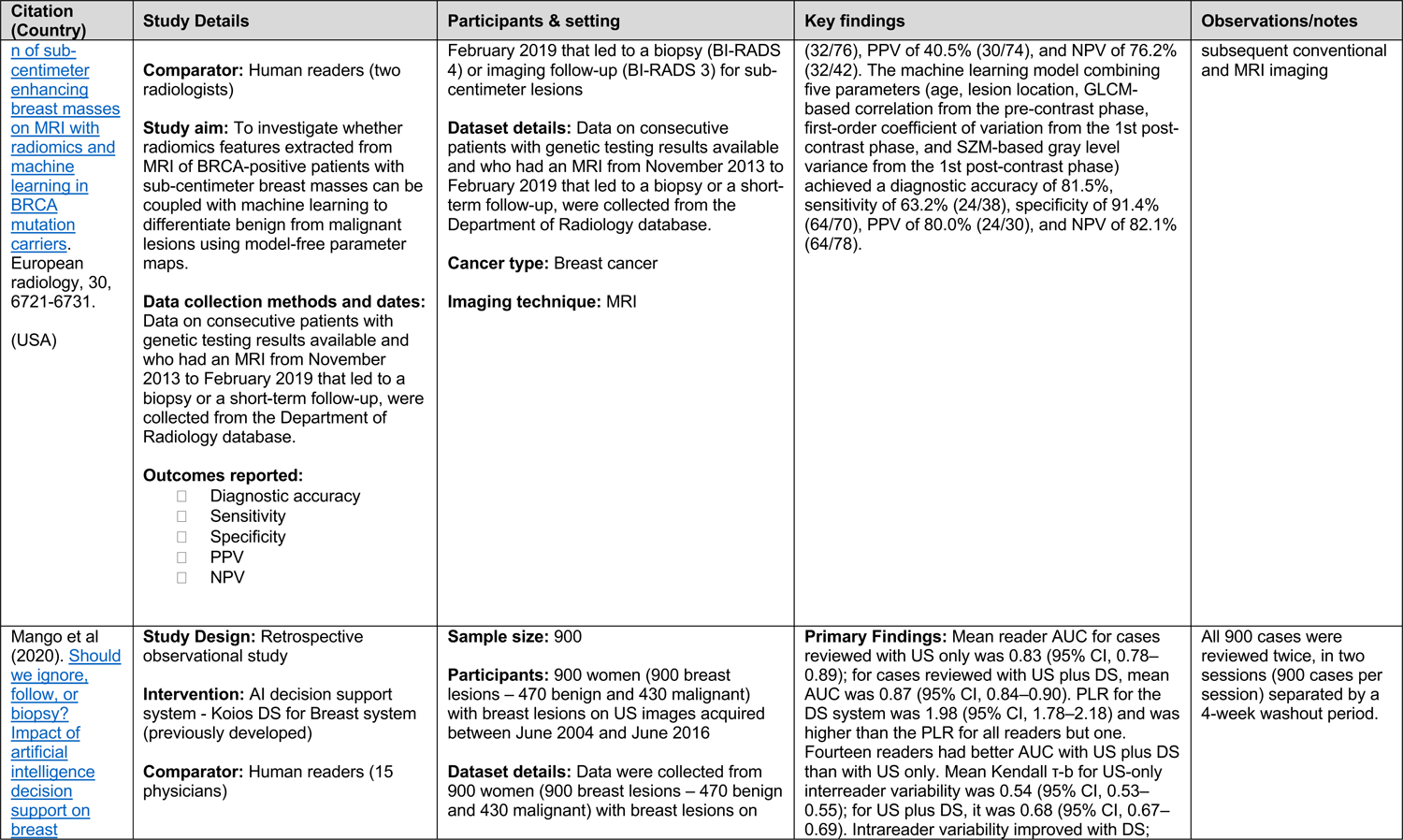
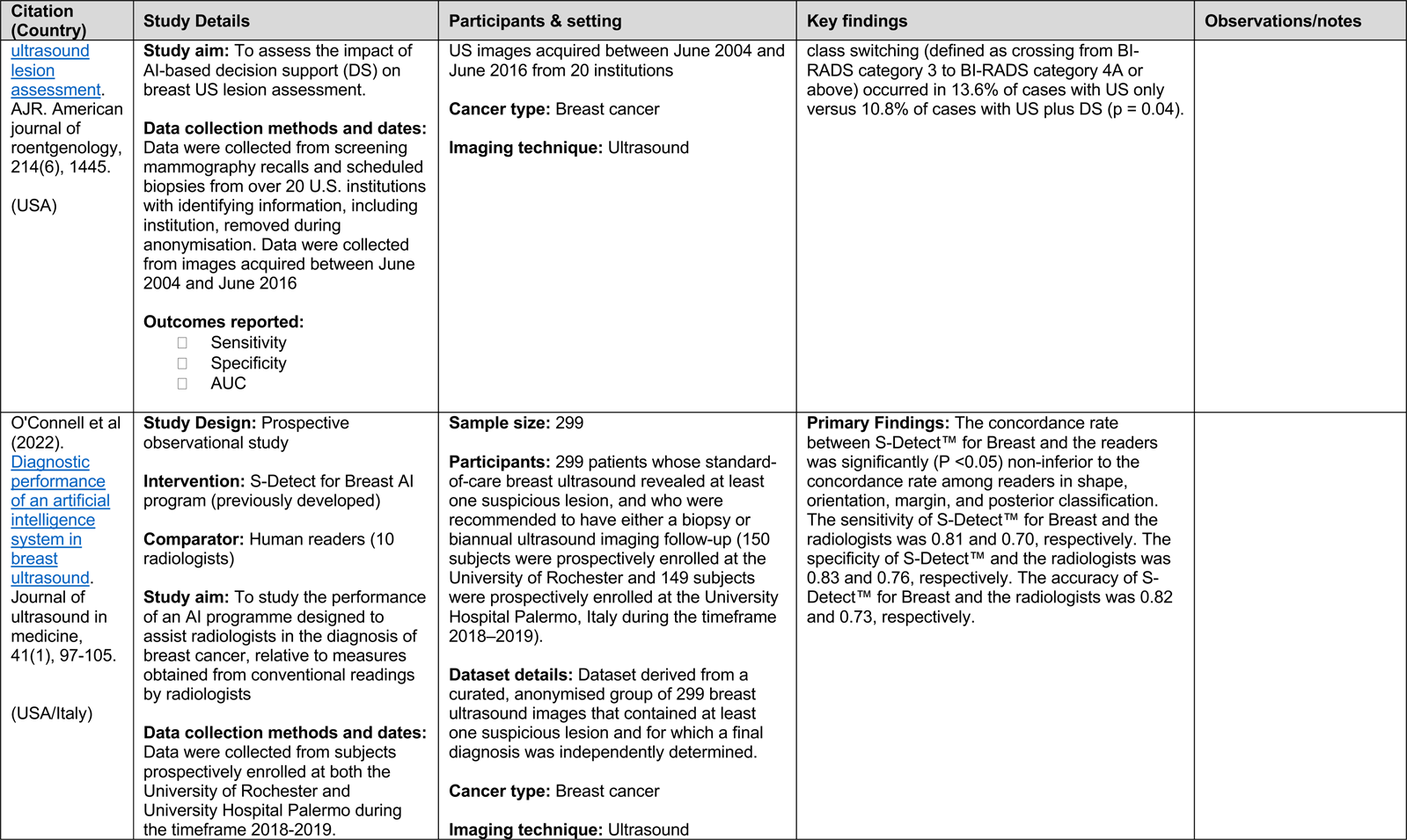
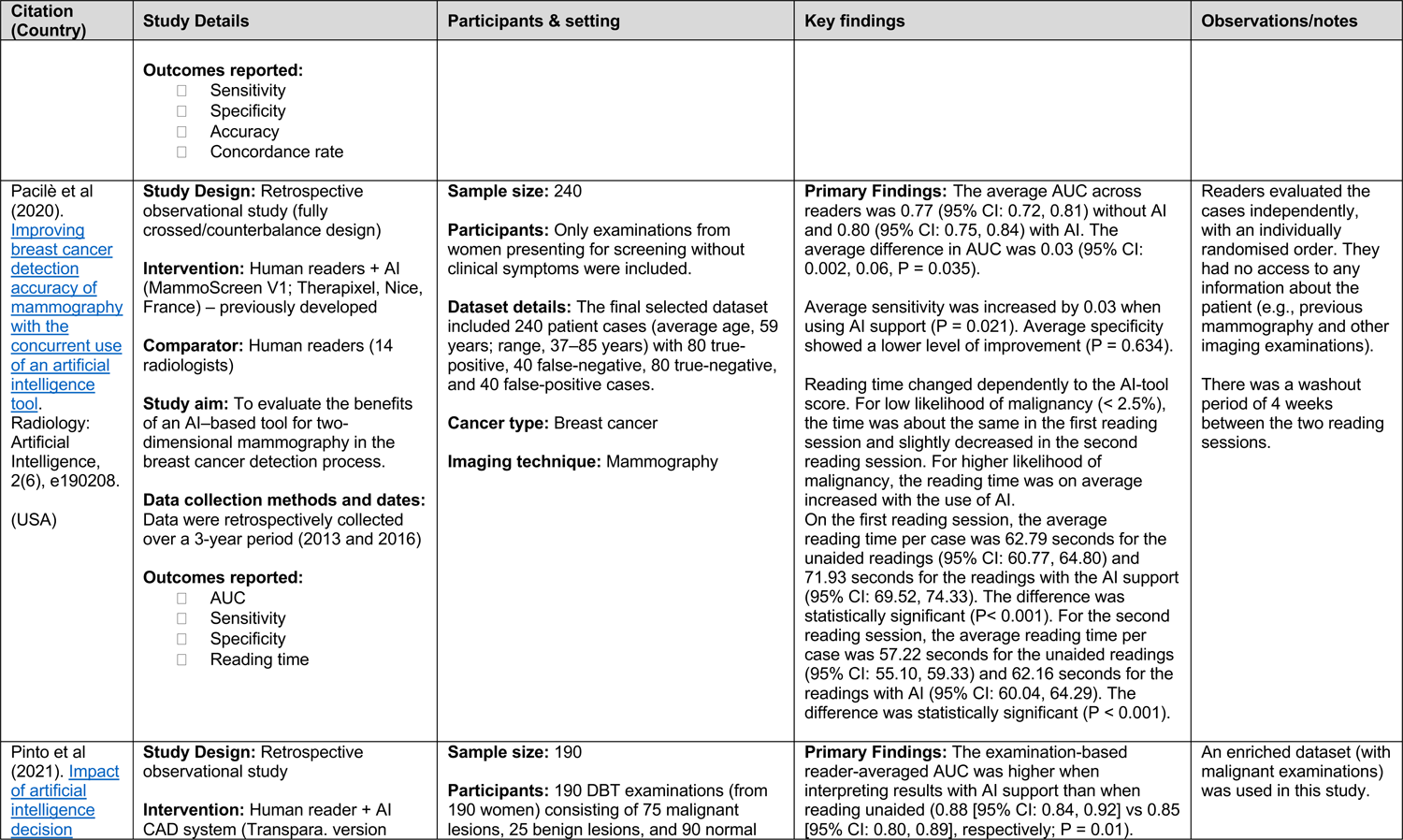
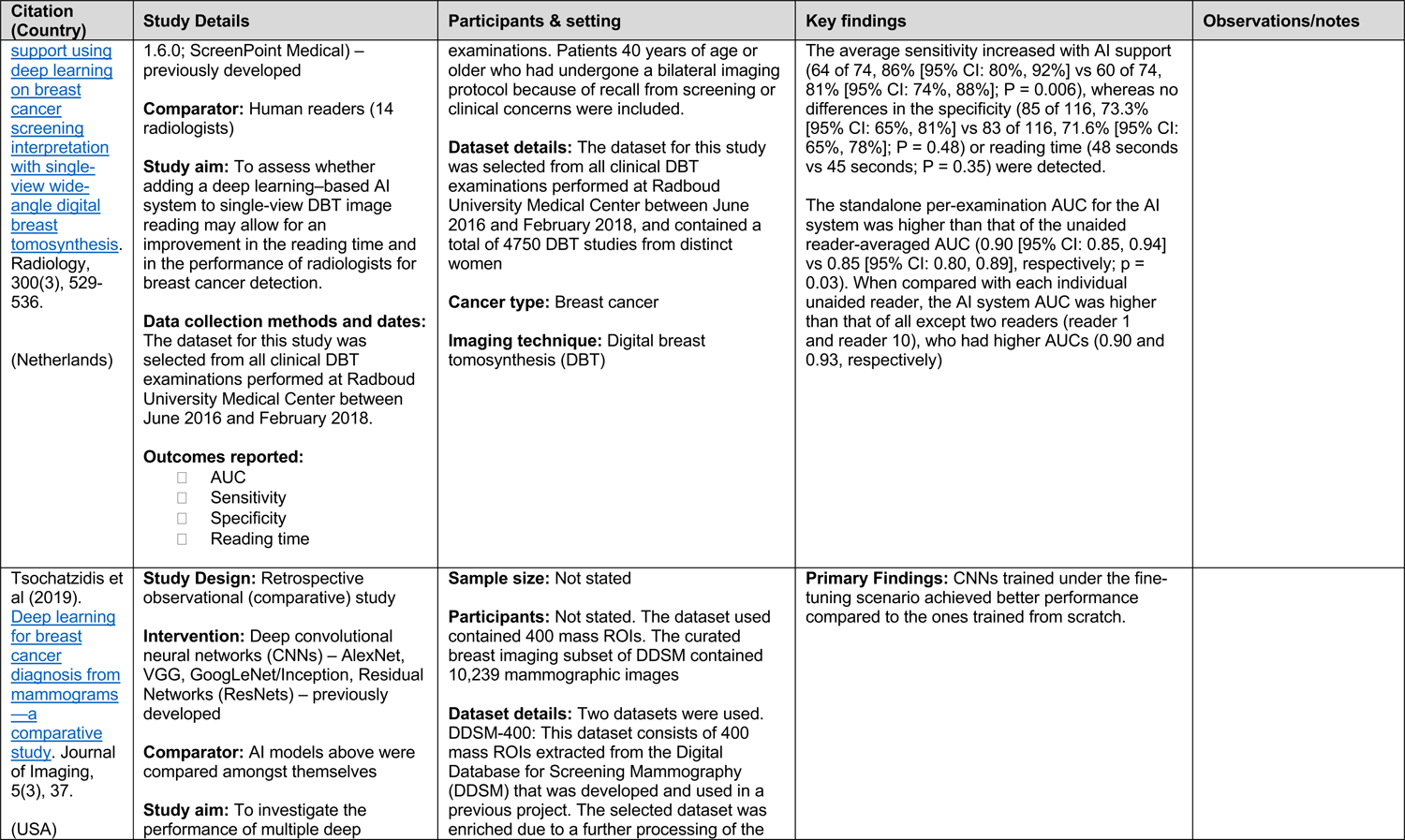
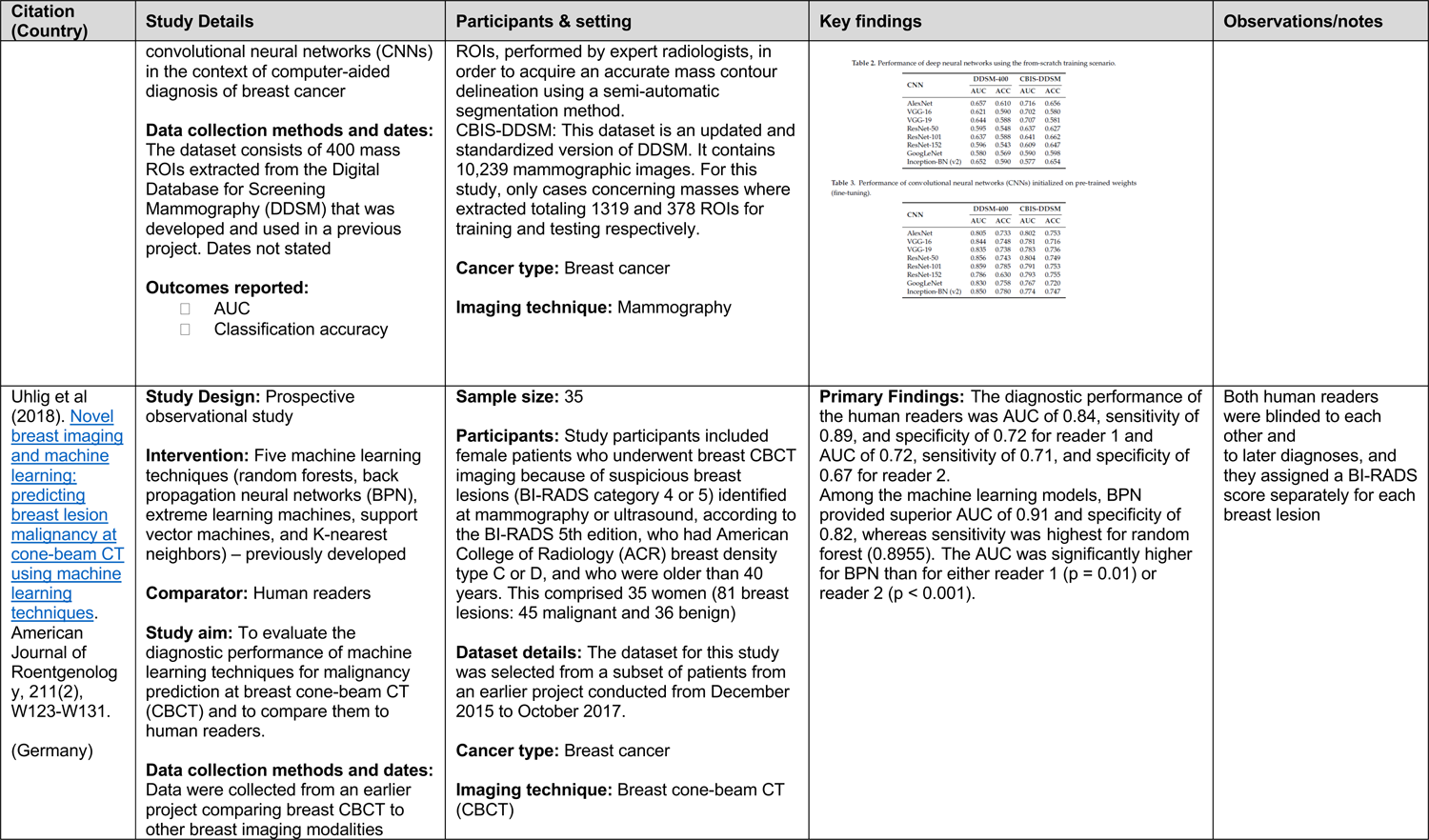
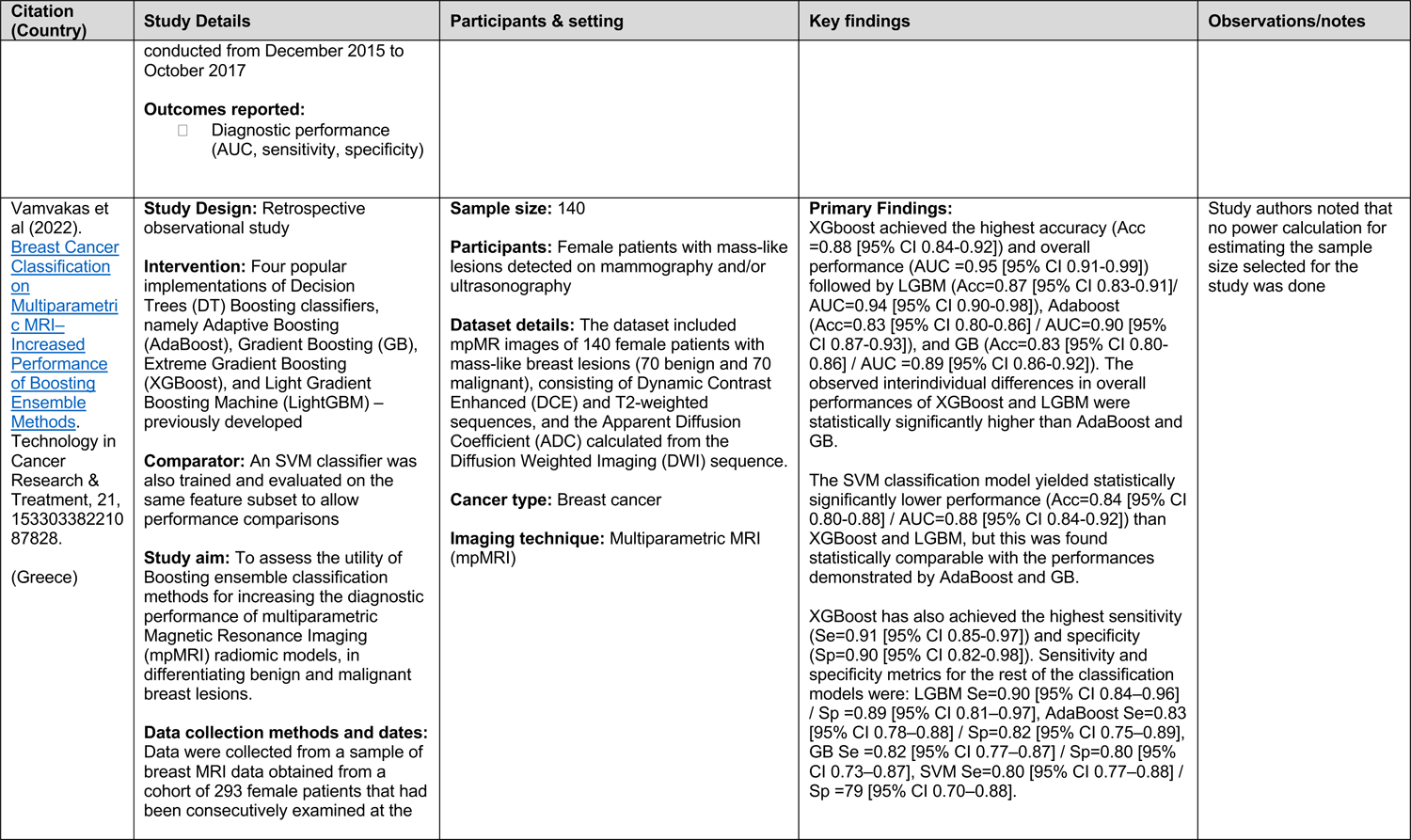
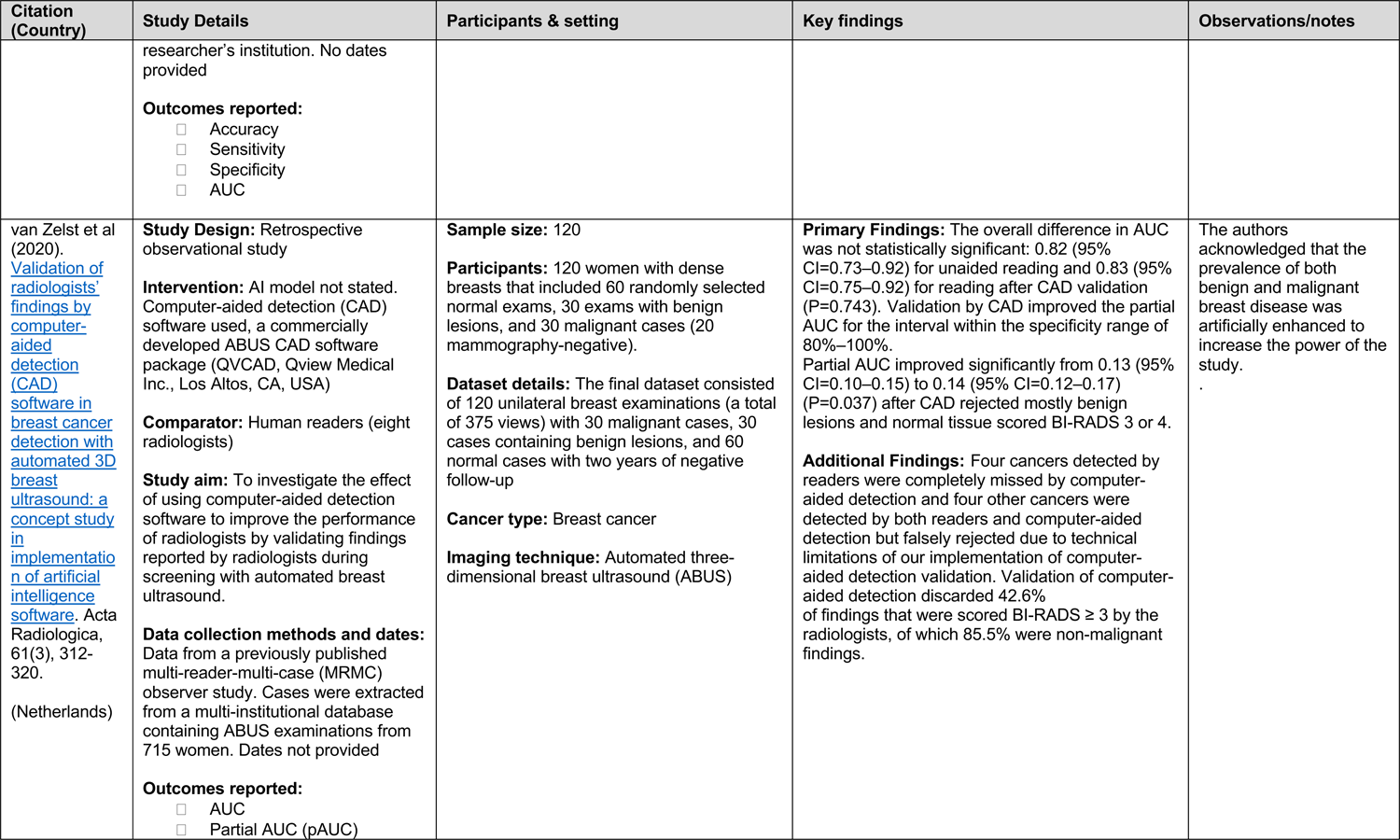
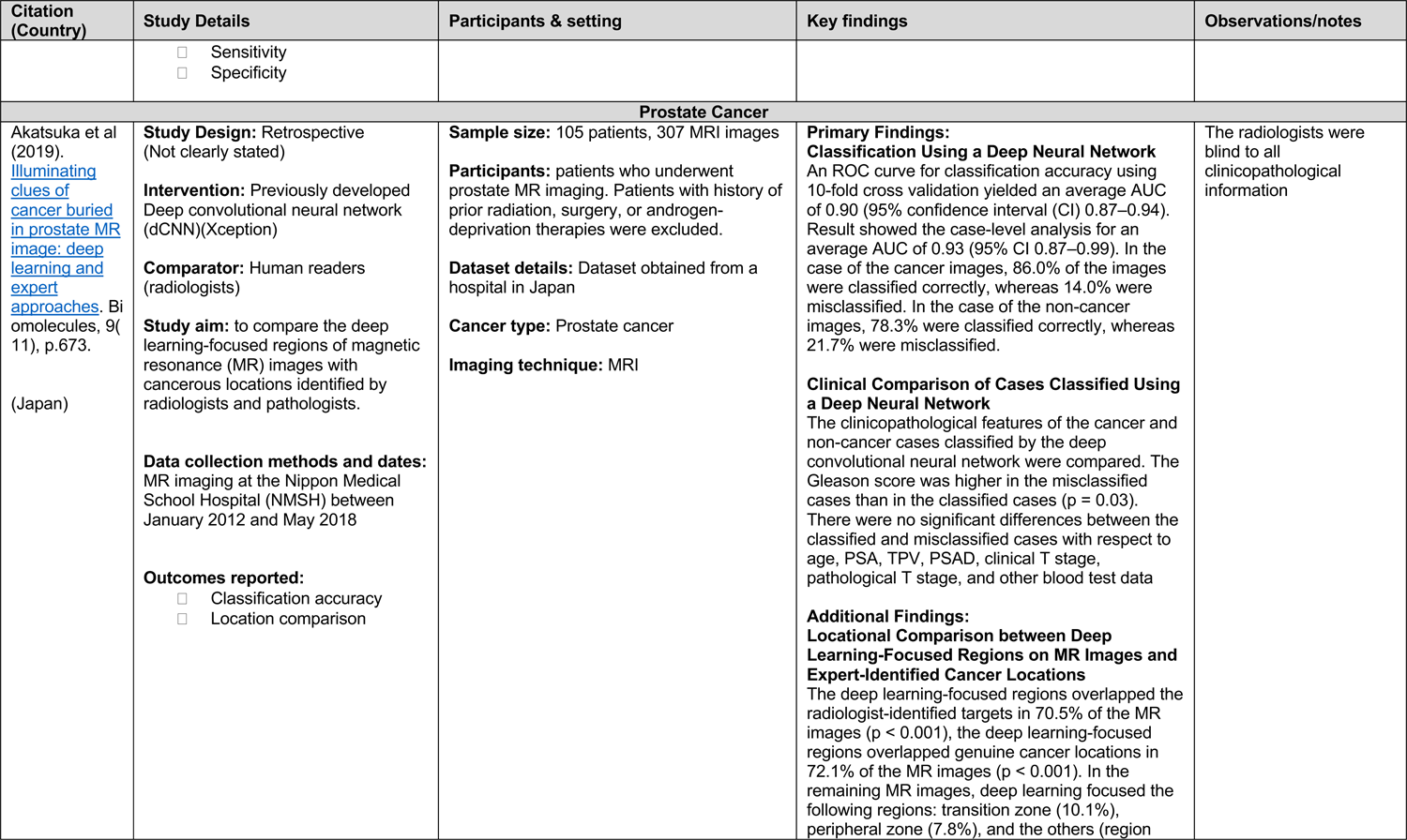
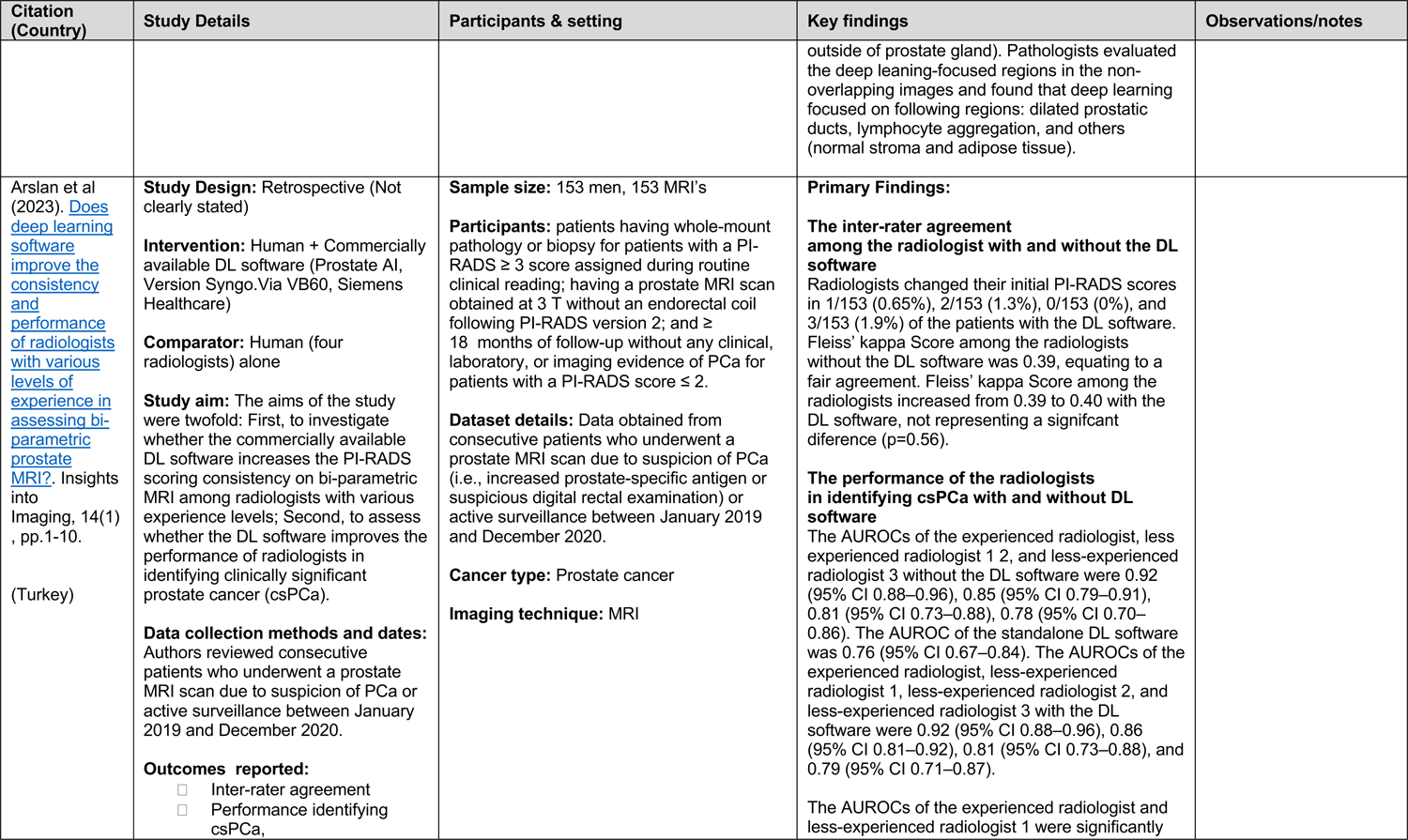
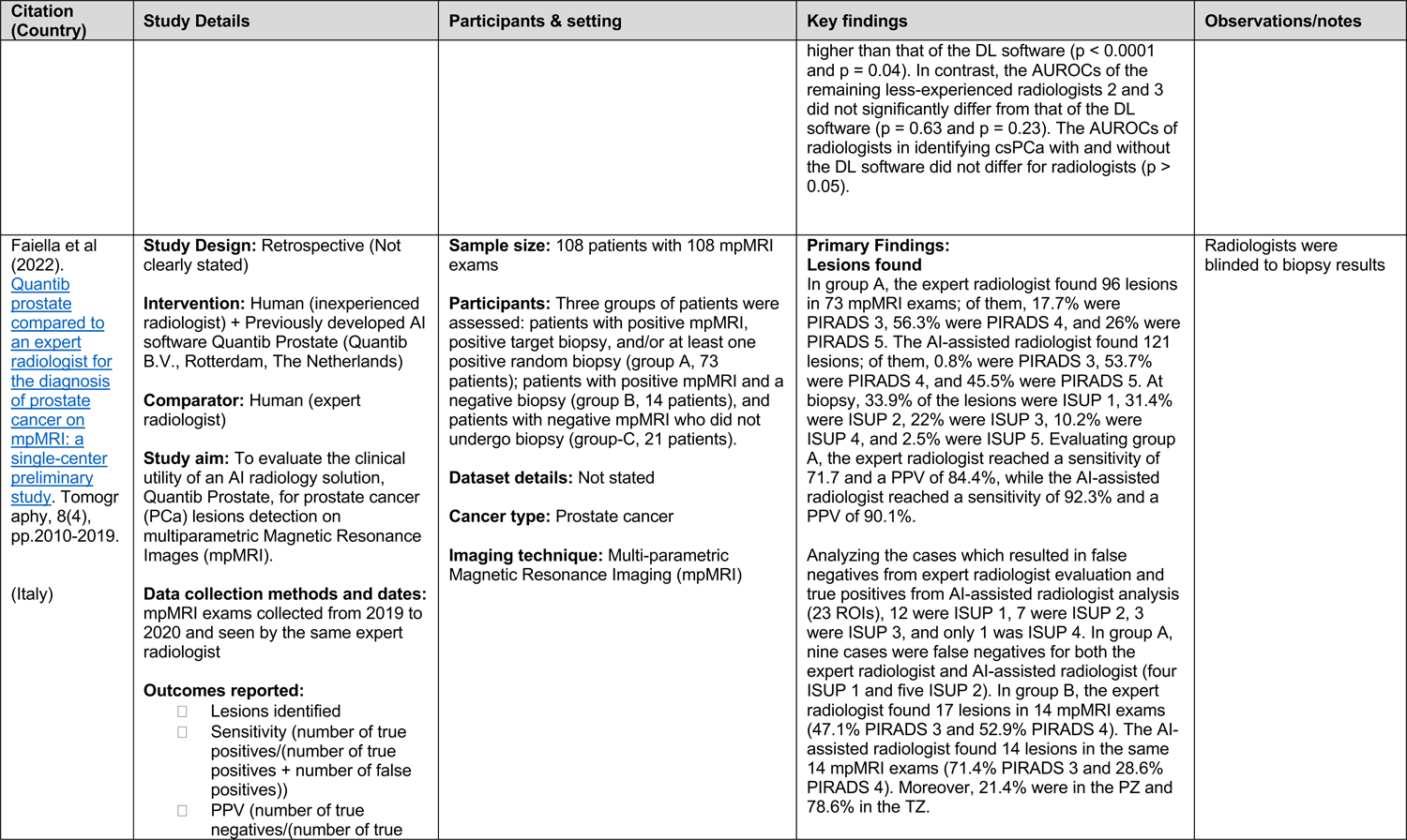
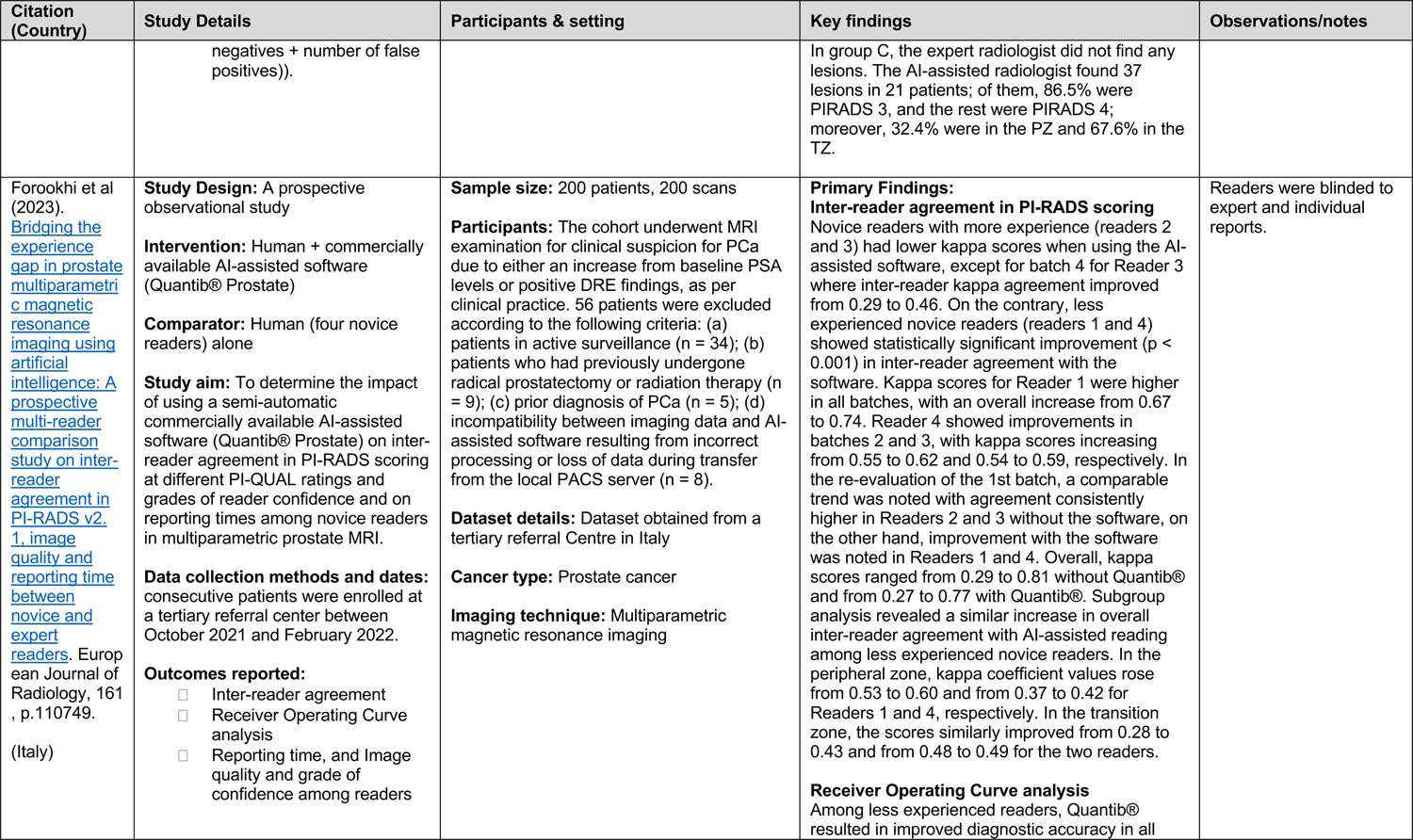
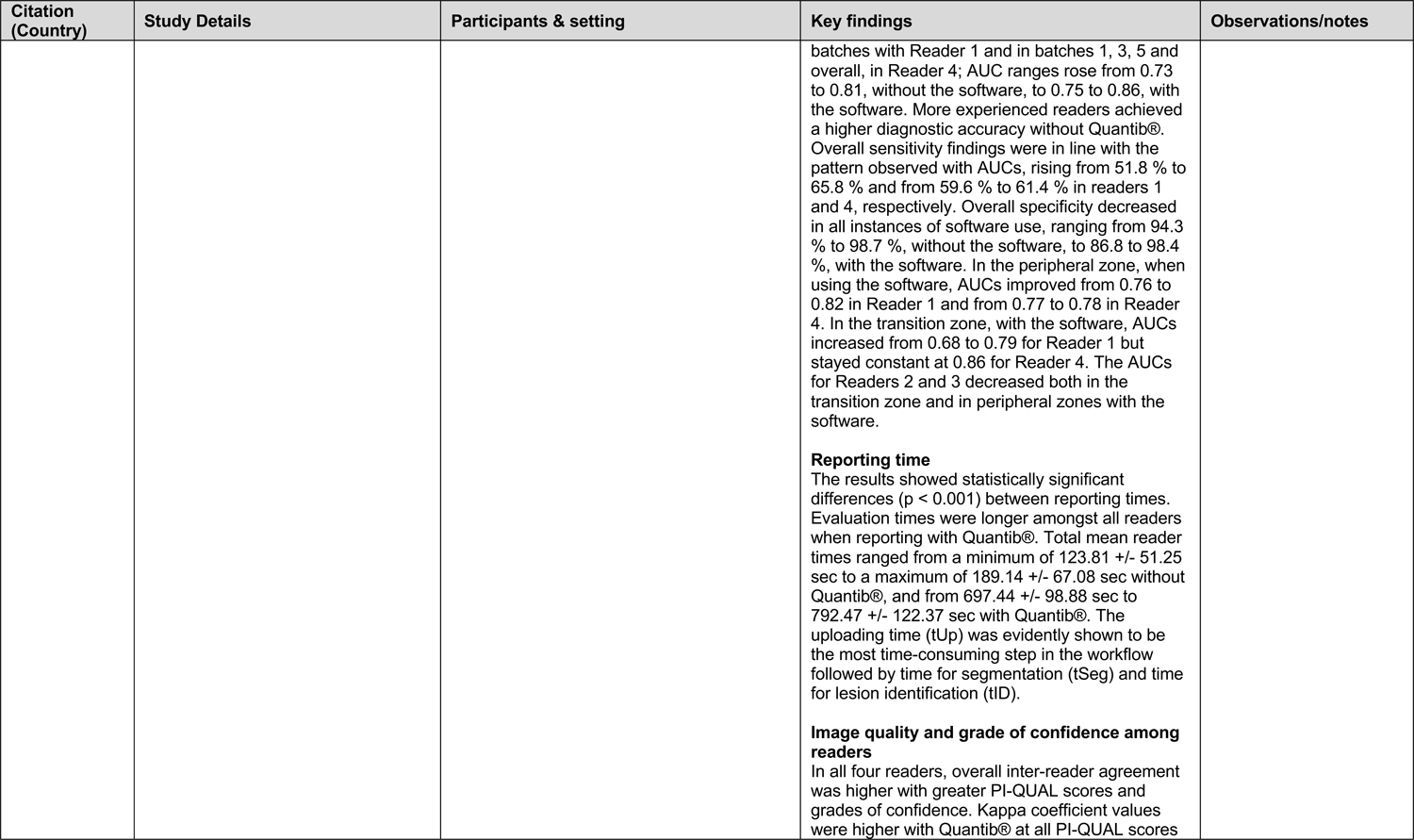
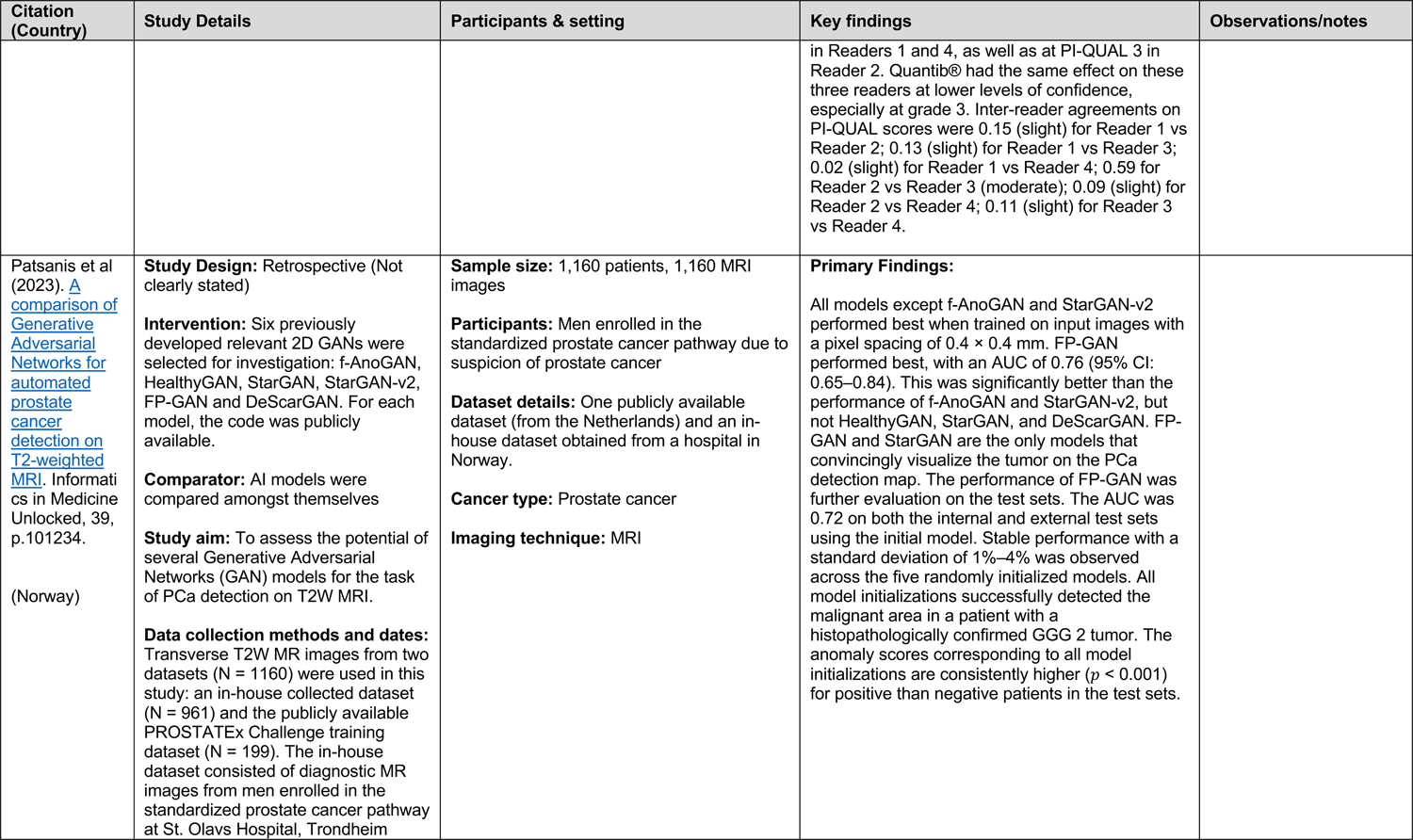
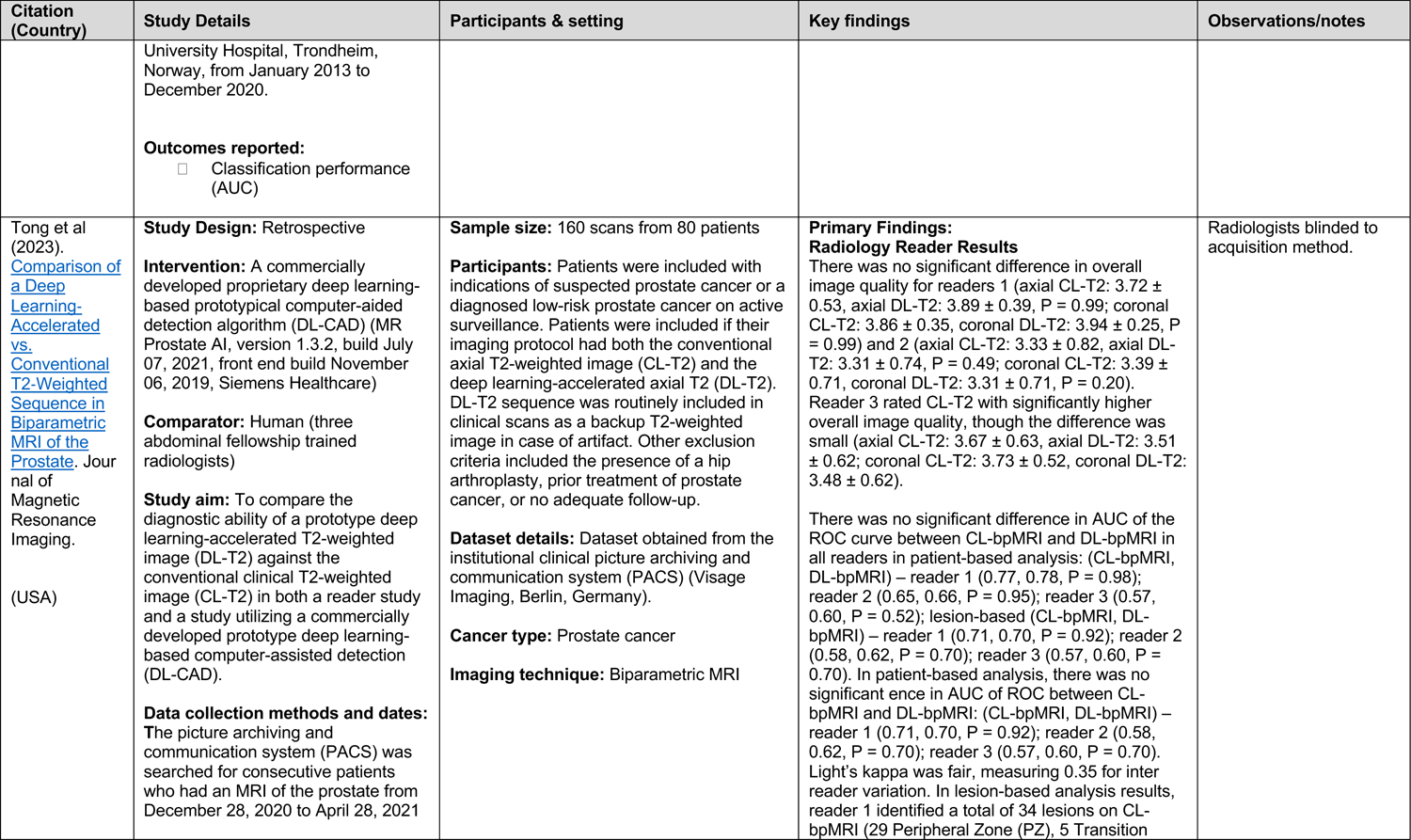
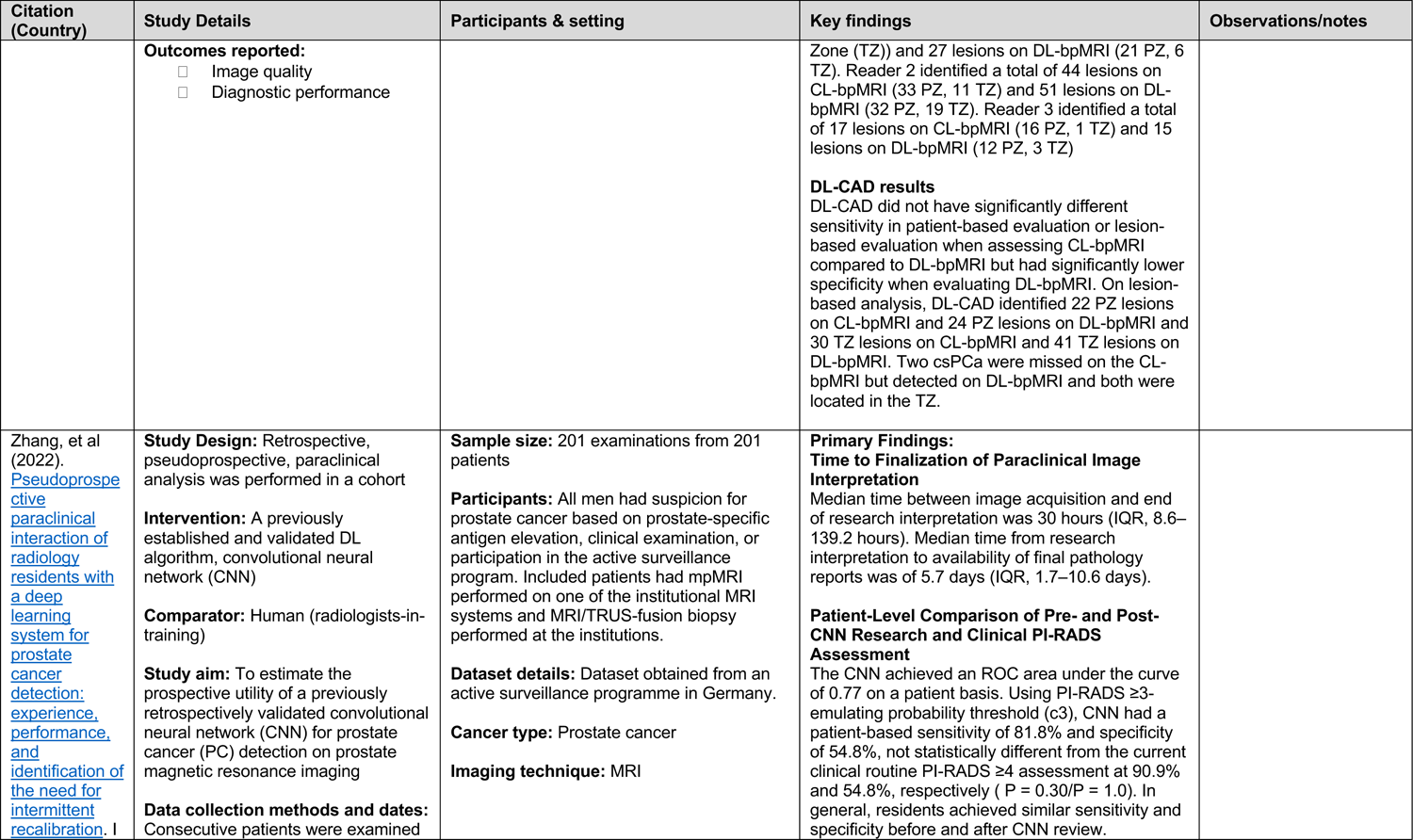
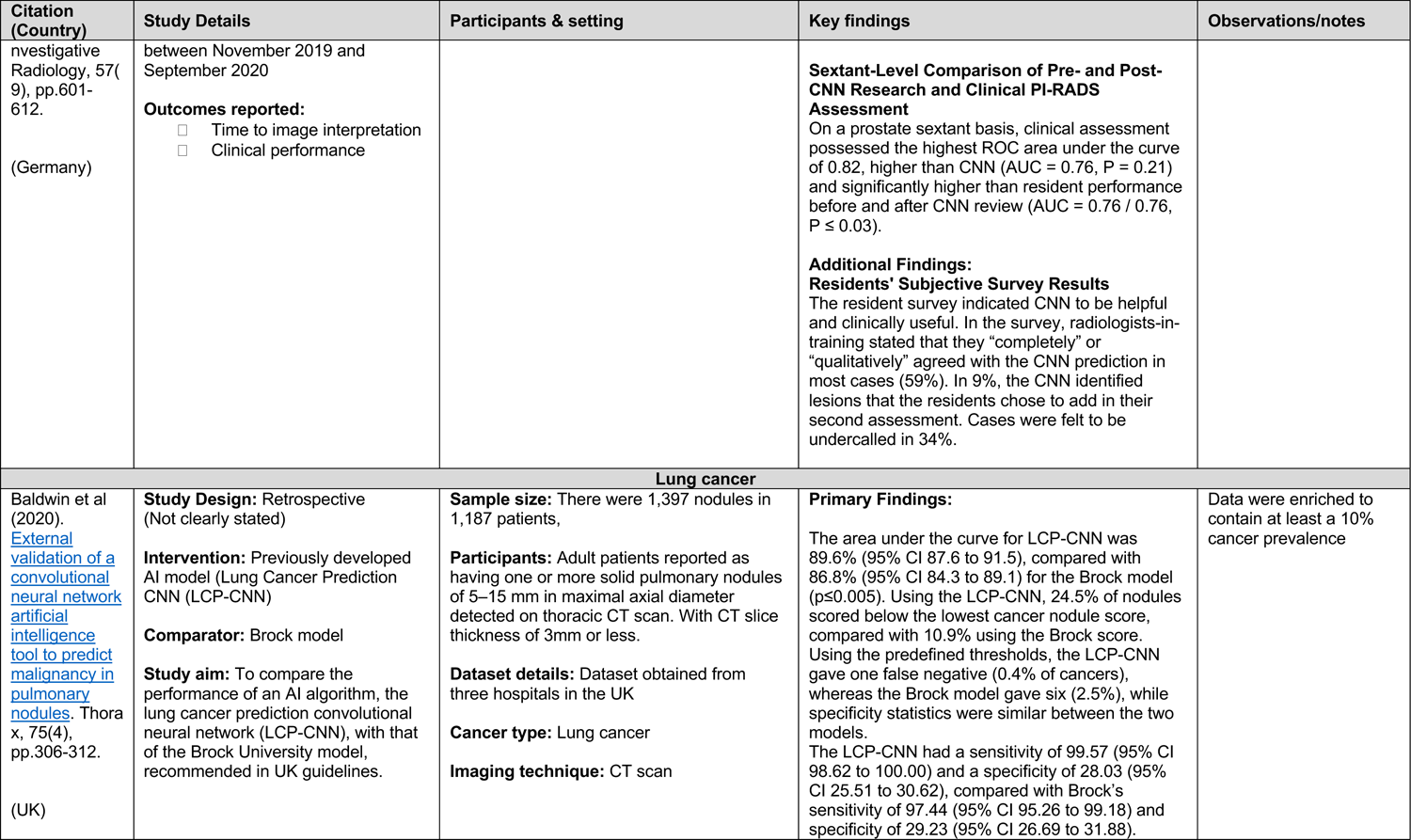
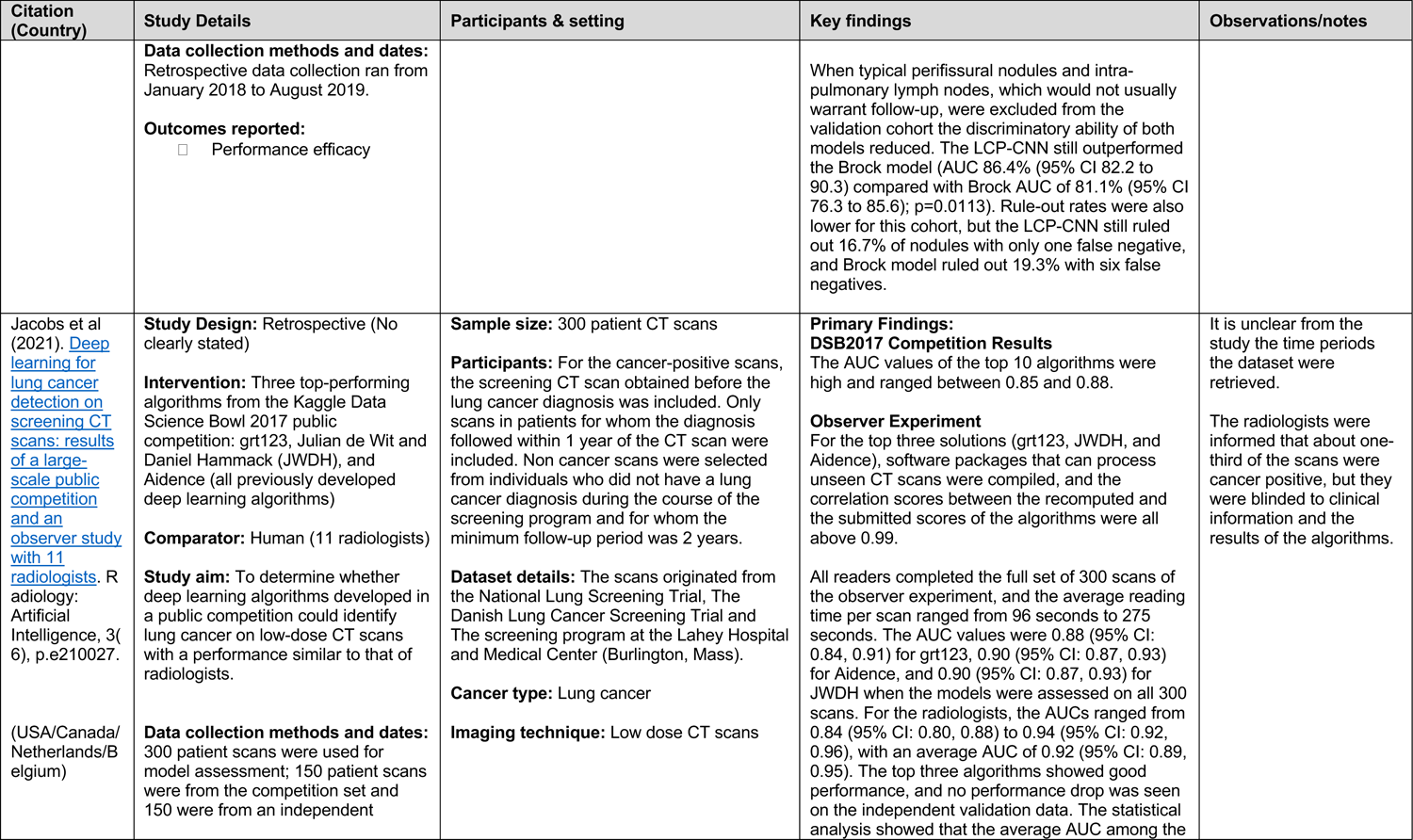
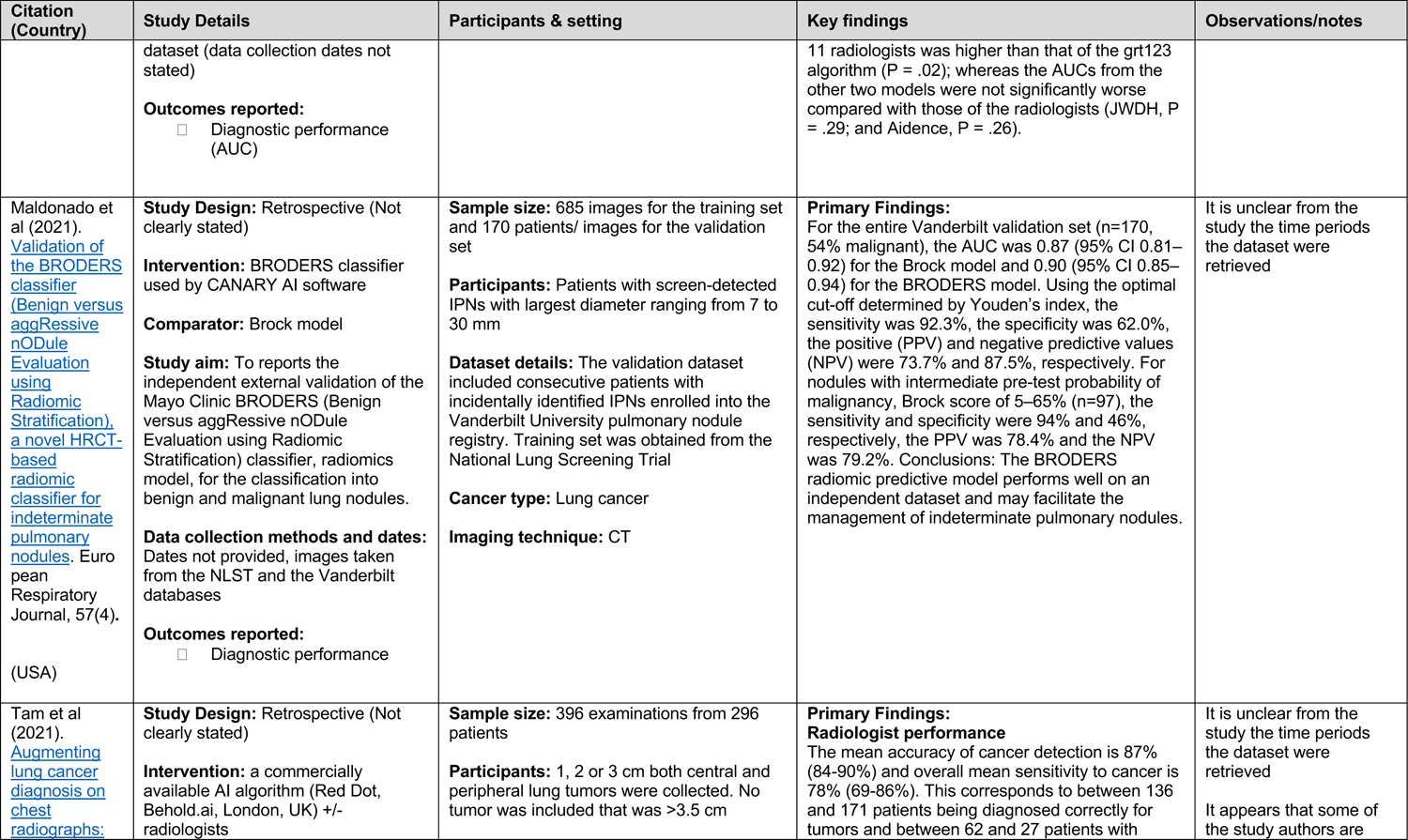
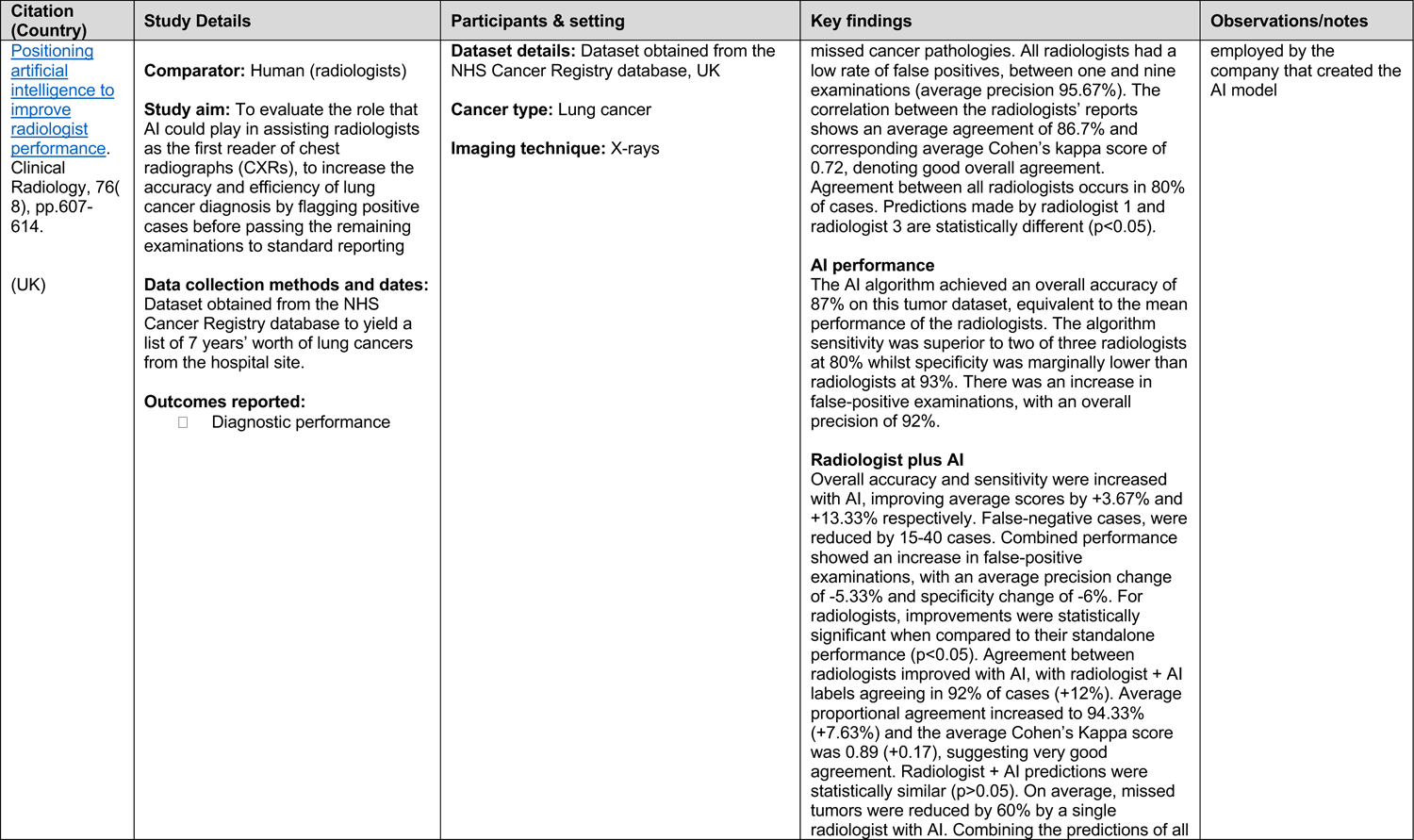
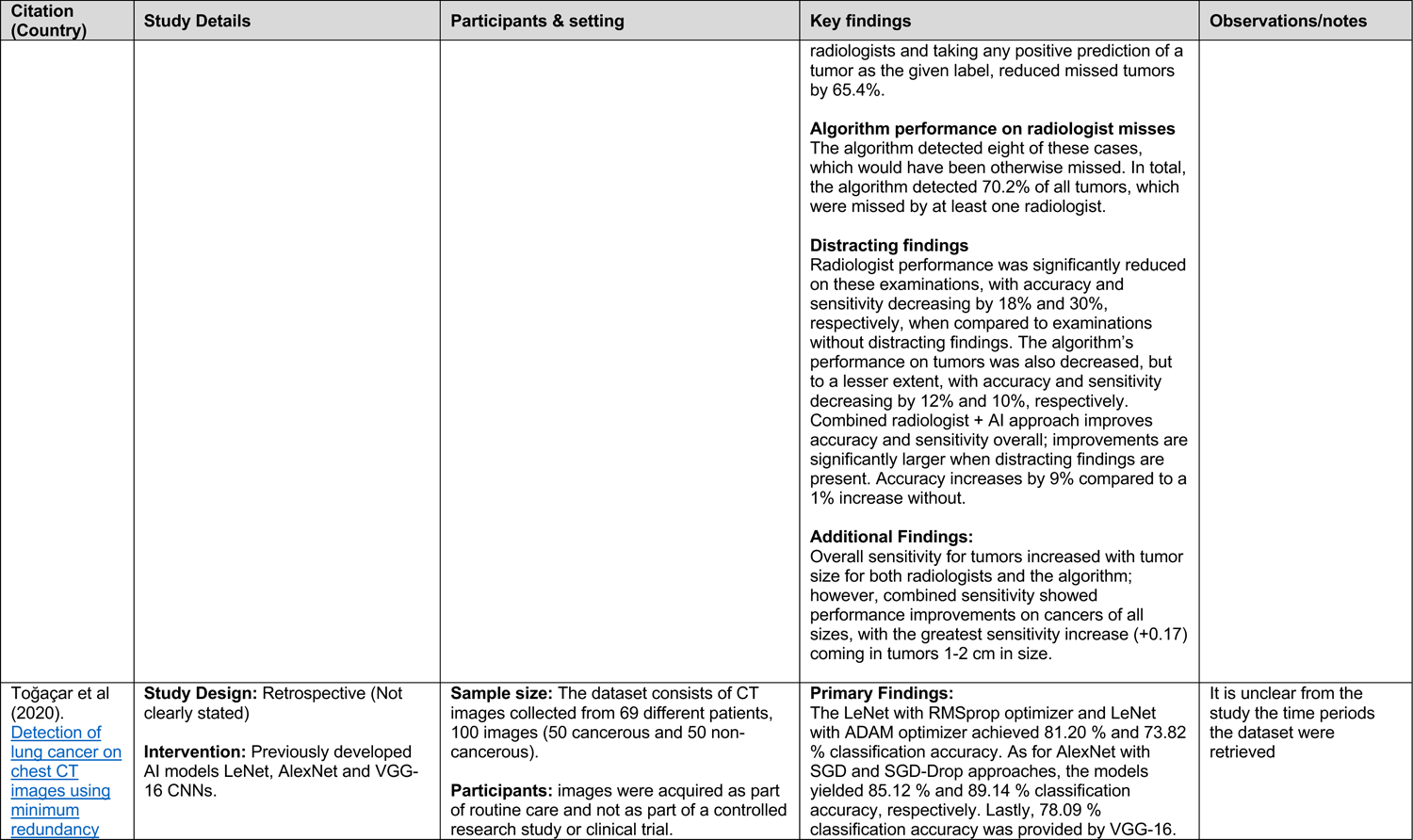
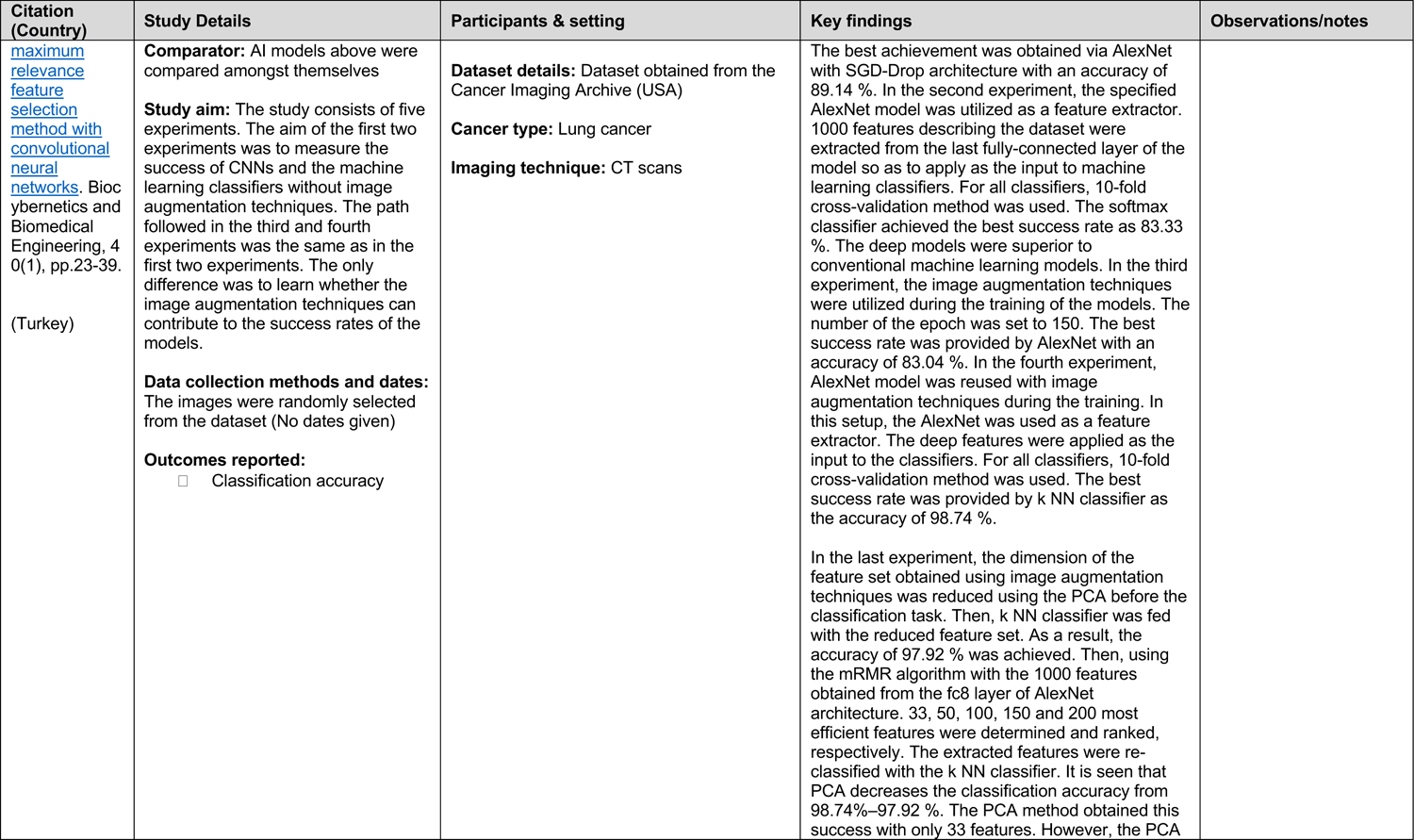
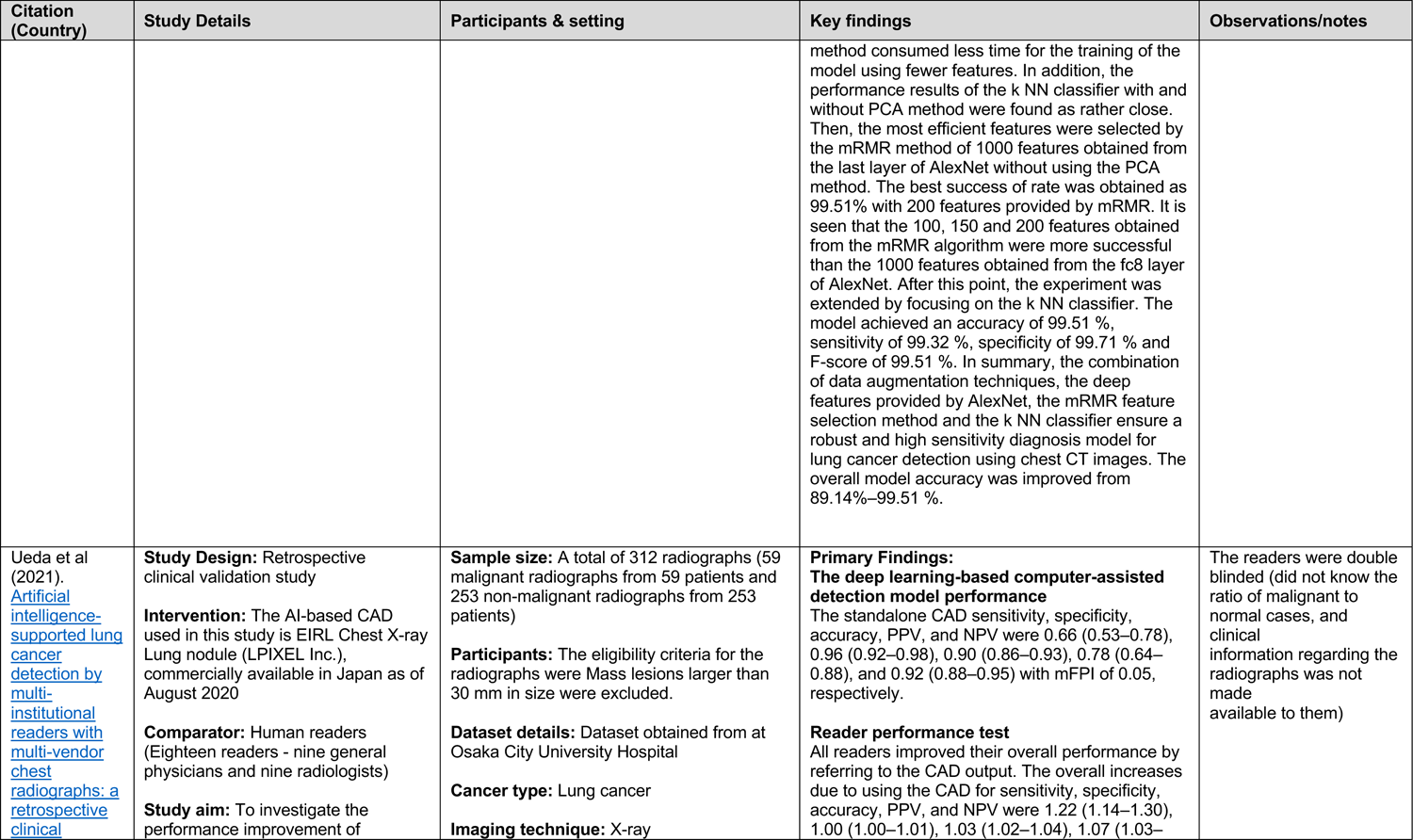
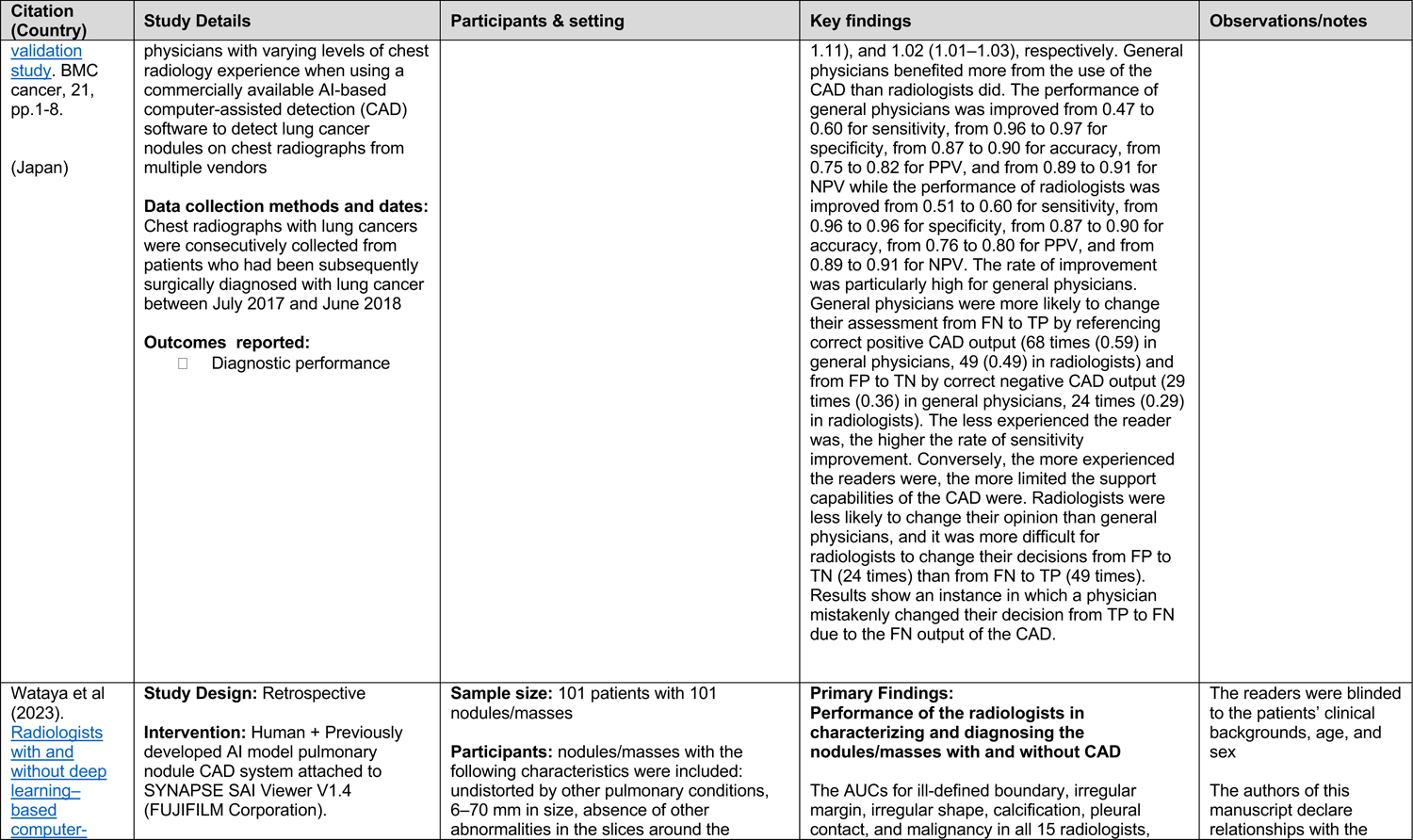
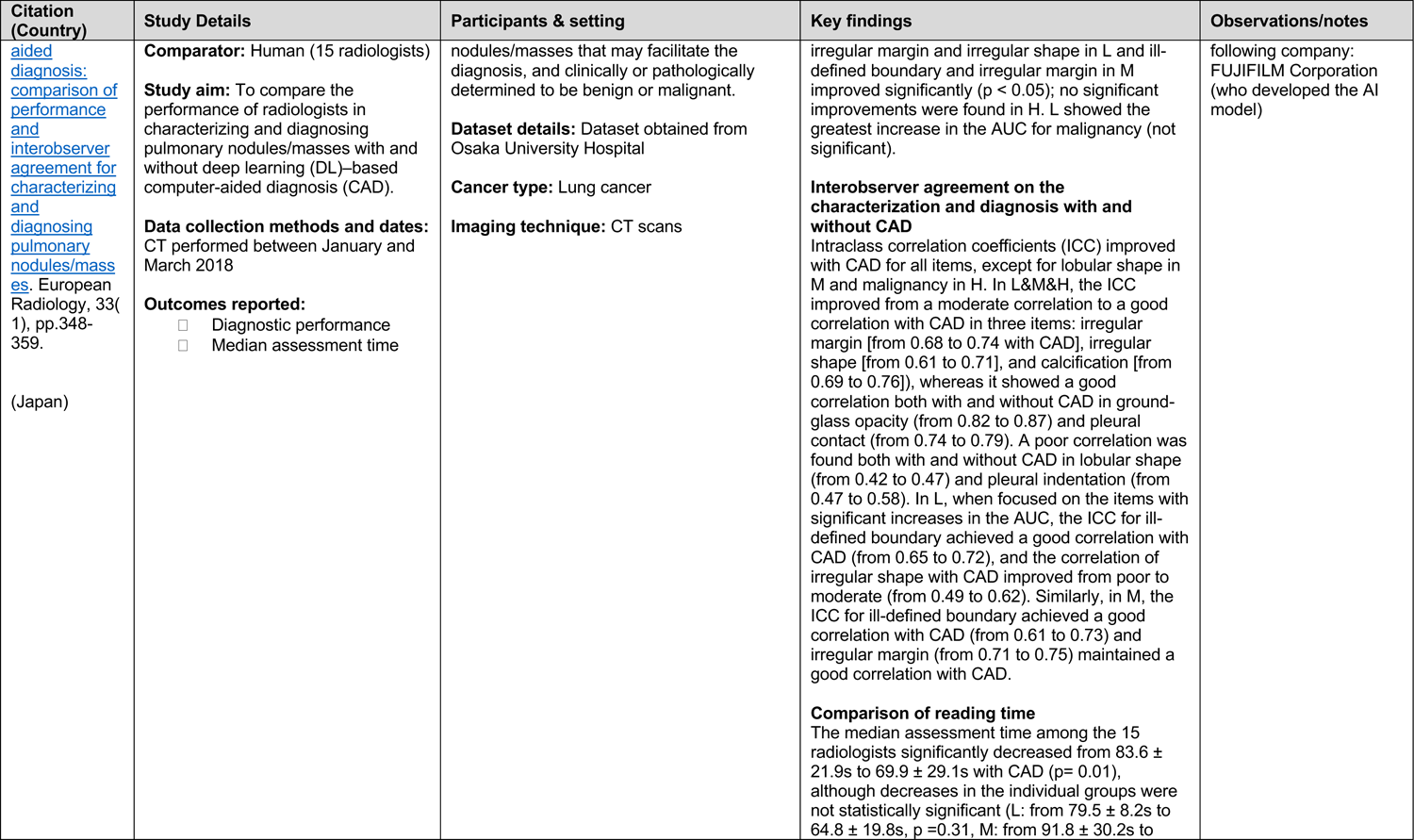
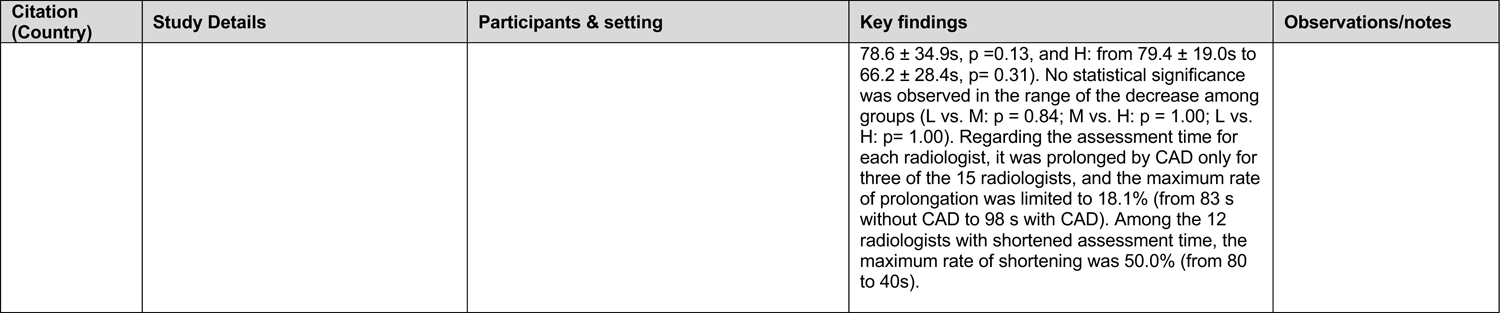
Summary of included studies.

### 7.3 Quality appraisal

**Table 7.**
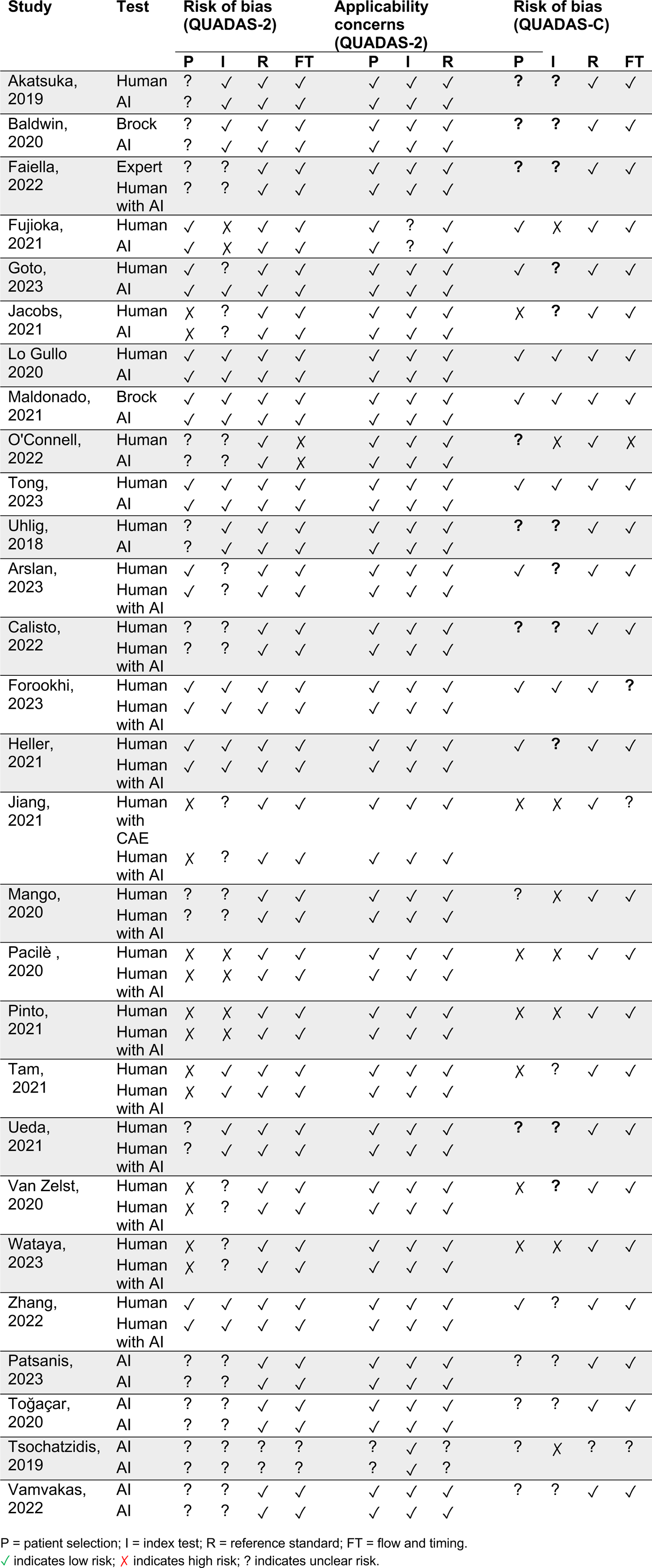
Quality appraisal results.

### 7.4 Information available on request

The following are available on request: protocol; search strategies for Embase, CENTRAL and ScanMedicine.

## Acknowledgements

The Public Health Wales team would like to thank James Triscott, Gareth Ashman, Delyth James, Josie Jackson, Rebecca Andrews, Christopher Rolls, David Jarrom, Anthony Cope, and Robert Hall for their time, expertise and contributions during stakeholder meetings in guiding the focus of the review and interpretation of findings.

## Abbreviations

Acronym: Full Description

AI: Artificial intelligence

AIS: Artificially Intelligent Systems

AUC: Area Under The Curve

BI-RADS: Breast Imaging Reporting and Data System

bpMRI: Biparametric Magnetic Resonance Imaging

BPN: Back Propagation Neural Networks

CAD: Computer Aided Diagnosis

CANARY: Computer-Aided Nodule Assessment and Risk Yield

CBCT: Cone-beam Computed Tomography

CE: The Conformité Européene

CI: Confidence Interval

CL-bpMRI: Conventional Biparametric Magnetic Resonance Imaging

CNN: Convolutional Neural Network

CQC: Care Quality Commission

CsPCa: Clinically significant prostate cancer

CT: Computed Tomography

DBT: Digital Breast Tomosynthesis

DCE: MRI Dynamic Contrast Material–Enhanced Magnetic Resonance Imaging

DCNN: Deep Convolutional Neural Network

DL: Deep learning

DLCAD: Deep Learning Computer Aided Diagnosis Software

DLCNN: Deep Learning Convolutional Neural Network

DL-bpMRI: Deep Learning-Accelerated Biparametric Magnetic Resonance Imaging

DNN: Deep Neural Network

DRE: Digital Rectal Examination

FDA: The United States Food and Drug Administration

GAN: Generative Adversarial Networks

kNN: k-Nearest Neighbour

LCP-CNN: Lung Cancer Prediction Convolutional Neural Network

ML: Machine Learning

MRI: Magnetic Resonance Imaging

mpMRI: Multiparametric Magnetic Resonance Imaging

mRMR: Minimum Redundancy Maximum Relevance

NHS: National Health Service

NICE: The National Institute for Health and Care Excellence

NPV: Negative Predictive Value

PCa: Prostate cancer

PI-QUAL: Prostate Imaging Quality

PI-RADS: Prostate Imaging Reporting & Data System

PPV: Positive Predictive Value

PSA: Prostate-Specific Antigen

ROC: Receiver Operating Characteristics

ROC: AUC Area Under The Receiver Operating Characteristic curve

ROI: Region of Interest

RR: Rapid Review

SD: Standard Deviation

SVM: Support Vector Machine

UK: United Kingdom

USA: United States of America

## APPENDIX

### APPENDIX 1: Reference list for studies included in the map

#### Breast

1. Adachi, M., Fujioka, T., Mori, M., Kubota, K., Kikuchi, Y., Xiaotong, W., Oyama, J., Kimura, K., Oda, G., Nakagawa, T., Uetake, H. and Tateishi, U. (2020) ‘Detection and Diagnosis of Breast Cancer Using Artificial Intelligence Based assessment of Maximum Intensity Projection Dynamic Contrast-Enhanced Magnetic Resonance Images’, Diagnostics, 10(5), pp. 20.
2. Barinov, L., Jairaj, A., Becker, M., Seymour, S., Lee, E., Schram, A., Lane, E., Goldszal, A., Quigley, D. and Paster, L. (2019) ‘Impact of Data Presentation on Physician Performance Utilizing Artificial Intelligence-Based Computer-Aided Diagnosis and Decision Support Systems’, Journal of Digital Imaging, 32(3), pp. 408-416.
3. Bhowmik, A., Monga, N., Belen, K., Varela, K., Sevilimedu, V., Thakur, S. B., Martinez, D. F., Sutton, E. J., Pinker, K. and Eskreis-Winkler, S. (2023) ‘Automated Triage of Screening Breast MRI Examinations in High-Risk Women Using an Ensemble Deep Learning Model’, Investigative radiology., 12.
4. Caballo, M., Hernandez, A. M., Lyu, S. H., Teuwen, J., Mann, R. M., van Ginneken, B., Boone, J. M. and Sechopoulos, I. (2021) ‘Computer-aided diagnosis of masses in breast computed tomography imaging: deep learning model with combined handcrafted and convolutional radiomic features’, Journal of Medical Imaging, 8(2), pp. 024501.
5. Calisto, F. M., Santiago, C., Nunes, N. and Nascimento, J. C. (2022) ‘BreastScreening-AI: Evaluating medical intelligent agents for human-AI interactions’, Artificial Intelligence in Medicine, 127.
6. Fujioka, T., Katsuta, L., Kubota, K., Mori, M., Kikuchi, Y., Kato, A., Oda, G., Nakagawa, T., Kitazume, Y. and Tateishi, U. (2020) ‘Classification of Breast Masses on Ultrasound Shear Wave Elastography using Convolutional Neural Networks’, Ultrasonic Imaging, 42(4), pp. 213-220.
7. Fujioka, T., Kubota, K., Mori, M., Kikuchi, Y., Katsuta, L., Kasahara, M., Oda, G., Ishiba, T., Nakagawa, T. and Tateishi, U. (2019) ‘Distinction between benign and malignant breast masses at breast ultrasound using deep learning method with convolutional neural network’, Japanese Journal of Radiology, 37(6), pp. 466-472.
8. Fujioka, T., Yashima, Y., Oyama, J., Mori, M., Kubota, K., Katsuta, L., Kimura, K., Yamaga, E., Oda, G., Nakagawa, T., Kitazume, Y. and Tateishi, U. (2021) ‘Deep-learning approach with convolutional neural network for classification of maximum intensity projections of dynamic contrast-enhanced breast magnetic resonance imaging’, Magnetic Resonance Imaging, 75, pp. 1-8.
9. Gheflati, B. and Rivaz, H. (2022) ‘Vision Transformers for Classification of Breast Ultrasound Images’, Annual International Conference Of The IEEE Engineering In Medicine And Biology Society, 2022, pp. 480-483.
10. Goto, M., Sakai, K., Toyama, Y., Nakai, Y. and Yamada, K. (2023) ‘Use of a deep learning algorithm for non-mass enhancement on breast MRI: comparison with radiologists’ interpretations at various levels’, Japanese Journal of Radiology, 18, pp. 18.
11. Hayashida, T., Odani, E., Kikuchi, M., Nagayama, A., Seki, T., Takahashi, M., Futatsugi, N., Matsumoto, A., Murata, T., Watanuki, R., Yokoe, T., Nakashoji, A., Maeda, H., Onishi, T., Asaga, S., Hojo, T., Jinno, H., Sotome, K., Matsui, A., Suto, A., Imoto, S. and Kitagawa, Y. (2022) ‘Establishment of a deep-learning system to diagnose BI-RADS4a or higher using breast ultrasound for clinical application’, Cancer Science, 113(10), pp. 3528-3534.
12. Hejduk, P., Marcon, M., Unkelbach, J., Ciritsis, A., Rossi, C., Borkowski, K. and Boss, A. (2022) ‘Fully automatic classification of automated breast ultrasound (ABUS) imaging according to BI-RADS using a deep convolutional neural network’, European Radiology, 32(7), pp. 4868-4878.
13. Heller, S. L., Wegener, M., Babb, J. S. and Gao, Y. (2020) ‘Can an Artificial Intelligence Decision Aid Decrease False-Positive Breast Biopsies?’, Ultrasound Quarterly, 37(1), pp. 10-15.
14. Hoffmann, R., Reich, C. and Skerl, K. (2022) ‘Evaluating different combination methods to analyse ultrasound and shear wave elastography images automatically through discriminative convolutional neural network in breast cancer imaging’, International Journal of Computer Assisted Radiology & Surgery, 17(12), pp. 2231-2237.
15. Interlenghi, M., Salvatore, C., Magni, V., Caldara, G., Schiavon, E., Cozzi, A., Schiaffino, S., Carbonaro, L. A., Castiglioni, I. and Sardanelli, F. (2022) ‘A Machine Learning Ensemble Based on Radiomics to Predict BI-RADS Category and Reduce the Biopsy Rate of Ultrasound-Detected Suspicious Breast Masses’, Diagnostics, 12.
16. Jiang, Y., Edwards, A. V. and Newstead, G. M. (2021) ‘Artificial Intelligence Applied to Breast MRI for Improved Diagnosis’, Radiology, 298(1), pp. 38-46.
17. Jing, X., Dorrius, M. D., Wielema, M., Sijens, P. E., Oudkerk, M. and van Ooijen, P. (2022) ‘Breast Tumor Identification in Ultrafast MRI Using Temporal and Spatial Information’, Cancers, 14.
18. Lo Gullo, R., Daimiel, I., Rossi Saccarelli, C., Bitencourt, A., Gibbs, P., Fox, M. J., Thakur, S. B., Martinez, D. F., Jochelson, M. S., Morris, E. A. and Pinker, K. (2020) ‘Improved characterization of sub-centimeter enhancing breast masses on MRI with radiomics and machine learning in BRCA mutation carriers’, European Radiology, 30(12), pp. 6721-6731.
19. Makrogiannis, S., Zheng, K. and Harris, C. (2021) ‘Discriminative Localized Sparse Approximations for Mass Characterization in Mammograms’, Frontiers in Oncology, 11, pp. 725320.
20. Mango, V. L., Sun, M., Wynn, R. T. and Ha, R. (2020) ‘Should We Ignore, Follow, or Biopsy? Impact of Artificial Intelligence Decision Support on Breast Ultrasound Lesion Assessment’, AJR. American Journal of Roentgenology, 214(6), pp. 1445-1452.
21. Naranjo, I. D., Gibbs, P., Reiner, J. S., Gullo, R. L., Thakur, S. B., Jochelson,
22. S., Thakur, N., Baltzer, P. A. T., Helbich, T. H. and Pinker, K. (2022) ‘Breast Lesion Classification with Multiparametric Breast MRI Using Radiomics and Machine Learning: A Comparison with Radiologists’ Performance’, Cancers, 14.
23. O’Connell, A. M., Bartolotta, T. V., Orlando, A., Jung, S. H., Baek, J. and Parker, K. J. (2022) ‘Diagnostic Performance of an Artificial Intelligence System in Breast Ultrasound’, Journal of Ultrasound in Medicine, 41(1), pp. 97-105.
24. Pacile, S., Lopez, J., Chone, P., Bertinotti, T., Grouin, J. M. and Fillard, P. (2020) ‘Improving Breast Cancer Detection Accuracy of Mammography with the Concurrent Use of an Artificial Intelligence Tool’, Radiology Artificial intelligence, 2(6), pp. e190208.
25. Patel, B. K., Ranjbar, S., Wu, T., Pockaj, B. A., Li, J., Zhang, N., Lobbes, M., Zhang, B. and Mitchell, J. R. (2018) ‘Computer-aided diagnosis of contrast-enhanced spectral mammography: A feasibility study’, European Journal of Radiology, 98, pp. 207-213.
26. Pfob, A., Sidey-Gibbons, C., Barr, R. G., Duda, V., Alwafai, Z., Balleyguier, C., Clevert, D. A., Fastner, S., Gomez, C., Goncalo, M., Gruber, I., Hahn, M., Hennigs, A., Kapetas, P., Lu, S. C., Nees, J., Ohlinger, R., Riedel, F., Rutten, M., Schaefgen, B., Schuessler, M., Stieber, A., Togawa, R., Tozaki, M., Wojcinski, S., Xu, C., Rauch, G., Heil, J. and Golatta, M. (2022) ‘The importance of multi-modal imaging and clinical information for humans and AI-based algorithms to classify breast masses (INSPiRED 003): an international, multicenter analysis’, European Radiology, 32(6), pp. 4101-4115.
27. Pinto, M. C., Rodriguez-Ruiz, A., Pedersen, K., Hofvind, S., Wicklein, J., Kappler, S., Mann, R. M. and Sechopoulos, I. (2021) ‘Impact of Artificial Intelligence Decision Support Using Deep Learning on Breast Cancer Screening Interpretation with Single-View Wide-Angle Digital Breast Tomosynthesis’, Radiology, 300(3), pp. 529-536.
28. Romeo, V., Clauser, P., Rasul, S., Kapetas, P., Gibbs, P., Baltzer, P. A. T., Hacker, M., Woitek, R., Helbich, T. H. and Pinker, K. (2022) ‘AI-enhanced simultaneous multiparametric 18F-FDG PET/MRI for accurate breast cancer diagnosis’, European Journal of Nuclear Medicine & Molecular Imaging, 49(2), pp. 596-608.
29. Romeo, V., Cuocolo, R., Apolito, R., Stanzione, A., Ventimiglia, A., Vitale, A., Verde, F., Accurso, A., Amitrano, M., Insabato, L., Gencarelli, A., Buonocore, R., Argenzio, M. R., Cascone, A. M., Imbriaco, M., Maurea, S. and Brunetti, A. (2021) ‘Clinical value of radiomics and machine learning in breast ultrasound: a multicenter study for differential diagnosis of benign and malignant lesions’, European Radiology, 31(12), pp. 9511-9519.
30. Sabani, A., Landsmann, A., Hejduk, P., Schmidt, C., Marcon, M., Borkowski, K., Rossi, C., Ciritsis, A. and Boss, A. (2022) ‘BI-RADS-Based Classification of Mammographic Soft Tissue Opacities Using a Deep Convolutional Neural Network’, Diagnostics, 12(7), pp. 28.
31. Takahashi, K., Fujioka, T., Oyama, J., Mori, M., Yamaga, E., Yashima, Y., Imokawa, T., Hayashi, A., Kujiraoka, Y., Tsuchiya, J., Oda, G., Nakagawa, T. and Tateishi, U. (2022) ‘Deep Learning Using Multiple Degrees of Maximum-Intensity Projection for PET/CT Image Classification in Breast Cancer’, Tomography, 8(1), pp. 131-141.
32. Tanaka, H., Chiu, S. W., Watanabe, T., Kaoku, S. and Yamaguchi, T. (2019) ‘Computer-aided diagnosis system for breast ultrasound images using deep learning’, Physics in Medicine & Biology, 64(23), pp. 235013.
33. Toprak, A. (2018) ‘Extreme Learning Machine (ELM)-Based Classification of Benign and Malignant Cells in Breast Cancer’, Medical Science Monitor, 24, pp. 6537-6543.
34. Truhn, D., Schrading, S., Haarburger, C., Schneider, H., Merhof, D. and Kuhl, C. (2019) ‘Radiomic versus Convolutional Neural Networks Analysis for Classification of Contrast-enhancing Lesions at Multiparametric Breast MRI’, Radiology, 290(2), pp. 290-297.
35. Tsochatzidis, L., Costaridou, L. and Pratikakis, I. (2019) ‘Deep Learning for Breast Cancer Diagnosis from Mammograms-A Comparative Study’, Journal of Imaging, 5(3), pp. 13.
36. Uhlig, J., Uhlig, A., Kunze, M., Beissbarth, T., Fischer, U., Lotz, J. and Wienbeck, S. (2018) ‘Novel Breast Imaging and Machine Learning: Predicting Breast Lesion Malignancy at Cone-Beam CT Using Machine Learning Techniques’, AJR. American Journal of Roentgenology, 211(2), pp. W123-W131.
37. Vamvakas, A., Tsivaka, D., Logothetis, A., Vassiou, K. and Tsougos, I. (2022) ‘Breast Cancer Classification on Multiparametric MRI - Increased Performance of Boosting Ensemble Methods’, Technology in Cancer Research & Treatment, 21, pp. 15330338221087828.
38. van Zelst, J. C., Tan, T., Mann, R. M. and Karssemeijer, N. (2020) ‘Validation of radiologists’ findings by computer-aided detection (CAD) software in breast cancer detection with automated 3D breast ultrasound: a concept study in implementation of artificial intelligence software’, Acta Radiologica, 61(3), pp. 312-320.
39. Wang, K., Patel, B. K., Wang, L., Wu, T., Zheng, B. and Li, J. (2019) ‘A dual-mode deep transfer learning (D2TL) system for breast cancer detection using contrast enhanced digital mammograms’, IISE Transactions on Healthcare Systems Engineering, 9, pp. 357-370.
40. Wu, N., Phang, J., Park, J., Shen, Y., Huang, Z., Zorin, M., Jastrzebski, S., Fevry, T., Katsnelson, J., Kim, E., Wolfson, S., Parikh, U., Gaddam, S., Lin, L. L. Y., Ho, K., Weinstein, J. D., Reig, B., Gao, Y., Toth, H., Pysarenko, K., Lewin, A., Lee, J., Airola, K., Mema, E., Chung, S., Hwang, E., Samreen, N., Kim, S. G., Heacock, L., Moy, L., Cho, K. and Geras, K. J. (2020) ‘Deep Neural Networks Improve Radiologists’ Performance in Breast Cancer Screening’, IEEE Transactions on Medical Imaging, 39(4), pp. 1184-1194.
41. Yang, K., Suzuki, A., Ye, J., Nosato, H., Izumori, A. and Sakanashi, H. (2022) ‘CTG-Net: Cross-task guided network for breast ultrasound diagnosis’, PLoS ONE [Electronic Resource], 17(8), pp. e0271106.

#### Gynaecological

1. Christiansen, F., Epstein, E. L., Smedberg, E., Akerlund, M., Smith, K. and Epstein, E. (2021) ‘Ultrasound image analysis using deep neural networks for discriminating between benign and malignant ovarian tumors: comparison with expert subjective assessment’, *Ultrasound in Obstetrics & Gynecology,* 57(1), pp. 155-163.
2. Nakagawa, M., Nakaura, T., Namimoto, T., Iyama, Y., Kidoh, M., Hirata, K., Nagayama, Y., Oda, S., Sakamoto, F., Shiraishi, S. and Yamashita, Y. (2019) ‘A multiparametric MRI-based machine learning to distinguish between uterine sarcoma and benign leiomyoma: comparison with 18F-FDG PET/CT’, *Clinical Radiology,* 74(2), pp. 167.e1-167.e7.
3. Nakagawa, M., Nakaura, T., Namimoto, T., Iyama, Y., Kidoh, M., Hirata, K., Nagayama, Y., Yuki, H., Oda, S., Utsunomiya, D. and Yamashita, Y. (2019) ‘Machine Learning to Differentiate T2-Weighted Hyperintense Uterine Leiomyomas from Uterine Sarcomas by Utilizing Multiparametric Magnetic Resonance Quantitative Imaging Features’, *Academic Radiology,* 26(10), pp. 1390-1399.
4. Saida, T., Mori, K., Hoshiai, S., Sakai, M., Urushibara, A., Ishiguro, T., Minami, M., Satoh, T. and Nakajima, T. (2022) ‘Diagnosing Ovarian Cancer on MRI: A Preliminary Study Comparing Deep Learning and Radiologist Assessments’, *Cancers,* 14(4), pp. 16.
5. Toyohara, Y., Sone, K., Noda, K., Yoshida, K., Kurokawa, R., Tanishima, T., Kato, S., Inui, S., Nakai, Y., Ishida, M., Gonoi, W., Tanimoto, S., Takahashi, Y., Inoue, F., Kukita, A., Kawata, Y., Taguchi, A., Furusawa, A., Miyamoto, Y., Tsukazaki, T., Tanikawa, M., Iriyama, T., Mori-Uchino, M., Tsuruga, T., Oda, K., Yasugi, T., Takechi, K., Abe, O. and Osuga, Y. (2022) ‘Development of a deep learning method for improving diagnostic accuracy for uterine sarcoma cases’, *Scientific Reports,* 12(1), pp. 19612.
6. Urushibara, A., Saida, T., Mori, K., Ishiguro, T., Inoue, K., Masumoto, T., Satoh, T. and Nakajima, T. (2022) ‘The efficacy of deep learning models in the diagnosis of endometrial cancer using MRI: a comparison with radiologists’, *BMC Medical Imaging,* 22(1), pp. 80.
7. Urushibara, A., Saida, T., Mori, K., Ishiguro, T., Sakai, M., Masuoka, S., Satoh, T. and Masumoto, T. (2021) ‘Diagnosing uterine cervical cancer on a single T2-weighted image: Comparison between deep learning versus radiologists’, *European Journal of Radiology,* 135.

#### Lung

1. Astaraki, M., Yang, G., Zakko, Y., Toma-Dasu, I., Smedby, O. and Wang, C. (2021) ‘A Comparative Study of Radiomics and Deep-Learning Based Methods for Pulmonary Nodule Malignancy Prediction in Low Dose CT Images’, Frontiers in Oncology, 11.
2. Baldwin, D. R., Gustafson, J., Pickup, L., Arteta, C., Novotny, P., Declerck, J., Kadir, T., Figueiras, C., Sterba, A., Exell, A., Potesil, V., Holland, P., Spence, H., Clubley, A., O’Dowd, E., Clark, M., Ashford-Turner, V., Callister, M. E. and Gleeson, F. V. (2020) ‘External validation of a convolutional neural network artificial intelligence tool to predict malignancy in pulmonary nodules’, Thorax., 5.
3. Gursoy Coruh, A., Yenigun, B., Uzun, C., Kahya, Y., Buyukceran, E. U., Elhan, A., Orhan, K. and Kayi Cangir, A. (2021) ‘A comparison of the fusion model of deep learning neural networks with human observation for lung nodule detection and classification’, British Journal of Radiology, 94(1123), pp. 20210222.
4. Hunter, B., Chen, M., Ratnakumar, P., Alemu, E., Logan, A., Linton-Reid, K., Tong, D., Senthivel, N., Bhamani, A., Bloch, S., Kemp, S. V., Boddy, L., Jain, S., Gareeboo, S., Rawal, B., Doran, S., Navani, N., Nair, A., Bunce, C., Kaye, S., Blackledge, M., Aboagye, E. O., Devaraj, A. and Lee, R. W. (2022) ‘A radiomics-based decision support tool improves lung cancer diagnosis in combination with the Herder score in large lung nodules’, EBioMedicine, 86, pp. 104344.
5. Jacobs, C., Setio, A. A. A., Scholten, E. T., Gerke, P. K., Bhattacharya, H., Hoesein, F. A. M., Brink, M., Ranschaert, E., de Jong, P. A., Silva, M., Geurts, B., Chung, K., Schalekamp, S., Meersschaert, J., Devaraj, A., Pinsky, P. F., Lam, S. C., van Ginneken, B. and Farahani, K. (2021) ‘Deep learning for lung cancer detection on screening ct scans: Results of a large-scale public competition and an observer study with 11 radiologists’, Radiology: Artificial Intelligence, 3.
6. Kitajima, K., Matsuo, H., Kono, A., Kuribayashi, K., Kijima, T., Hashimoto, M., Hasegawa, S., Murakami, T. and Yamakado, K. (2021) ‘Deep learning with deep convolutional neural network using FDG-PET/CT for malignant pleural mesothelioma diagnosis’, Oncotarget, 12(12), pp. 1187-1196.
7. Maldonado, F., Varghese, C., Rajagopalan, S., Duan, F., Balar, A. B., Lakhani, D. A., Antic, S. L., Massion, P. P., Johnson, T. F., Karwoski, R. A., Robb, R. A., Bartholmai, B. J. and Peikert, T. (2021) ‘Validation of the BRODERS classifier (Benign versus aggRessive nODule Evaluation using Radiomic Stratification), a novel HRCT-based radiomic classifier for indeterminate pulmonary nodules’, European Respiratory Journal, 57(4), pp. 04.
8. Nishio, M., Nishizawa, M., Sugiyama, O., Kojima, R., Yakami, M., Kuroda, T. and Togashi, K. (2018) ‘Computer-aided diagnosis of lung nodule using gradient tree boosting and Bayesian optimization’, PLoS ONE [Electronic Resource], 13(4), pp. e0195875.
9. Sollini, M., Kirienko, M., Gozzi, N., Bruno, A., Torrisi, C., Balzarini, L., Voulaz, E., Alloisio, M. and Chiti, A. (2023) ‘The Development of an Intelligent Agent to Detect and Non-Invasively Characterize Lung Lesions on CT Scans: Ready for the “Real World”?’, Cancers, 15(2), pp. 05.
10. Tam, M., Dyer, T., Dissez, G., Morgan, T. N., Hughes, M., Illes, J., Rasalingham, R. and Rasalingham, S. (2021) ‘Augmenting lung cancer diagnosis on chest radiographs: positioning artificial intelligence to improve radiologist performance’, Clinical Radiology, 76(8), pp. 607-614.
11. Togacar, M., Ergen, B. and Comert, Z. (2020) ‘Detection of lung cancer on chest CT images using minimum redundancy maximum relevance feature selection method with convolutional neural networks’, Biocybernetics and Biomedical Engineering, 40, pp. 23-39.
12. Ueda, D., Yamamoto, A., Shimazaki, A., Walston, S. L., Matsumoto, T., Izumi, N., Tsukioka, T., Komatsu, H., Inoue, H., Kabata, D., Nishiyama, N. and Miki, Y. (2021) ‘Artificial intelligence-supported lung cancer detection by multi-institutional readers with multi-vendor chest radiographs: a retrospective clinical validation study’, BMC Cancer, 21(1), pp. 1120.
13. Wataya, T., Yanagawa, M., Tsubamoto, M., Sato, T., Nishigaki, D., Kita, K., Yamagata, K., Suzuki, Y., Hata, A., Kido, S. and Tomiyama, N. (2023) ‘Radiologists with and without deep learning-based computer-aided diagnosis: comparison of performance and interobserver agreement for characterizing and diagnosing pulmonary nodules/masses’, European Radiology, 33, pp. 348-359.
14. Zhang, Q. and Yoon, S. (2022) ‘A novel self-adaptive convolutional neural network model using spatial pyramid pooling for 3D lung nodule computer-aided diagnosis’, IISE Transactions on Healthcare Systems Engineering, 12, pp. 75-88.

#### Prostate

1. Akatsuka, J., Yamamoto, Y., Sekine, T., Numata, Y., Morikawa, H., Tsutsumi, K., Yanagi, M., Endo, Y., Takeda, H., Hayashi, T., Ueki, M., Tamiya, G., Maeda, I., Fukumoto, M., Shimizu, A., Tsuzuki, T., Kimura, G. and Kondo, Y. (2019) ‘Illuminating Clues of Cancer Buried in Prostate MR Image: Deep Learning and Expert Approaches’, *Biomolecules,* 9(11), pp. 30.
2. Arslan, A., Alis, D., Erdemli, S., Seker, M. E., Zeybel, G., Sirolu, S., Kurtcan, S. and Karaarslan, E. (2023) ‘Does deep learning software improve the consistency and performance of radiologists with various levels of experience in assessing bi-parametric prostate MRI?’, *Insights into Imaging,* 14.
3. Bhattacharya, I., Seetharaman, A., Kunder, C., Shao, W., Chen, L. C., Soerensen, S. J. C., Wang, J. B., Teslovich, N. C., Fan, R. E., Ghanouni, P., Brooks, J. D., Sonn, G. A. and Rusu, M. (2022) ‘Selective identification and localization of indolent and aggressive prostate cancers via CorrSigNIA: an MRI-pathology correlation and deep learning framework’, *Medical Image Analysis,* 75, pp. 102288.
4. Bonekamp, D., Kohl, S., Wiesenfarth, M., Schelb, P., Radtke, J. P., Gotz, M., Kickingereder, P., Yaqubi, K., Hitthaler, B., Gahlert, N., Kuder, T. A., Deister, F., Freitag, M., Hohenfellner, M., Hadaschik, B. A., Schlemmer, H. P. and Maier-Hein, K. H. (2018) ‘Radiomic Machine Learning for Characterization of Prostate Lesions with MRI: Comparison to ADC Values’, *Radiology,* 289(1), pp. 128-137.
5. Faiella, E., Vertulli, D., Esperto, F., Cordelli, E., Soda, P., Muraca, R. M., Moramarco, L. P., Grasso, R. F., Beomonte Zobel, B. and Santucci, D. (2022) ‘Quantib Prostate Compared to an Expert Radiologist for the Diagnosis of Prostate Cancer on mpMRI: A Single-Center Preliminary Study’, *Tomography,* 8(4), pp. 2010-2019.
6. Forookhi, A., Laschena, L., Pecoraro, M., Borrelli, A., Massaro, M., Dehghanpour, A., Cipollari, S., Catalano, C. and Panebianco, V. (2023) ‘Bridging the experience gap in prostate multiparametric magnetic resonance imaging using artificial intelligence: A prospective multi-reader comparison study on inter-reader agreement in PI-RADS v2.1, image quality and reporting time between novice and expert readers’, *European Journal of Radiology,* 161, pp. 110749.
7. Gresser, E., Schachtner, B., Stuber, A. T., Solyanik, O., Schreier, A., Huber, T., Froelich, M. F., Magistro, G., Kretschmer, A., Stief, C., Ricke, J., Ingrisch, M. and Norenberg, D. (2022) ‘Performance variability of radiomics machine learning models for the detection of clinically significant prostate cancer in heterogeneous MRI datasets’, *Quantitative Imaging in Medicine and Surgery,* 12, pp. 4990-5003.
8. Hamm, C. A., Baumgartner, G. L., Biessmann, F., Beetz, N. L., Hartenstein, A., Savic, L. J., Frobose, K., Drager, F., Schallenberg, S., Rudolph, M., Baur, A. D. J., Hamm, B., Haas, M., Hofbauer, S., Cash, H. and Penzkofer, T. (2023) ‘Interactive Explainable Deep Learning Model Informs Prostate Cancer Diagnosis at MRI’, *Radiology,* 307(4), pp. e222276.
9. Hosseinzadeh, M., Saha, A., Brand, P., Slootweg, I., de Rooij, M. and Huisman, H. (2022) ‘Deep learning-assisted prostate cancer detection on bi-parametric MRI: minimum training data size requirements and effect of prior knowledge’, *European Radiology,* 32(4), pp. 2224-2234.
10. Patsanis, A., Sunoqrot, M. R. S., Langorgen, S., Wang, H., Selnaes, K. M., Bertilsson, H., Bathen, T. F. and Elcho, M. (2023) ‘A comparison of Generative Adversarial Networks for automated prostate cancer detection on T2-weighted MRI’, *Informatics in Medicine Unlocked,* 39.
11. Tong, A., Bagger, B., Patricelli, R., Amerika, P., VI, A., Qian, K., Grimm, R., Kaman, A., Keerthivasan, M. B., Nickel, M. D., von Busch, H. and Chandarana, H. (2023) ‘Comparison of a Deep Learning-Accelerated vs. Conventional T2-Weighted Sequence in Biparametric MRI of the Prostate’, *Journal of Magnetic Resonance Imaging*.
12. Zhang, K. S., Schelb, P., Netzer, N., Tavakoli, A. A., Keymling, M., Wehrse, E., Hog, R., Rotkopf, L. T., Wennmann, M., Glemser, P. A., Thierjung, H., von Knebel Doeberitz, N., Kleesiek, J., Gortz, M., Schutz, V., Hielscher, T., Stenzinger, A., Hohenfellner, M., Schlemmer, H. P., Maier-Hein, K. and Bonekamp, D. (2022) ‘Pseudoprospective Paraclinical Interaction of Radiology Residents With a Deep Learning System for Prostate Cancer Detection: Experience, Performance, and Identification of the Need for Intermittent Recalibration’, *Investigative Radiology,* 57(9), pp. 601-612.

#### Other cancer types

1. Ammari, S., Bone, A., Balleyguier, C., Moulton, E., Chouzenoux, E., Volk, A., Menu, Y., Bidault, F., Nicolas, F., Robert, P., Rohe, M. M. and Lassau, N. (2022) ‘Can Deep Learning Replace Gadolinium in Neuro-Oncology?: A Reader Study’, *Investigative Radiology,* 57(2), pp. 99-107.
2. Anai, K., Hayashida, Y., Ueda, I., Hozuki, E., Yoshimatsu, Y., Tsukamoto, J., Hamamura, T., Onari, N., Aoki, T. and Korogi, Y. (2022) ‘The effect of CT texture-based analysis using machine learning approaches on radiologists’ performance in differentiating focal-type autoimmune pancreatitis and pancreatic duct carcinoma’, *Japanese Journal of Radiology,* 40(11), pp. 1156-1165.
3. Erdim, C., Yardimci, A. H., Bektas, C. T., Kocak, B., Koca, S. B., Demir, H. and Kilickesmez, O. (2020) ‘Prediction of Benign and Malignant Solid Renal Masses: Machine Learning-Based CT Texture Analysis’, *Academic Radiology,* 27(10), pp. 1422-1429.
4. Malinauskaite, I., Hofmeister, J., Burgermeister, S., Neroladaki, A., Hamard, M., Montet, X. and Boudabbous, S. (2020) ‘Radiomics and Machine Learning Differentiate Soft-Tissue Lipoma and Liposarcoma Better than Musculoskeletal Radiologists’, *Sarcoma,* 2020, pp. 7163453.
5. Massa’a, R. N., Stoeckl, E. M., Lubner, M. G., Smith, D., Mao, L., Shapiro, D. D., Abel, E. J. and Wentland, A. L. (2022) ‘Differentiation of benign from malignant solid renal lesions with MRI-based radiomics and machine learning’, *Abdominal Radiology,* 47(8), pp. 2896-2904.
6. Matsuo, H., Nishio, M., Kanda, T., Kojita, Y., Kono, A. K., Hori, M., Teshima, M., Otsuki, N., Nibu, K. I. and Murakami, T. (2020) ‘Diagnostic accuracy of deep-learning with anomaly detection for a small amount of imbalanced data: discriminating malignant parotid tumors in MRI’, *Scientific Reports,* 10(1), pp. 19388.
7. Nakagawa, M., Nakaura, T., Yoshida, N., Azuma, M., Uetani, H., Nagayama, Y., Kidoh, M., Miyamoto, T., Yamashita, Y. and Hirai, T. (2023) ‘Performance of Machine Learning Methods Based on Multi-Sequence Textural Parameters Using Magnetic Resonance Imaging and Clinical Information to Differentiate Malignant and Benign Soft Tissue Tumors’, *Academic Radiology,* 30(1), pp. 83-92.
8. Okimoto, N., Yasaka, K., Kaiume, M., Kanemaru, N., Suzuki, Y. and Abe, O. (2023) ‘Improving detection performance of hepatocellular carcinoma and interobserver agreement for liver imaging reporting and data system on CT using deep learning reconstruction’, *Abdominal Radiology,* 48(4), pp. 1280-1289.
9. Shin, S. Y., Shen, T. C., Wank, S. A. and Summers, R. M. (2023) ‘Fully-automated detection of small bowel carcinoid tumors in CT scans using deep learning’, *Medical Physics,* 29, pp. 29.
10. Takeuchi, M., Seto, T., Hashimoto, M., Ichihara, N., Morimoto, Y., Kawakubo, H., Suzuki, T., Jinzaki, M., Kitagawa, Y., Miyata, H. and Sakakibara, Y. (2021) ‘Performance of a deep learning-based identification system for esophageal cancer from CT images’, *Esophagus,* 18(3), pp. 612-620.
11. Uhlig, J., Biggemann, L., Nietert, M. M., Beisbarth, T., Lotz, J., Kim, H. S., Trojan, L. and Uhlig, A. (2020) ‘Discriminating malignant and benign clinical T1 renal masses on computed tomography: A pragmatic radiomics and machine learning approach’, *Medicine,* 99(16), pp. e19725.
12. von Schacky, C. E., Wilhelm, N. J., Schafer, V. S., Leonhardt, Y., Gassert, F. G., Foreman, S. C., Gassert, F. T., Jung, M., Jungmann, P. M., Russe, M. F., Mogler, C., Knebel, C., von Eisenhart-Rothe, R., Makowski, M. R., Woertler, K., Burgkart, R. and Gersing, A. S. (2021) ‘Multitask Deep Learning for Segmentation and Classification of Primary Bone Tumors on Radiographs’, *Radiology,* 301(2), pp. 398-406.
13. von Schacky, C. E., Wilhelm, N. J., Schafer, V. S., Leonhardt, Y., Jung, M., Jungmann, P. M., Russe, M. F., Foreman, S. C., Gassert, F. G., Gassert, F. T., Schwaiger, B. J., Mogler, C., Knebel, C., von Eisenhart-Rothe, R., Makowski, M. R., Woertler, K., Burgkart, R. and Gersing, A. S. (2022) ‘Development and evaluation of machine learning models based on X-ray radiomics for the classification and differentiation of malignant and benign bone tumors’, *European Radiology,* 32(9), pp. 6247-6257.
14. Vos, M., Starmans, M. P. A., Timbergen, M. J. M., van der Voort, S. R., Padmos, G. A., Kessels, W., Niessen, W. J., van Leenders, G., Grunhagen, D. J., Sleijfer, S., Verhoef, C., Klein, S. and Visser, J. J. (2019) ‘Radiomics approach to distinguish between well differentiated liposarcomas and lipomas on MRI’, *British Journal of Surgery,* 106(13), pp. 1800-1809.
15. Weng, J., Wildman-Tobriner, B., Buda, M., Yang, J., Ho, L. M., Allen, B. C., Ehieli, W. L., Miller, C. M., Zhang, J. and Mazurowski, M. A. (2023) ‘Deep learning for classification of thyroid nodules on ultrasound: validation on an independent dataset’, *Clinical Imaging,* 99, pp. 60-66.
16. Wentland, A. L., Yamashita, R., Kino, A., Pandit, P., Shen, L., Brooke Jeffrey, R., Rubin, D. and Kamaya, A. (2023) ‘Differentiation of benign from malignant solid renal lesions using CT-based radiomics and machine learning: comparison with radiologist interpretation’, *Abdominal Radiology,* 48(2), pp. 642-648.
17. Zhang, M., Tong, E., Hamrick, F., Lee, E. H., Tam, L. T., Pendleton, C., Smith, B. W., Hug, N. F., Biswal, S., Seekins, J., Mattonen, S. A., Napel, S., Campen, C. J., Spinner, R. J., Yeom, K. W., Wilson, T. J. and Mahan, M. A. (2021) ‘Machine-Learning Approach to Differentiation of Benign and Malignant Peripheral Nerve Sheath Tumors: A Multicenter Study’, *Neurosurgery,* 89(3), pp. 509-517.
18. Zhuge, Y., Ning, H., Mathen, P., Cheng, J. Y., Krauze, A. V., Camphausen, K. and Miller, R. W. (2020) ‘Automated glioma grading on conventional MRI images using deep convolutional neural networks’, *Medical Physics,* 47(7), pp. 3044-3053.
19. Ziegelmayer, S., Reischl, S., Havrda, H., Gawlitza, J., Graf, M., Lenhart, N., Nehls, N., Lemke, T., Wilhelm, D., Lohofer, F., Burian, E., Neumann, P. A., Makowski, M. and Braren, R. (2023) ‘Development and Validation of a Deep Learning Algorithm to Differentiate Colon Carcinoma From Acute Diverticulitis in Computed Tomography Images’, *JAMA Network Open,* 6(1), pp. e2253370.

### APPENDIX 2: Titles and weblinks for ongoing or recently completed trials

1. An AI Platform Integrating Imaging Data and Models, Supporting Precision Care Through Prostate Cancer’s Continuum (ProCAncer-I). Available at: https://clinicaltrials.gov/ct2/show/record/NCT05380518
2. Does triage of chest X-rays with artificial intelligence shorten the time to lung cancer diagnosis: a randomised controlled trial. Available at: https://www.cochranelibrary.com/central/doi/10.1002/central/CN-02538677/full
3. Jager et al (2023). Clinical Trial Protocol: Developing an Image Classification Algorithm for Prostate Cancer Diagnosis on Three-dimensional Multiparametric Transrectal Ultrasound. Available at: https://www.ncbi.nlm.nih.gov/pmc/articles/PMC9975006/
4. Mammography Screening With Artificial Intelligence (MASAI) (MASAI). Available at: https://clinicaltrials.gov/study/NCT04838756
5. Diagnostic Performance of Breast Cancer Screening Second Reading Process Assisted by AI (IMA-L2). Available at: https://clinicaltrials.gov/ct2/show/record/NCT05800132
6. A Retrospective Analysis of Magnetic Resonance Imaging Data for Breast Cancer Screening in the Open Consortium for Decentralized Medical Artificial Intelligence (ODELIA). Available at: https://clinicaltrials.gov/ct2/show/record/NCT05698056
7. Detection of ISUP≥2 Prostate Cancers Using Multiparametric MRI: Prospective Multicenter Comparison of the PI-RADS Score and an Artificial Intelligence System. Available at: https://clinicaltrials.gov/ct2/show/record/NCT04732156
8. DOLCE: Determining the Impact of Optellum’s Lung Cancer Prediction (LCP) Artificial Intelligence Solution on Service Utilisation, Health Economics and Patient Outcomes. Available at: https://clinicaltrials.gov/ct2/show/record/NCT05389774
9. Artificial intelligence in mammography study. Available at: https://www.isrctn.com/ISRCTN60839016
10. Evaluating the Performance of AI in Evaluating Breast MRI Performed With Dose Reduction. Available at: https://clinicaltrials.gov/ct2/show/record/NCT04340180
11. Artificial Intelligence in Large-scale Breast Cancer Screening (ScreenTrustCAD). Available at: https://clinicaltrials.gov/ct2/show/record/NCT04778670
12. Development and validation of the AI-based diagnosis system for pathological findings in invasive front of colorectal cancer. Available at: https://upload.umin.ac.jp/cgi-open-bin/ctr_e/ctr_view.cgi?recptno=R000044488
13. Can ovarian cancer detection be improved using AI-driven diagnostic support applied to ultrasound images? Available at: https://www.isrctn.com/ISRCTN88222986
14. Automatic Detection in MRI of Prostate Cancer: DAICAP (DAICAP). Available at: https://clinicaltrials.gov/ct2/show/record/NCT05513820
15. A prospective clinical study for a rectal CRM automatic detection system based on Faster-RCNN. Available at: https://www.cochranelibrary.com/central/doi/10.1002/central/CN-02434527/full
16. Diagnostic Efficiency and Impact on Physicians’ Learning Process of an Artificial Intelligence Ultrasound Diagnosis System for Thyroid Nodules: a Multicentre Randomized Controlled Trial. Available at: https://www.cochranelibrary.com/central/doi/10.1002/central/CN-01975026/full
17. Clinical Research on a Novel Deep-learning Based System in Pancreatic Mass Diagnosis. Available at: https://www.cochranelibrary.com/central/doi/10.1002/central/CN-02183990/full
18. Development and validation of artificial intelligence-based rapid on-site cytologic evaluation during endoscopic ultrasound guided fine needle aspiration for pancreatic mass. Available at: https://www.clinicalkey.com/#!/content/playContent/1-s2.0-S0016510723015626?returnurl=https://2F2Flinkinghub.elsevier.com%2Fretrieve%2Fpii%2FS0016510723015626%3Fshowall%3Dtrue&referre=https:%2F%2Fwww.giejournal.org%2F
19. IDEAL: Artificial Intelligence and Big Data for Early Lung Cancer Diagnosis Prospective Study (Phase 2). Available at: https://clinicaltrials.gov/ct2/show/record/NCT03753724
20. New Strategies Based on Artificial Intelligence in Breast Cancer Screening Programs in Córdoba With Digital Mammography and Digital Breast Tomosynthesis. A Prospective Evaluation. Available at: https://clinicaltrials.gov/ct2/show/record/NCT04949776
21. Clinical Utility of Artificial Intelligence Augmented Endobronchial Ultrasound Elastography in Lymph Node Staging for Lung Cancer https://clinicaltrials.gov/ct2/show/record/NCT04816981

#### APPENDIX 3: Summary of Artificial intelligence (AI) models investigated as interventions

| Citation (Dataset country) | Name and type of intervention/control | AI model description | Reference standard |
| --- | --- | --- | --- |
| **Breast cancer** |  |  |  |
| **Calisto et al (2022)**<br>**Portugal** | **Intervention:** BreastScreening-AI. A DenseNet model (Deep neural network) (previously developed) + clinician<br>**Control:** Clinician only | The DenseNet was developed using PyTorch - a deep learning library that is widely used by the machine learning community.<br>The DenseNet architecture used in BreastScreening-AI was DenseNet-161. The DenseNet was initially pretrained on ImageNet, a large dataset of 1.2 million images from 1000 classes. Training was performed using the Adam optimizer, with the default parameters (learning rate of $10^{-3}$ and weight decay of $10^{-4}$). | Set by the head of radiology using BI-RADS |
| **Fujioka et al (2021)**<br>**Japan<sup>3</sup>** | **Intervention:** CNN models constructed to calculate the probability of malignancy of an image using Xception, InceptionV3, InceptionResNetV2, DenseNet121, DenseNet161, and NASNetMobile (previously developed)<br>**Control:** Human readers (a breast surgeon and a radiologist) | The CNNs were performed on DEEPstation (UEI, Tokyo, Japan) containing the graphics processing unit GeForce GTX 1080 (NVIDIA, Santa Clara, CA, USA), central processing unit Core i7-8700 (Intel, CA, USA), and graphical user interface-based DL tool Deep Analyzer (GHELIA, Tokyo, Japan).<br>The CNNs were initialized by the ImageNet (http://www.image-net.org/) pretraining model and fine-tuned to yield better performance. The parameters of optimisation were as follows: optimiser algorithm = Adam (lr = 0.0001, $\beta_1 = 0.9$, $\beta_2 = 0.999$, eps = $1e-8$, decay = 0, amsgrad = False). The image sets for training and validation phase were randomly split into training data and validation data in the ratio of 9:1 per epoch, and supervised learning with 500 epochs was performed | Histopathological examination/ follow-up |
| **Goto et al (2023)**<br>**Japan** | **Intervention:** Deep learning algorithm using pretrained Residual Networks 50 (ResNet50) architecture (previously developed).<br>**Control:** Human readers (three radiologists) | The training was implemented using the Adam optimizer fixed to 0.001. The diagnostic model was trained with a random selection of 70% of the data set. Tenfold cross-validation was performed to create the trained model. The classification performance was evaluated using 30% of the test data set. | Histopathological examination |

<sup>3</sup> If the country the dataset was obtained from was not stated within the study, the country the study was conducted in is provided

|  |  |  |  |
| --- | --- | --- | --- |
| **Heller et al (2021)**<br>**USA** | **Intervention:** A commercially available FDA-approved AI software (Koios DS; Koios Medical, New York, New York) + human reader<br>**Control:** Human readers (two radiologists) only | The Koios DS for Breast Study Tool core engine, which uses a deep learning algorithm that characterises sonographically visualised breast lesions by generating a probability of malignancy, which is in turn equated to BI-RADS categories. The software is vendor-neutral and is the first and currently only FDA-approved decision support tool based on proprietary algorithms. The algorithms are stated to be derived from both pathology proven cases or imaging follow-up obtained from more than 400,000 cases, more than 25 institutions, and multiple ultrasound machine types and vendors. | Histopathological examination |
| **Jiang et al (2021)**<br>**USA<sup>2</sup>** | **Intervention:** QuantX (AI software) + human readers<br>**Control:** Human readers (19 radiologists) alone | QuantX is a computer-assisted diagnostic MRI software aid for use in the interpretation of breast images. This system was developed at the University of Chicago and then translated and produced as QuantX at Quantitative Insights, now Qlarity Imaging (Chicago, Ill). Training and validation steps were performed on patient cases independent from the test set. | Ground truth based on final pathology reports for biopsied cancers and biopsied noncancers. Ground truth for the nonbiopsied noncancers was obtained from clinical and radiology reports and a negative follow-up MRI examination at a minimum of 12 months after the considered MRI examination |
| **Lo Gullo et al (2020)**<br>**USA** | **Intervention:** Radiomics + Machine learning model (previously developed)<br>**Control:** Human readers (two radiologists) | An in-house code written in MATLAB (The MathWorks, Inc.) was used to input the ROIs into the open-source CERR software environment (freely available through GitHub) which calculated the radiomics features. No details on training or validation provided. | Histopathology established by either image-guided needle biopsy or surgery |
| **Mango et al (2020)**<br>**USA** | **Intervention:** AI decision support system - Koios DS for Breast system (previously developed)<br>**Control:** Human readers (15 physicians) | The training data (over 400,000 clinical examples) were gathered from over 25 machines and 25 different healthcare systems and sites. The 900 cases used in this validation study were completely excluded from the testing and development of the AI system. | Pathology or imaging follow-up |
| **O'Connell et al (2022)**<br>**USA/Italy** | **Intervention:** S-Detect for Breast AI program (previously developed)<br>**Control:** Human readers (10 radiologists) | S-Detect™ for Breast is a software based on a convolutional neural network (Samsung Medison Co., Ltd., South Korea) that has been trained to classify lesions using over 10,000 breast scans against “gold standard” biopsy assessments. | Ground truth generated from biopsy or a 24-month follow-up |
| **Pacilè et al (2020)**<br>**USA** | **Intervention:** Human readers + AI (MammoScreen V1; Therapixel, Nice, France) – previously developed<br>**Control:** Human readers (14 radiologists) | The AI system used (MammoScreen V1; Therapixel, Nice, France) uses two groups of deep convolutional neural networks (CNN) combined together with an aggregation module. The (symmetric) dream-nets were trained directly from images and their status (negative or positive). The detection CNN was trained from images and their annotations regardless of the statuses, while the (symmetric) characterization CNN were trained from image annotations and their status. | Histopathological examination |
| **Pinto et al (2021)**<br>**Netherlands** | **Intervention:** Human reader + AI CAD system (Transpara. version 1.6.0; ScreenPoint Medical) – previously developed<br>**Control:** Human readers (14 radiologists) | The AI system identifies suspicious regions and assigns to them a score between 1 and 100 representing the level of suspicion (LoS) of cancer present (100 indicates the highest suspicion). Information about the AI training and validation not provided. | One-year normal follow-up or histopathologic assessment |
| **Tsochatzidis et al (2019)**<br>**USA** | **Intervention:** Deep convolutional neural networks (CNNs) – AlexNet, VGG, GoogLeNet/Inception, Residual Networks (ResNets) – previously developed<br>**Control:** AI models above were compared amongst themselves | For the training process the Adam optimization method was employed. | Not stated |
| **Uhlig et al (2018)**<br>**Germany** | **Intervention:** Five machine learning techniques (random forests, back propagation neural networks (BPN), extreme learning machines, support | Implementation of machine learning techniques comprises two steps: an initial training step, using the clinical dataset with an input-output pair to train the model, followed by a testing step, where the performance of the prediction model is tested on new data not used for training and therefore unknown to the model. | Core needle biopsy with subsequent histopathologic evaluation for |
|  | vector machines, and K-nearest neighbors) – previously developed<br>**Control:** Human readers (two readers) |  | breast lesions rated as BI-RADS category 4 or 5 by at least one reader or no evidence of size progression on imaging follow-up for at least 12 months for breast lesions rated as BI-RADS categories 1–3 by both readers |
| **Vamvakas et al (2022)**<br>**Greece** | **Intervention:** Four popular implementations of Decision Trees (DT) Boosting classifiers, namely Adaptive Boosting (AdaBoost), Gradient Boosting (GB), Extreme Gradient Boosting (XGBoost), and Light Gradient Boosting Machine (LightGBM) – previously developed<br>**Control:** An SVM classifier | Python implementations for XGBoost and LightGBM were obtained from their original sources [https://github.com/dmlc/xgboost], [https://github.com/microsoft/LightGBM] and used through the scikit-learn Application Programming Interface (API), which is a common framework for ML applications. The final feature subset was used to train the GB, AdaBoost, XGBoost, LightGBM and SVM classifiers in differentiating benign from malignant breast lesions. | Histological verification from core needle biopsy or surgical excision |
| **van Zelst et al (2020)**<br>**Netherlands<sup>2</sup>** | **Intervention:** AI model not stated. Computer-aided detection (CAD) software used, a commercially developed ABUS CAD software package (QVCAD, Qview Medical Inc., Los Altos, CA, USA)<br>**Control:** Human readers (eight radiologists) | Information about the AI training not provided. | Histopathologic examination |
| **Prostate cancer** |  |  |  |
| **Akatsuka et al (2019)**<br>**Japan** | **Intervention:** Previously developed Deep convolutional neural network (dCNN)(Xception) | Three deep convolutional neural network models, Xception, inceptionV3 and VGG16, were pre-trained on ImageNet with classification layers | Histopathologic diagnosis |
|  | **Control:** Human readers (radiologists) | adapted to the study's labels. Xception was selected in this study because it showed the most precise performance for MR image classification. 10-fold cross-validation was used to test the prediction models, randomly dividing the whole cases in a 1:9 ratio, using one part for testing and the other nine parts for training |  |
| **Arslan et al (2023)**<br>**Turkey** | **Intervention:** Human + Commercially available DL software (Prostate AI, Version Syngo.Via VB60, Siemens Healthcare)<br>**Control:** Human readers (four radiologists) | The DL software used in this study has three modules: (i) preprocessing module, (ii) DL-based lesion detection module, and (iii) DL-based lesion classification module.<br>No model training or fine-tuning was performed in this study. The model was only used for performance testing. | Whole-mount pathology or MRI/ultrasound fusion-guided biopsy |
| **Faiella et al (2022)**<br>**Italy** | **Intervention:** Previously developed AI software Quantib Prostate (Quantib B.V., Rotterdam, The Netherlands) + human (inexperienced radiologist)<br>**Control:** Human reader (expert radiologist) | Quantib Prostate offers features for the reading of prostate MRI in one workflow. The semi-automated combination of bi-parametric data provided supports Region of Interest (ROI) determination and enables prostate lesion evaluation.<br>No information about AI training and validation provided. | Biopsy results and radiology reports |
| **Forookhi et al (2023)**<br>**Italy** | **Intervention:** Human + commercially available AI-assisted software (Quantib® Prostate)<br>**Control:** Human readers (four novice readers) | No information about AI training and validation provided. | Reports from the expert radiologist |
| **Patsanis et al (2023)**<br>**Norway** | **Intervention:** Six previously developed relevant 2D GANs were selected for investigation: f-AnoGAN, HealthyGAN, StarGAN, StarGAN-v2, FP-GAN and DeScarGAN. For each model, the code was publicly available.<br>**Control:** AI models above were compared amongst themselves | For each model, the code was publicly available. All input annotations for training were on the image level, i.e., each input image was labeled as negative or positive. All models were trained to generate 2D images representing a negative patient. | Manual delineations of PI-RADS $\geq 3$ lesions by imaging experts /radiologists based on targeted biopsy |
| **Tong et al (2023)**<br>**USA<sup>2</sup>** | **Intervention:** A commercially developed proprietary deep learning-based prototypical computer-aided detection algorithm (DL-CAD) (MR | The deep learning-based reconstruction algorithm was trained in a supervised manner using more than 10,000 slices of fully sampled T2 TSE acquisitions obtained from volunteers using 1.5 T and 3 T MR scanning systems (MAGNETOM scanners, Siemens Healthcare) of various regions | Adequate follow up was defined as a prostate biopsy within 1 |
|  | Prostate AI, version 1.3.2, build July 07, 2021, front end build November 06, 2019, Siemens Healthcare)<br>**Control:** Human readers (three abdominal fellowship trained radiologists) | of the body, including head, pelvis, and knee. The training was implemented in PyTorch and performed using a commercially available GPU cluster with 32 GB of memory. | year of the MRI or stability of PSA of at least 1 year if mpMRI was prospectively determined to be PI-RADS 1 or 2. |
| **Zhang et al (2022)**<br>**Germany** | **Intervention:** A previously established and validated DL algorithm, convolutional neural network (CNN)<br>**Control:** Human readers (radiologists-in-training) | The utilized deep learning (DL) system is based on a task-specific CNN U-Net architecture and was previously retrospectively trained and validated using single-scanner biparametric data (T2-weighted and diffusion-weighted images), demonstrating similar performance to clinical PI-RADS assessment. | Combined extended systematic and targeted MRI/TRUS-fusion biopsy. |
| **Lung cancer** |  |  |  |
| **Baldwin et al (2020)**<br>**UK** | **Intervention:** Previously developed AI model (Lung Cancer Prediction CNN (LCP-CNN)<br>**Control:** Brock model | The LCP-CNN is an AI model that analyses parts of a CT scan around a nodule of interest and provides a score from 0 to 100 for that nodule.<br>In the first phase of training, the LCP-CNN was primed using hundreds of thousands of images selected and curated carefully to teach the network the kinds of visual discrimination tasks that may form the building blocks for a nodule discrimination task. The second phase comprised full supervised training on a version of the NLST data. This was performed on CT images of all solid and semi-solid nodules of at least 6mm in diameter from the NLST dataset that were not reported as pure ground glass opacities (GGO); all GGOs were excluded because there were too few examples of malignant GGOs to train the system reliably.<br>The derivation and internal validation of the LCP-CNN was performed using eight-fold cross-validation, where all images and nodules for a given patient were assigned to the same fold, and in each of the eight training operations, one eighth of the data were reserved as an auxiliary set for convergence testing and parameter/threshold setting, and one final eighth was kept for internal validation. Because the NLST contains many more benign nodules than cancers, class balancing was used during training, since otherwise the resulting network would be tuned always to predict "benign", since that would be the dominant class. | Ground truth (based on histology (required for any cancer), resolution, stability or (for pulmonary lymph nodes only) expert opinion |
| **Jacobs et al (2021)**<br>**USA/Canada/Netherlands/Belgium** | **Intervention:** Three top-performing algorithms from the Kaggle Data Science Bowl 2017 public competition: grt123, Julian de Wit and Daniel Hammack (JWDH), and Aidence (all pPreviously developed deep learning algorithms)<br>**Control:** Human readers (11 radiologists) | No information about AI training and validation provided. | Set by histopathologic examination for cancer-positive scans and imaging follow-up for at least 2 years for cancer-negative scans |
| **Maldonado et al (2021)**<br>**USA** | **Intervention:** BRODERS classifier used by CANARY AI software<br>**Control:** Brock model | The BRODERS classifier (Benign versus aggrressive nODule Evaluation using Radiomic Stratification) is a conventional predictive radiomic model based on eight imaging features capturing nodule location, shape, size, texture and surface characteristics. A training set of 726 incidentally detected indeterminate pulmonary nodules (IPNs) from the NLST database was used to internally validate the BRODERS classifier | Final diagnosis |
| **Tam et al (2021)**<br>**UK** | **Intervention:** A commercially available AI algorithm (Red Dot, Behold.ai, London, UK) +/- radiologists<br>**Control:** Human readers (radiologists) | No information about AI training and validation provided. | The ground-truth of each examination was established by a combination of the cancer registry database records, the electronic clinical record, and review of both subsequent and prior imaging. |
| **Toğaçar et al (2020)**<br>**Turkey<sup>2</sup>** | **Intervention:** Previously developed AI models LeNet, AlexNet and VGG-16 CNNs.<br>**Control:** AI models above were compared amongst themselves | In training of the deep models, RMSprop, ADAM and Stochastic Gradient Descent (SGD) optimization methods were examined. | Each sample in the dataset was examined and labeled by experienced specialists. |
| **Ueda et al (2021)**<br>**Japan** | **Intervention:** The AI-based CAD used in this study is EIRL Chest X-ray Lung nodule (LPIXEL Inc.), commercially available in Japan as of August 2020<br>**Control:** Human readers (Eighteen readers - nine general physicians and nine radiologists) | The CAD was developed based on an encoder-decoder network categorizing segmentation technique in DL. No information about AI training and validation provided. | Set by two author radiologists |
| **Wataya et al (2023)**<br>**Japan** | **Intervention:** Human + Previously developed AI model pulmonary nodule CAD system attached to SYNAPSE SAI Viewer V1.4 (FUJIFILM Corporation).<br>**Control:** Human readers (15 radiologists) | No information about AI training and validation provided. | Established through the agreement between two board-certificated chest radiologists with 17 and 25 years of diagnostic experience. |
| **Abbreviations:** Artificial intelligence (AI), Breast Imaging Reporting and Data System (BI-RADS), Back Propagation Neural Networks (BPN), Convolutional Neural Network (CNN), Computer Aided Diagnosis (CAD), DT (Decision Trees), Deep convolutional neural network (DCNN), Deep Learning (DL), Deep Learning Computer Aided Diagnosis (DLCAD), The United States Food and Drug Administration (FDA), Gradient Boosting (GB), Lung Cancer Prediction Convolutional Neural Network (LCP-CNN), Region of Interest (ROI), Level of suspicion (LOS), Stochastic Gradient Descent (SGD), Support Vector Machine (SVM), Prostate Imaging Reporting and Data System (PI-RADS). |  |  |  |

### APPENDIX 4: MEDLINE search strategy

Ovid MEDLINE(R) ALL <1946 to June 19, 2023>

1. (“artificial intelligence” or AI).tw. 61802
2. (radiomic* or “machine learning” or “deep learning” or “neural network*”).tw. 182269
3. artificial intelligence/ or machine learning/ 68965
4. or/1-3 250797
5. (diagnos* adj3 imag*).tw. 54007
6. “diagnostic aid*”.tw. 3847
7. exp Diagnostic imaging/ 2912910
8. (“medical imaging” adj3 diagnos*).tw. 504
9. ((X-ray or CT or MRI or PET or CBCT or MRCP or MIBG or MRS or ultrasound) adj5 diagnos*).tw. 95951
10. mammogra*.tw. 36225
11. ((“positron emission tomography” or “computed tomography”) adj5 diagnos*).tw. 19194
12. (“Metaiodobenzylguanidine scan” adj5 diagnos*).tw. 3
13. (“Magnetic resonance” adj (spectroscopy or imaging or angiogram*) adj5 diagnos*).tw. 13135
14. ((cerebral or brain) adj angiogram* adj5 diagnos*).tw. 144
15. or/5-14 2977628
16. (neoplas* or tumo?r* or malignan* or cancer* or carcinoma* or adenocarcinoma* or melanoma* or lymphoma* or myeloma* or sarcoma*).tw. 4122728
17. exp Neoplasms/ 3844620
18. 16 or 17 5092487
19. 4 and 15 and 18 11866
20. (case reports or comment or editorial or letter or review or systematic review or meta analysis).pt. 7437910
21. (“systematic review” or meta-analysis or meta-analyses).ti. 301263
22. 20 or 21 7474118
23. 19 not 22 10317
24. limit 23 to (english language and yr=“2018 -Current”) 7881
25. afghanistan/ or africa/ or africa, northern/ or africa, central/ or africa, eastern/ or “africa south of the sahara”/ or africa, southern/ or africa, western/ or albania/ or algeria/ or andorra/ or angola/ or “antigua and barbuda”/ or argentina/ or armenia/ or azerbaijan/ or bahamas/ or bahrain/ or bangladesh/ or barbados/ or belize/ or benin/ or bhutan/ or bolivia/ or borneo/ or “bosnia and herzegovina”/ or botswana/ or brazil/ or brunei/ or bulgaria/ or burkina faso/ or burundi/ or cabo verde/ or cambodia/ or cameroon/ or central african republic/ or chad/ or exp china/ or comoros/ or congo/ or cote d’ivoire/ or croatia/ or cuba/ or “democratic republic of the congo”/ or cyprus/ or djibouti/ or dominica/ or dominican republic/ or ecuador/ or egypt/ or el salvador/ or equatorial guinea/ or eritrea/ or eswatini/ or ethiopia/ or fiji/ or gabon/ or gambia/ or “georgia (republic)”/ or ghana/ or grenada/ or guatemala/ or guinea/ or guinea-bissau/ or guyana/ or haiti/ or honduras/ or independent state of samoa/ or exp india/ or indian ocean islands/ or indochina/ or indonesia/ or iran/ or iraq/ or jamaica/ or jordan/ or kazakhstan/ or kenya/ or kosovo/ or kuwait/ or kyrgyzstan/ or laos/ or lebanon/ or liechtenstein/ or lesotho/ or liberia/ or libya/ or madagascar/ or malaysia/ or malawi/ or mali/ or malta/ or mauritania/ or mauritius/ or mekong valley/ or melanesia/ or micronesia/ or monaco/ or mongolia/ or montenegro/ or morocco/ or mozambique/ or myanmar/ or namibia/ or nepal/ or nicaragua/ or niger/ or nigeria/ or oman/ or pakistan/ or palau/ or exp panama/ or papua new guinea/ or paraguay/ or peru/ or philippines/ or qatar/ or “republic of belarus”/ or “republic of north macedonia”/ or romania/ or exp russia/ or rwanda/ or “saint kitts and nevis”/ or saint lucia/ or “saint vincent and the grenadines”/ or “sao tome and principe”/ or saudi arabia/ or serbia/ or sierra leone/ or senegal/ or seychelles/ or singapore/ or somalia/ or south africa/ or south sudan/ or sri lanka/ or sudan/ or suriname/ or syria/ or taiwan/ or tajikistan/ or tanzania/ or thailand/ or timor-leste/ or togo/ or tonga/ or “trinidad and tobago”/ or tunisia/ or turkmenistan/ or uganda/ or ukraine/ or united arab emirates/ or uruguay/ or uzbekistan/ or vanuatu/ or venezuela/ or vietnam/ or west indies/ or yemen/ or zambia/ or zimbabwe/ 1291789
26. “Organisation for Economic Co-Operation and Development”/ 539
27. australasia/ or exp australia/ or austria/ or baltic states/ or belgium/ or exp canada/ or chile/ or colombia/ or costa rica/ or czech republic/ or exp denmark/ or estonia/ or europe/ or finland/ or exp france/ or exp germany/ or greece/ or hungary/ or iceland/ or ireland/ or israel/ or exp italy/ or exp japan/ or korea/ or latvia/ or lithuania/ or luxembourg/ or mexico/ or netherlands/ or new zealand/ or north america/ or exp norway/ or poland/ or portugal/ or exp “republic of korea”/ or “scandinavian and nordic countries”/ or slovakia/ or slovenia/ or spain/ or sweden/ or switzerland/ or turkey/ or exp united kingdom/ or exp united states/ 3490253
28. European Union/ 17665
29. Developed Countries/ 21362
30. or/26-29 3506177
31. 25 not 30 1202341
32. 24 not 31 7813

## Footnotes

1 For the purposes of this review, an AI model was classed as commercially available if it was stated as such in the primary study. An AI model was classed as previously developed if it was stated as such in the primary study or if it was made clear that the model had not been developed for use in the study.

2 The direction of effect was determined based on whether the results were statistically significant.

3 If the country the dataset was obtained from was not stated within the study, the country the study was conducted in is provided

